# Trajectories matter: Discovery and validation of ordered EHR sequences that inform clinical risk predictions

**DOI:** 10.1101/2025.09.14.25335720

**Authors:** Brice Edelman, Hannah Kim, Jeffrey Skolnick

## Abstract

**Objective:** To test whether temporally ordered clinical sequences mined from EHRs improve the understanding of downstream adverse outcomes versus unordered bag-of-codes representations.

**Materials and Methods:** Within the NIH All of Us Controlled Tier, we mined frequent ordered event pairs (A→B) and tested for risk elevation of later adverse events (C) to create three-event (A→B→C) sequences. The algorithm used observation-period–aware indexing, a maximum 5-year A→B gap, and 90-day latency before follow-up. Comparators were (B without prior A)→C (primary), B→A→C, (A without B)→C, and a calendar-time baseline. Covariates were balanced with inverse probability of treatment weighting (IPTW) Weighted Aalen–Johansen estimated cumulative incidence at 1, 2, and 5 years. Discovery and confirmation were analyzed separately with global Benjamini-Hochberg false discover rate (BH-FDR).

**Results:** Of 633,545 persons, 432,617 met eligibility (≥365 observed days) and were split 70/30 into discovery and confirmation. We mined 3,066,183 trajectories from the discovery set by combining 340,687 sufficiently supported A→B pairs with each of 9 curated adverse third events.

Due to compute constraints, we tested 20,565 of these trajectories (0.67%) for risk elevation. After discovery FDR, 234 trajectories advanced, and 39 validated for at least one time horizon in confirmation. At 5 years, the median risk ratio (RR) was 3.96 versus baseline and 2.18 versus B→C with no prior A. Reverse-order checks were feasible for 89.7% of hypotheses; the median (A→B→C) vs (B→A→C) RR was 1.21.

**Discussion:** Ordered trajectories captured clinically coherent pathways where temporal order added information beyond diagnosis presence alone.

**Conclusion:** Trajectory mining and confirmation reveal actionable, risk pathways that complement conventional risk models.

## Background and Significance

Quantitative risk stratification is central to modern clinical decision-making.[1, 2] Foundational calculators such as the Framingham general cardiovascular risk score and CHA₂DS₂-VASc for atrial fibrillation stroke risk demonstrated that a compact set of clinical factors can yield actionable estimates that guide prevention and treatment pathways.[3, 4] These tools, however, summarize risk at a single point in time and typically do not encode how a patient’s history unfolds.

The digitization of health systems has subsequently enabled learning from large-scale electronic health records (EHRs). Machine-learning models trained on routine clinical data can improve discrimination for important outcomes, and unsupervised representations derived from EHRs have shown broad predictive utility across disease areas.[5, 6] At the same time, influential sequence models (e.g., recurrent neural networks and attention mechanisms) learn from longitudinal visit data and can forecast diagnoses and medications.[7–13] Yet, in many deployed or benchmarked settings, patient histories are still featurized as unordered sets or counts of codes (“bag-of-codes”), which can obscure the role of temporal order; where temporal models are used, their end-to-end complexity can impede interpretability and clinical auditability, raising concerns in high-stakes settings.[14] Our premise is that bridging this gap requires methods that explicitly test whether the order of events carries incremental prognostic information while remaining transparent enough for clinical scrutiny.

Concurrently, systems-level studies have reframed disease as a set of interlinked processes that progress along structured pathways.[15] Network medicine shows that disorders cluster within biological modules, and population-level “disease networks” reveal empiric comorbidity patterns across the phenome.[16, 17] Phenotypic disease networks constructed from tens of millions of medical records suggest that patients tend to develop conditions adjacent to those they already have and that network position relates to mortality risk.[18] Population-scale registry analyses in Denmark distilled thousands of statistically supported, directional disease trajectories (i.e. evidence that the sequence in which conditions arise is not random) and released tools to explore these trajectories at scale.[19, 20] Together, this literature motivates a trajectory-aware view of risk that complements molecular and clinical perspectives.

Methodologically, observational prediction from longitudinal EHRs must also guard against time-related biases that can masquerade as temporal effects. For example, immortal time bias and misaligned “time zero” can inflate or invert associations.[21, 22] In addition, many representation methods compress histories in ways that ignore cross-visit order, making it difficult to assess whether risk is driven by the presence of two conditions or by their ordering.[6]

In this study, we operationalize an interpretable, trajectory-aware approach: we mine frequent ordered pairs of clinical events (A→B) in a large, diverse U.S. cohort from the NIH All of Us Research Program and rigorously test whether specific orderings elevate downstream risk of subsequent adverse health events (C) relative to clinically relevant comparators. By quantifying the incremental value of temporal order beyond their “bag-of-codes” presence, we aim to connect population-scale A→B→C disease trajectories with patient-level risk calibration in a way that is both statistically principled and clinically interpretable.

## Materials and Methods

We conducted a retrospective cohort study in the NIH All of Us (Aou) Research Program Controlled Tier environment.[23] Analyses were run inside the AoU Workbench against the Observational Medical Outcomes Partnership (OMOP) Common Data Model using Google BigQuery and a Python stack.

Eligibility required ≥365 total days of observation after merging overlapping or adjacent observation periods per person. A given BigQuery query computes person-level intervals of observation, totals days across merged intervals, and selects eligible participants (≥365 days). All observation intervals for eligible persons are then retrieved and cached for downstream, interval-aware censoring. The observation intervals are normalized to datetimes and validated (drop missing/invalid; ensure start ≤ end) before use in episode building.

Eligible persons were split 70/30 into discovery and confirmation sets using stratified sampling by age band (decades), sex at birth, and race/ethnicity; assignments were cached for reproducibility.

We evaluated a curated panel of clinically important outcomes to use as sequence-end events, defined as OMOP ancestor concept sets resolved via the vocabulary tables. The configuration file enumerates outcomes and stores their ancestor concept identifiers for descendant expansion at phenotype time. The full set of adverse “final” outcomes analyzed in this study are breast cancer, ovarian cancer, colorectal cancer, lung cancer, leukemia, lymphoma, myocardial infarction, dementia, and death.

For time-to-event analyses, all non-death outcomes treat death as a competing event; for the “death” outcome, there is no competing event. On exact ties between an outcome and death on the same day, the tie-breaking rule assigns death to occur infinitesimally earlier to avoid misclassification.

The notebook constructs a curated event vocabulary that maps source OMOP concepts to higher-level, analysis-friendly features in the form of a CSV file of manually curated features plus descendants via concept_ancestor. This CSV file can be changed between analyses to determine the set of conditions, drug exposures, or procedures being investigated for significant links to the terminal outcomes. A “blacklist” removes any feature that descends from the outcome definitions to avoid label leakage. The ledger of person-level events (events_df) is built by joining the curated vocabulary to OMOP domain tables and by restricting event dates to observation periods. Deduplication yields one record per (person, date, feature).

Two derived, accessibility-oriented covariate events are also added to the ledger: smoking status (from AoU Participant-Provided Information (PPI) survey responses) and BMI (body mass index) categories (from measurements), each mapped to explicit derived concept identifiers to support categorical modeling.

In the discovery set, the notebook mines begins by ordering pairs of events A→B using first-observed dates per person. A minimum support threshold ensures statistical power and stability of downstream analyses. The configuration used for this report requires at least 500 occurrences (globally and within key strata) for a candidate sequence to be retained for hypothesis testing.

For each candidate A→B and for each outcome, the engine builds episodes for:

- Sequence: A→B (B strictly after A by default),
- Primary comparator: B with no prior A,
- Reverse: B→A (temporal order flipped),
- Tertiary: A with no subsequent B within the allowed gap, and
- Baseline: one calendar-time–anchored episode per person.

When building sequences, we excluded same-day events by default (i.e., B must be strictly after A), and the maximum allowed A→B (or B→A) gap is 1825 days (5 years). These policies are applied consistently when deriving all comparators.

Each episode has an index date and a time zero defined as index_date + latency, with a default latency of 90 days to mitigate reverse causation. A key design guardrail requires that both index date and time zero fall inside the same observation interval, and censoring occurs at that interval’s end. For the B-no-prior-A, A-no-B, and Baseline cohorts, the interval must start at least 1825 days (5 years) before the index date to ensure adequate lookback for covariates and history checks.

The Baseline comparator uses a single observation period (OP)-aware index per person, set to the observation period start plus the lookback requirement, with time zero required to be in the same interval (and thus comparable calendar time anchor). Baseline represents each individual’s background risk, independent of A or B: for each person, we anchor a single episode at the observation-period start plus the required 5-year lookback, then begin follow-up 90 days later, ensuring both dates lie within the same interval.

For each (person, index date), the notebook derives covariates anchored at time zero:

- Smoking status and BMI category: the most recent value within 730 days prior to time zero (fallback to “Unknown” if none).
- Healthcare utilization: count of distinct visits in the prior 365 days.
- Socioeconomic status: deprivation index at the person level (with median imputation if missing).
- Comorbidity burden: Charlson Comorbidity Index (leave-one-out, CCI-LOO) computed over 730 days before time zero and explicitly excluding any Charlson categories that descend from A, B, or the outcome’s ancestor set, to avoid overadjustment on pathway-defining concepts.

The covariate engine outputs a table keyed by person and index date and is merged into each cohort’s episode table before modeling.

To adjust for confounding, we fit logistic regression propensity models separately for each comparison—Sequence vs B-no-prior-A (primary), Sequence vs B→A (secondary), Sequence vs A-no-B (tertiary), and Sequence vs Baseline—including the covariates: age at index, sex at birth, race/ethnicity, calendar year of index, smoking status, BMI category, deprivation index, healthcare utilization, and CCI-LOO. Categorical covariates are one-hot encoded; Inverse association IPTW = 1/PS for treated and 1/(1–PS) for control, clipped at the 99th percentile to stabilize extremes. Weights are attached to episode rows and carried into survival analyses.

We estimate cumulative incidence functions (CIFs) for each possible event C from the curated outcomes list at 1, 2, and 5 years after time zero using a weighted Aalen–Johansen (AJ) estimator on integer days. For non-death outcomes, death is treated as a competing event (tied same-day events are attributed to death); for the death outcome there is no competitor. Confidence intervals and two-sided p-values for the risk difference (ΔCIF) calculated with respect to an alternative scenario (e.g. with respect to baseline) use a Poisson bootstrap with early stopping (start 100 draws, up to 250; stop when the ΔCIF CI half-width stabilizes to ≤0.2 percentage-points absolute or ≤5% relative). For each horizon, we report CIFs for C given A→B and the comparator, ΔCIF, and the risk ratio (CIF_(A→B→C)/CIF_(comp)). The code also implements a Greenwood-type variance approximation, but the analysis used in this report was configured to use the Poisson bootstrap method.

To maintain reliability, analyses at a given horizon are gated to ensure adequate information: minimum at-risk counts (≥150) and minimum observed events (≥15) per group (with optional effective sample size checks based on weight dispersion).

In discovery, we compute p-values per outcome × horizon and then apply global Benjamini– Hochberg False Discovery Rate (FDR) control at α=0.05 across all discovery tests to obtain q-values. We then apply simple post-hoc effect-size / directionality triage—retaining only hypotheses with risk ratio >1.2, ΔCIF >0.5 percentage-points, p<0.05 versus the primary comparator, and risk ratio >1.1 versus the reverse comparator (A→B→C > B→A→C)—to form a frozen list for confirmation.

(Applying triage after FDR only removes rejections and does not inflate FDR.) In confirmation, we rebuild episodes, recompute covariates and PS weights, and repeat the competing-risk analysis for the frozen set of (sequence, outcome, horizon) hypotheses. We then apply global BH FDR across all confirmation p-values to produce q-values reported in the results table.

Pre-specified sensitivity analyses vary latency (30, 90, 180 days) and maximum allowed A→B gap (365, 1825 [primary], 3650 days), while enforcing the same observation-interval rules (index and time zero inside the same interval with adequate lookback). These analyses are run for the top confirmed signals and recompute covariates, weights, and CIFs (including the reverse comparator).

All counts and tables obey AoU Controlled Tier dissemination rules: cells with counts <20 are suppressed; all other reported counts are rounded down to the nearest multiple of 5. Suppression propagates to derived estimates (e.g., CIFs, ΔCIF, q-values) in the public table to prevent back-calculation of small cells. The notebook records configuration (e.g., RNG seed, risk horizons, gating thresholds, SAME_DAY policy, gap and latency parameters) and writes intermediate artifacts (e.g., mined sequences, episodes, raw/confirmed results) for resumability.

## Results

### Cohort, mining, and confirmation

From 633,545 persons in the OMOP source, 432,617 met eligibility criteria (≥365 observation days) and were split 70/30 into discovery (302,834) and confirmation (129,783) sets. We mined 340,687 frequent ordered pairs of clinical events (A→B) in discovery under a ≥500-support threshold, then combined them with each of the adverse outcomes to create 3,066,183 total three-event trajectories for evaluation.

Due to compute limitations, we were able to test 20,565 trajectories (0.67%) for risk elevation of the third event (the adverse outcome) in this preliminary analysis. Each trajectory is evaluated for statistically significant risk elevation of the adverse outcome at three different time horizons: 1 year, 2 years, and 5 years. 367 trajectory-horizon combinations (234 unique trajectories with at least one significant time horizon) passed the discovery-set false discovery rate (FDR) of 0.05 and advanced to confirmation. 68 trajectory-horizon combinations, which included 39 different three-event trajectories significant for at least one time horizon, were validated under global FDR control in the confirmation set.

It is critical to note that only 0.67% of potential trajectories have been tested so far. An extrapolation from this sample would indicate approximately 5,500 to 6,000 trajectories remain to be discovered using our approach given more computational time.

All counts obey All of Us dissemination rules: cells corresponding to less than 20 cases (or statistics that refer to 20 or fewer patients) are suppressed; otherwise, counts are rounded down to the nearest multiple of 5.

**Table 1.**
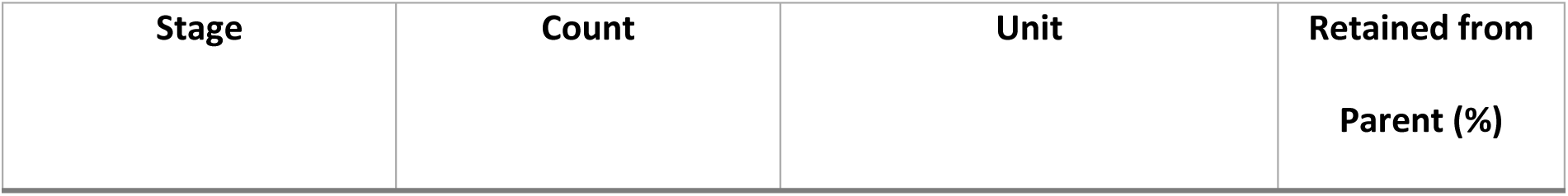
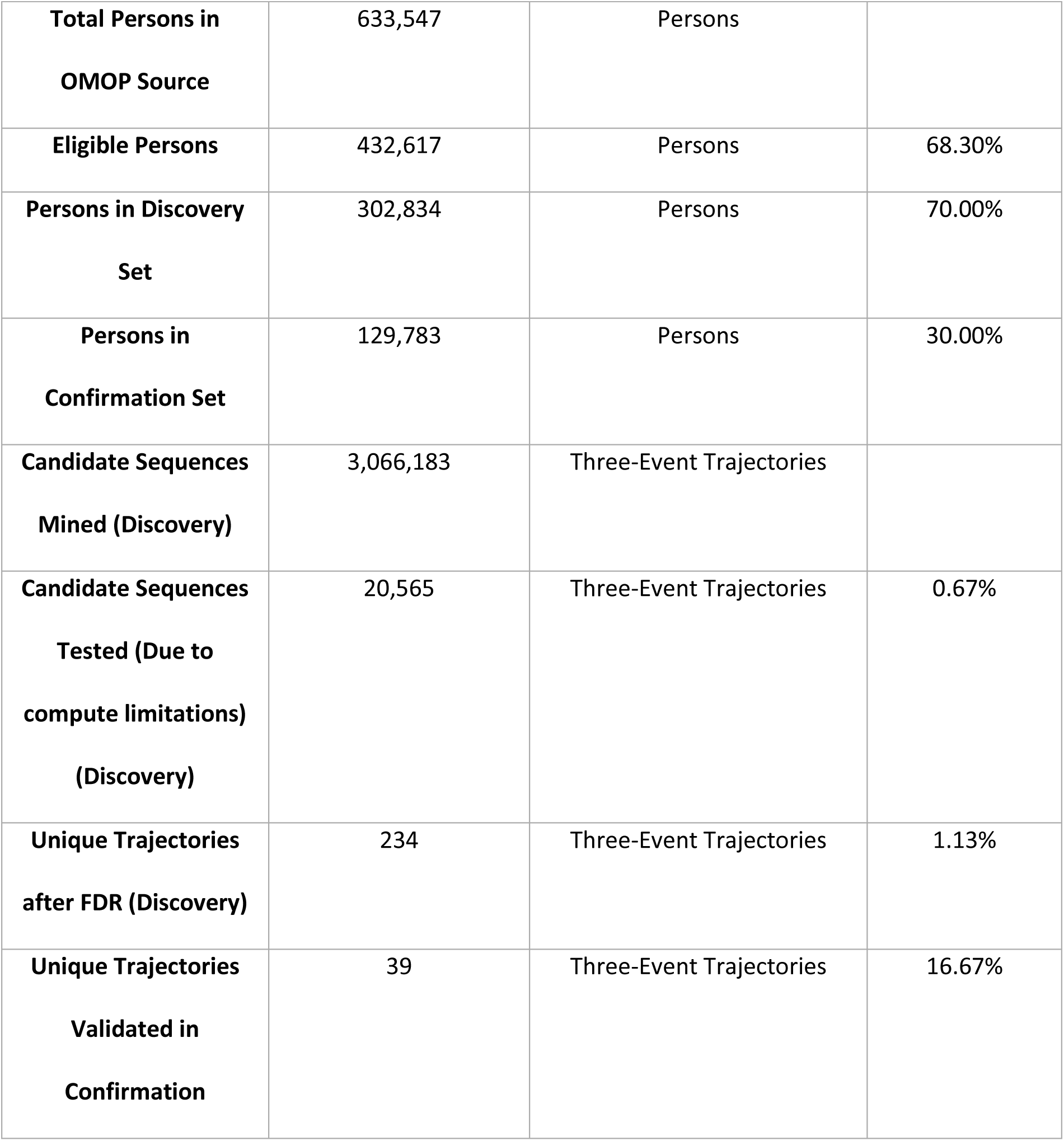
Attrition of persons, sequences, and hypotheses between steps.

| Stage | Count | Unit | Retained from<br>Parent (%) |
| --- | --- | --- | --- |
| **Total Persons in OMOP Source** | 633,547 | Persons |  |
| **Eligible Persons** | 432,617 | Persons | 68.30% |
| **Persons in Discovery Set** | 302,834 | Persons | 70.00% |
| **Persons in Confirmation Set** | 129,783 | Persons | 30.00% |
| **Candidate Sequences Mined (Discovery)** | 3,066,183 | Three-Event Trajectories |  |
| **Candidate Sequences Tested (Due to compute limitations) (Discovery)** | 20,565 | Three-Event Trajectories | 0.67% |
| **Unique Trajectories after FDR (Discovery)** | 234 | Three-Event Trajectories | 1.13% |
| **Unique Trajectories Validated in Confirmation** | 39 | Three-Event Trajectories | 16.67% |

### Overview of validated trajectory–outcome signals

Validated signals spanned 39 distinct A→B→C trajectories carrying significantly elevated risk for at least one time horizon. Outcomes covered six clinically important endpoints: myocardial infarction (MI; 19 sequence pairs), lung cancer (10), dementia (7), and single sequence pairs for colorectal cancer, breast cancer, and leukemia. At the 5-year horizon, the risk-ratio (RR) distribution had a median of 2.18 (IQR 1.76–2.65; maximum 5.28), and the absolute risk differences (ΔCIF, sequence vs the primary comparator “(B without prior A)→C”) had a median of 2.3 percentage-points (pp) (Interquartile Range (IQR) 1.4–3.7 pp) and a maximum of 5.9 pp.

**Table 2.** shown below, provides aggregated summary statistics for the validated signals across outcome events and horizons.

| Outcome | Horizon<br>(Years) | Validated<br>Signals (n) | ΔCIF —<br>Median<br>[IQR] | Risk<br>Ratio (vs<br>Primary)<br>—<br>Median<br>[IQR] | Risk<br>Ratio (vs<br>Reverse)<br>—<br>Median<br>[IQR] | Risk<br>Ratio<br>(vs A,<br>no<br>subseq.<br>B) —<br>Median<br>[IQR] | Risk<br>Ratio (vs<br>Baseline)<br>—<br>Median<br>[IQR] |
| --- | --- | --- | --- | --- | --- | --- | --- |
| breast_cancer | 5 | 1 | 1.41%<br>[1.41%,<br>1.41%] | 2.08<br>[2.08,<br>2.08] | 2.13<br>[2.13,<br>2.13] | 1.42<br>[1.42,<br>1.42] | 1.94<br>[1.94,<br>1.94] |
| colorectal_cancer | 5 | 1 | 0.87%<br>[0.87%,<br>0.87%] | 2.94<br>[2.94,<br>2.94] | 0.85<br>[0.85,<br>0.85] | 1.02<br>[1.02,<br>1.02] | 3.82<br>[3.82,<br>3.82] |
| **dementia** | 2 | 4 | 0.80%<br>[0.65%,<br>0.99%] | 2.05<br>[1.92,<br>2.23] | 1.04<br>[0.89,<br>1.23] | 1.22<br>[1.08,<br>1.37] | 4.16<br>[3.20,<br>5.36] |
| **dementia** | 5 | 6 | 1.96%<br>[1.77%,<br>2.22%] | 1.86<br>[1.77,<br>2.06] | 1.30<br>[1.13,<br>1.38] | 1.59<br>[1.53,<br>1.83] | 4.25<br>[3.28,<br>4.71] |
| **leukemia** | 5 | 1 | 0.92%<br>[0.92%,<br>0.92%] | 4.45<br>[4.45,<br>4.45] | 1.41<br>[1.41,<br>1.41] | 1.57<br>[1.57,<br>1.57] | 4.04<br>[4.04,<br>4.04] |
| **lung_cancer** | 1 | 3 | 0.78%<br>[0.60%,<br>0.86%] | 8.58<br>[6.17,<br>9.64] | 1.59<br>[1.27,<br>1.64] | 1.55<br>[1.21,<br>1.61] | 9.38<br>[8.14,<br>10.69] |
| **lung_cancer** | 2 | 5 | 1.56%<br>[0.62%,<br>1.74%] | 3.65<br>[2.93,<br>9.67] | 1.33<br>[1.29,<br>1.38] | 1.60<br>[1.58,<br>1.83] | 9.50<br>[5.10,<br>10.17] |
| **lung_cancer** | 5 | 10 | 1.32%<br>[1.00%,<br>2.41%] | 2.76<br>[2.46,<br>4.41] | 1.25<br>[1.09,<br>1.37] | 1.44<br>[1.24,<br>1.59] | 5.39<br>[3.79,<br>6.74] |
| **myocardial_infarction** | 1 | 9 | 1.14%<br>[0.87%,<br>1.60%] | 2.31<br>[2.10,<br>2.40] | 1.28<br>[1.10,<br>1.66] | 2.04<br>[1.43,<br>2.37] | 4.89<br>[4.04,<br>5.85] |
| **myocardial_infarction** | 2 | 13 | 1.78%<br>[1.40%,<br>2.51%] | 2.30<br>[2.01,<br>2.37] | 1.08<br>[1.02,<br>1.36] | 1.53<br>[1.45,<br>1.95] | 4.47<br>[3.57,<br>5.60] |
| **myocardial_infarction** | 5 | 15 | 3.72%<br>[2.81%,<br>4.58%] | 2.04<br>[1.62,<br>2.28] | 1.15<br>[1.03,<br>1.25] | 1.60<br>[1.43,<br>1.95] | 3.70<br>[2.88,<br>4.36] |

Support was substantial: the median number of A→B episodes contributing to each validated AB pair was 1825 (IQR 1315–2755; range 535–6845). Reverse-order comparators (B→A) satisfied information gates for 11 of 12 sequences at the 1-year horizon, 20 of 22 sequences at the 2-year horizon, and 30 of 34 sequences at the 5-year horizon, yielding a median A→B→C vs B→A→C RR of 1.21 (IQR 1.02–1.41); in 64% of sequence-horizon combinations where the reverse order comparator could be calculated (39/61) the A→B→C vs B→A→C risk ratio was greater than 1.1. This heterogeneity across order-reversal checks motivated case-level inspection (below), while the primary inference (sequence vs B without prior A) remained consistently elevated among validated pairs. 36 hypotheses had risk ratios above 1.1 for the sequence versus all four comparators.

Figure 1, shown below, illustrates the distribution of cumulative incidence factors and risk ratios versus comparators.

**Figure 1.**
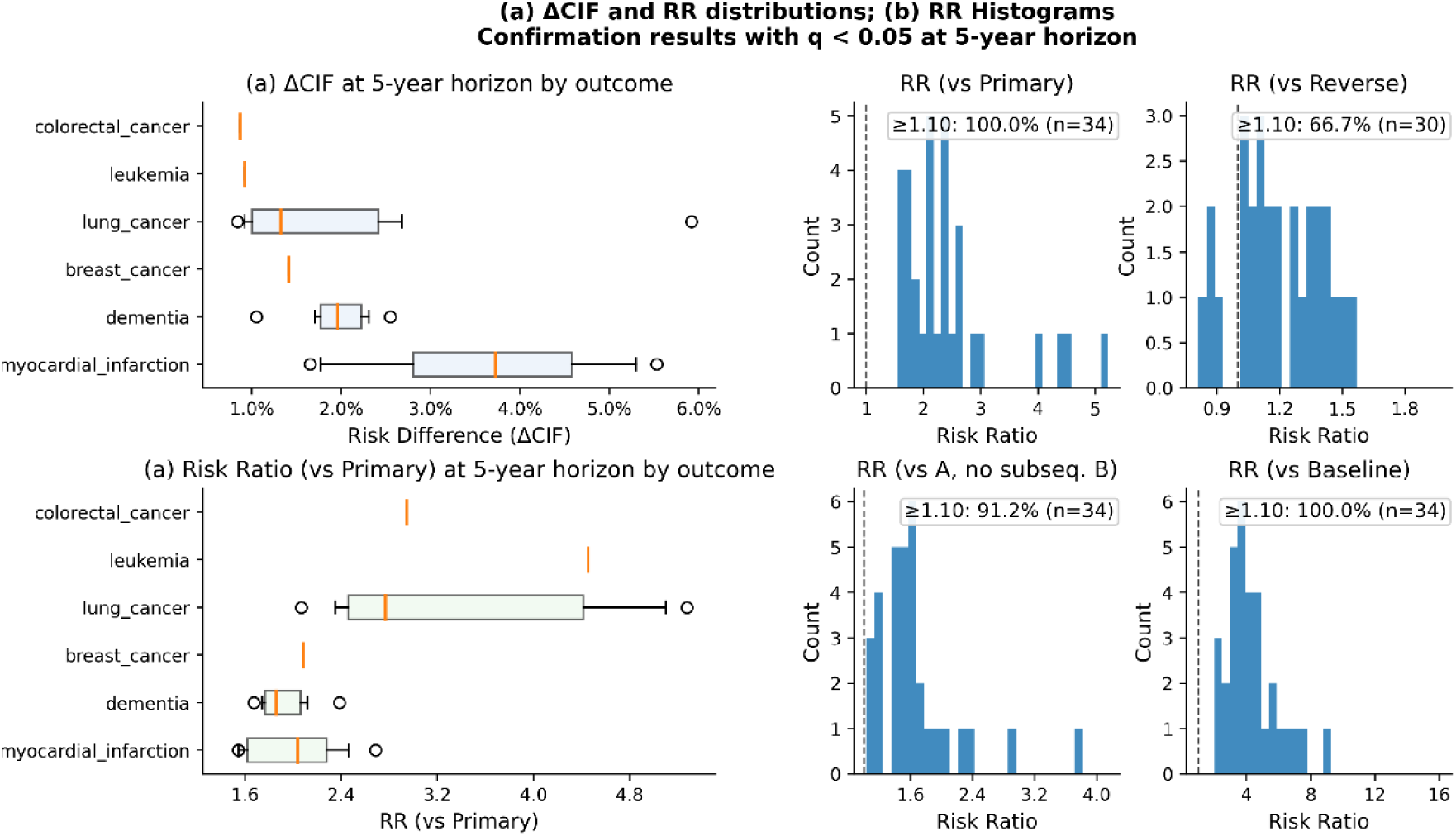
Distribution of effect sizes across validated A→B→C trajectories. The figure summarizes the spread of cumulative incidence and risk ratios for validated sequences versus their comparators across outcomes and follow-up. Medians and interquartile ranges are indicated; higher values reflect larger excess risk relative to comparators. (RR, risk ratio.)

Inverse probability of treatment weighting (IPTW) achieved covariate balance between A→B and comparators across age, sex at birth, race/ethnicity, calendar time, smoking, BMI, deprivation index, healthcare utilization, and CCI-LOO; overlap diagnostics were acceptable.

Figure 2, below, shows love plots of two illustrative disease trajectory sequences.

**Figure 2.**
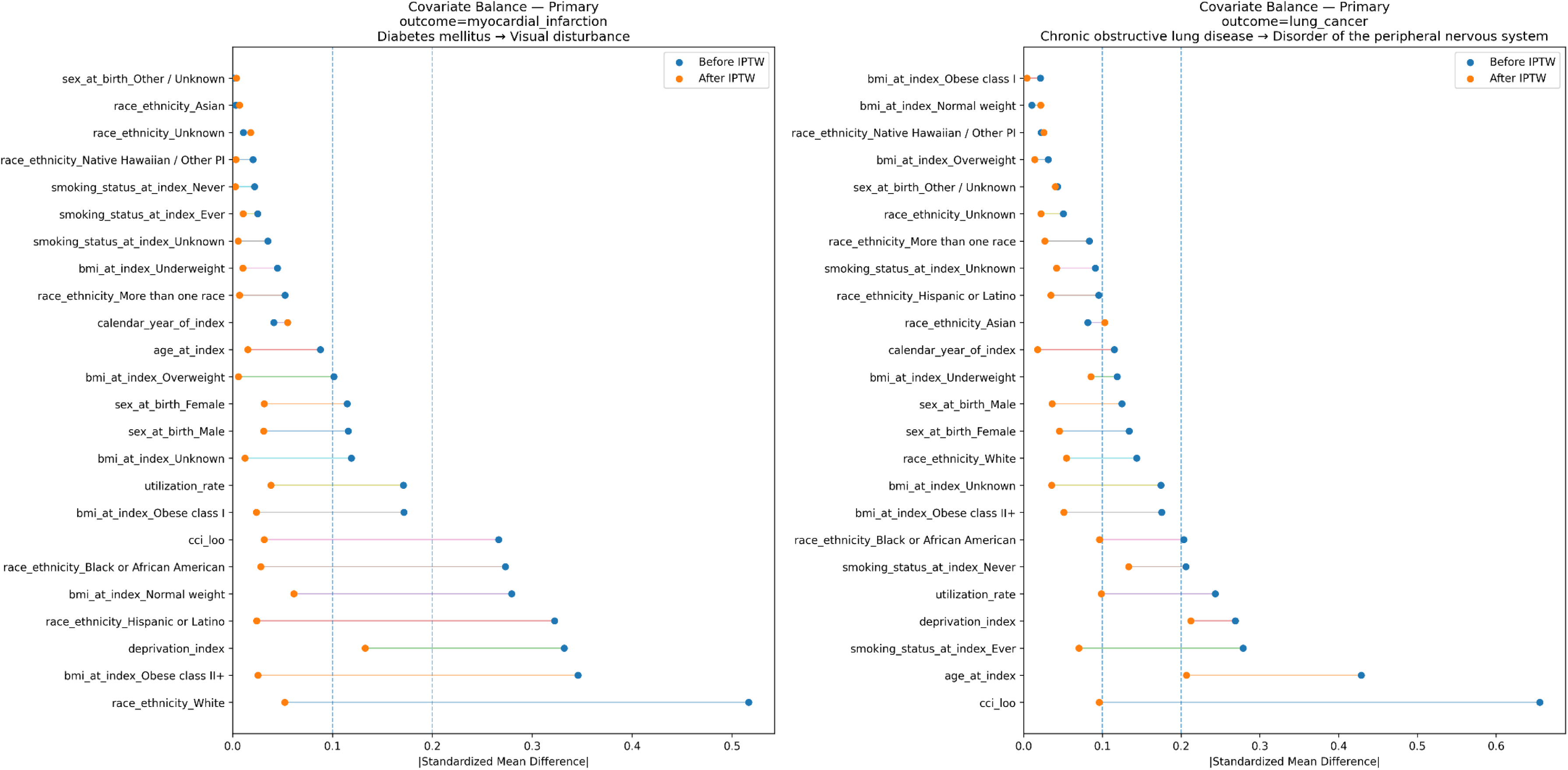
Covariate balance (“love”) plots for two illustrative sequences. (A) Acquired absence of organ → obstructive sleep apnea (OSA)→ dementia. (B) Diabetes mellitus → visual disturbance → myocardial infarction (MI). Points show standardized mean differences (SMD) for baseline covariates before and after inverse probability of treatment weighting (IPTW); values closer to 0 indicate improved balance. (IPTW, inverse probability of treatment weighting; SMD, standardized mean difference.)

### Comparator consistency and robustness checks

Across validated sequences, results were consistent against tertiary comparators (A without subsequent B) and a calendar-time–anchored baseline, with directionally concordant and attenuated effects as expected.

Pre-specified sensitivity analyses varying latency (30, 90, 180 days) and the maximum allowable A→B gap (1, 5, 10 years) yielded qualitatively similar estimates for top signals.

Validated signals clustered around clinically coherent pathways: cardiometabolic sequences (e.g., diabetes/obesity/angina) tending to lead to MI; pulmonary and systemic-inflammation sequences (COPD or tobacco-related exposures) tending towards lung cancer; and sleep/neurologic pathways tending towards dementia.

**Table 3.** 25 significant validated hypotheses at the 5-year time horizon and their risk ratios versus all comparators.

| Sequence (A→B) | Outcome (C) | Horizon<br>(Years) | Risk Ratio<br>(vs<br>Baseline) | Risk<br>Ratio (vs<br>Primary) | Risk Ratio<br>(vs Reverse) | Risk<br>Ratio (vs<br>A, no<br>subseq.<br>B) | q-value<br>(Confirmation) |
| --- | --- | --- | --- | --- | --- | --- | --- |
| Diabetes mellitus<br>→ Congestive<br>heart failure | Myocardial<br>Infarction | 5 | 7.6863 | 1.6235 | 1.5154 | 3.8178 | <1E-4 |
| Malignant neoplastic disease<br>→ Disorder of the peripheral nervous system | Lung Cancer | 5 | 6.8826 | 5.0998 | 1.3731 | 1.5773 | <1E-4 |
| insulin lispro → Disorder of lumbar spine | Dementia | 5 | 6.5472 | 2.1186 | 1.4052 | 1.8898 | 0.0365 |
| Malignant neoplastic disease<br>→ Inflammatory disorder of digestive tract | Lung Cancer | 5 | 6.3067 | 5.2761 | 1.057 | 1.4702 | <1E-4 |
| Atherosclerosis of coronary artery without angina pectoris → Aortic valve disorder | Myocardial Infarction | 5 | 5.8381 | 1.7141 | 1.0231 | 1.4303 | 0.0365 |
| Chronic obstructive lung disease → Disorder of the peripheral nervous system | Lung Cancer | 5 | 5.6241 | 2.8877 | 1.1012 | 1.1183 | <1E-4 |
| Chronic lung disease → Edema | Lung Cancer | 5 | 5.1463 | 2.6389 | 1.153 | 1.0852 | <1E-4 |
| Disorder of brain<br>→ Hypo-<br>osmolality and or<br>hyponatremia | Dementia | 5 | 4.7549 | 1.674 | 1.0809 | 1.5267 | 0.0365 |
| Hypertensive<br>disorder →<br>Angina pectoris | Myocardial<br>Infarction | 5 | 4.6816 | 2.0937 | 1.1676 | 2.9583 | <1E-4 |
| Acquired absence<br>of organ →<br>Obstructive sleep<br>apnea syndrome | Dementia | 5 | 4.5851 | 1.8335 | 1.4459 | 1.5296 | <1E-4 |
| Disorder due to<br>type 2 diabetes<br>mellitus →<br>Obesity | Myocardial<br>Infarction | 5 | 4.4575 | 2.6845 | 1.209 | 1.5965 | <1E-4 |
| Malignant<br>neoplastic disease<br>→ Inflammatory<br>disorder of<br>digestive tract | Leukemia | 5 | 4.0429 | 4.4518 | 1.4146 | 1.5674 | <1E-4 |
| Disorder of<br>hematopoietic<br>structure →<br>Venous varices | Lung Cancer | 5 | 3.988 | 2.6408 | 1.3483 | 1.2002 | 0.0365 |
| Diabetes mellitus<br>→ Visual<br>disturbance | Myocardial<br>Infarction | 5 | 3.932 | 2.4608 | 1.5562 | 1.8482 | <1E-4 |
| Syncope and<br>collapse →<br>Anxiety | Dementia | 5 | 3.9135 | 1.8823 | 1.2778 | 2.266 | 0.0365 |
| heparin →<br>Inflammatory<br>disorder of<br>digestive tract | Lung Cancer | 5 | 3.7187 | 2.3997 | 1.0146 | 1.4121 | <1E-4 |
| Diabetes mellitus<br>→ Blood glucose<br>abnormal | Myocardial<br>Infarction | 5 | 3.695 | 2.3256 | 1.4709 | 2.0496 | <1E-4 |
| Disorder of<br>endocrine system<br>→ glucagon | Myocardial<br>Infarction | 5 | 3.6851 | 1.7615 | 1.12 | 2.3633 | <1E-4 |
| Type 2 diabetes<br>mellitus →<br>Gastroesophageal<br>reflux disease<br>without<br>esophagitis | Myocardial<br>Infarction | 5 | 3.4243 | 2.1259 | 1.2037 | 1.7075 | <1E-4 |
| nicotine →<br>Depressive<br>disorder | Myocardial<br>Infarction | 5 | 3.2542 | 2.2371 | 1.2684 | 1.5809 | <1E-4 |
| epinephrine →<br>Disorder of<br>esophagus | Lung Cancer | 5 | 3.1937 | 2.0672 | 1.3638 | 1.6166 | <1E-4 |
| Anxiety disorder<br>→ Disorder of<br>kidney and/or<br>ureter | Dementia | 5 | 3.0726 | 2.3838 | 1.3128 | 1.6425 | <1E-4 |
| Chronic pain →<br>duloxetine | Myocardial<br>Infarction | 5 | 2.5087 | 1.5458 | 1.1277 | 1.4367 | <1E-4 |
| Hypertensive<br>disorder →<br>Cataract | Myocardial<br>Infarction | 5 | 2.1238 | 2.3961 | 1.0458 | 1.1819 | <1E-4 |
| Noninflammatory<br>disorder of the<br>female genital<br>organs →<br>Disorder of brain | Breast<br>Cancer | 5 | 1.9377 | 2.0816 | 2.1338 | 1.4161 | <1E-4 |

A table of all confirmation-tested trajectory–horizon results (with CIFs, ΔCIFs, Risk Ratios (RR), and Poisson-bootstrap 95% CIs) accompanies the manuscript in the Supplementary Materials.

## Discussion

### Disease trajectories provide valuable insights

Our central finding is that the temporal order in which common clinical events unfold carries clinically meaningful, additional information about downstream risk. This information would be largely invisible to “bag-of-codes” featurizations that ignore sequence. Across six prespecified endpoints, we validated 39 distinct A→ B→ C sequences, with effects that were non-trivial in magnitude and consistent across multiple comparators and sensitivity analyses. At 5 years, the median absolute risk difference between an ordered sequence and the primary comparator (“(B with no prior A) → C”) had a median RR of 2.18 (IQR 1.76–2.65; maximum 5.28. These effect sizes are large enough to be relevant to clinical decision making in settings where baseline risks are modest and interventions carry cost or harm.

The trajectories generated by the pipeline generally appear to be biologically plausible. Below, we investigate four trajectories uncovered by the generator, including a potential mechanistic analysis and literature anchoring.

### Illustrative trajectories: clinical coherence and literature anchoring

#### COPD → peripheral nervous system disorder → lung cancer

Individuals who developed a peripheral nervous system (PNS) disorder after chronic obstructive pulmonary disease (COPD) had higher subsequent lung-cancer risk than those with the same “B” event but no prior COPD: ΔCIF 0.4pp (95% CI 0.1–0.9) at 1 year, 0.3pp (–0.1–0.8) at 2 years, and 1.4pp (0.7–2.4) at 5 years; the 5-year RR was 2.89, with A→B exceeding B→A at 5 years (RR 1.10). This pattern aligns with extensive evidence that COPD independently elevates lung-cancer risk beyond smoking, plausibly via a chronic, protumor inflammatory milieu that promotes oxidative DNA damage and abnormal epithelial repair. Reviews synthesize epidemiology showing markedly increased lung-cancer risk in COPD (even after adjustment for smoking) and delineate shared pathogenic pathways (persistent airway inflammation, immune dysregulation, and defective epithelial repair) that plausibly drive oncogenesis in COPD patients.[24, 25]

Moreover, several well-characterized PNS entities associated with lung cancer frequently precede tumor detection, supporting the interpretation that neuropathic symptoms can emerge along the cancer development and detection pathway. In a large anti-Hu (ANNA-1) cohort (n=466), neurological symptoms preceded cancer diagnosis in 84% of patients; when cancer was found, 89% were predominantly small-cell lung cancer (SCLC). Separately, Lambert–Eaton myasthenic syndrome is paraneoplastic in ∼40–62% of cases, overwhelmingly linked to SCLC, and the neuromuscular symptoms typically antedate cancer diagnosis, patterns that make a COPD → PNS disorder → lung cancer trajectory biologically coherent.[26, 27]

#### Chronic pain → duloxetine → myocardial infarction

Relative to incident duloxetine without prior chronic-pain coding, those with chronic pain preceding duloxetine showed higher myocardial infarction (MI) risk at 1, 2, and 5 years (ΔCIF ∼0.6, 1.2, 1.8pp; RRs ∼2.5, 2.2, 1.6). While residual confounding by indication is possible, the biological pathway is credible: chronic pain states exhibit sustained sympathetic overactivity (↑HR/BP, endothelial stress, pro-thrombotic milieu), and large cohorts link chronic or multisite pain to excess MI, stroke, heart failure, and cardiovascular mortality independent of traditional risk factors. This places patients with chronic pain on a higher baseline risk trajectory before any drug exposure.[28, 29]

Superimposing duloxetine’s serotonin-norepinephrine reuptake inhibitor pharmacology can plausibly amplify that risk in susceptible patients: meta-analysis shows duloxetine modestly raises heart rate (∼2 bpm) and diastolic BP, and safety catalogs cardiovascular adverse events, including myocardial ischemia, during treatment. Mechanistically, added noradrenergic tone can worsen supply-demand balance and provoke ischemia in individuals already primed by pain-related sympathetic activation.[30, 31] The combination of these factors indicates a plausible biological pathway for the ordered sequence of chronic pain, duloxetine, and myocardial infarction.

#### Acquired absence of organ → obstructive sleep apnea → dementia

Across this cohort, a coded acquired absence of organ followed by incident obstructive sleep apnea (OSA) carried higher 5-year dementia risk than OSA without prior organ absence (ΔCIF 1.9pp [0.5–3.6]; 5-year RR 1.83 vs primary event) and exceeded the reverse ordering (A→B > B→A; RR 1.45). This sequence is clinically coherent, with the most straightforward mechanism pointing towards patients who have undergone bilateral oophorectomy. Removal of the ovaries can trigger abrupt estrogen/progesterone loss, weight gain, and ventilatory/upper-airway changes that heighten subsequent OSA risk; in two large prospective cohorts, surgical (vs natural) menopause independently increased incident OSA by ∼25%. Premenopausal oophorectomy is also associated with higher long-term risk of cognitive impairment/dementia, stacking risk even before OSA is considered.[32, 33]

Downstream, OSA is consistently linked to cognitive decline and dementia via intermittent hypoxia and sleep fragmentation, with meta-analyses reporting ∼25–50% higher risk of cognitive impairment/dementia among people with OSA. Converging biomarker/imaging data show AD-type pathology in OSA (lower CSF Aβ42, higher tau, and greater brain amyloid burden on PET) supporting accelerated amyloid deposition as a mechanism.[34–37] Together, these anchors make the temporally ordered trajectory (acquired organ absence → OSA → dementia) more potentially riskier than either condition alone.

#### Diabetes mellitus → visual disturbance → myocardial infarction

Diabetes followed by a coded visual disturbance showed a large and graded association with MI versus incident visual disturbance without prior diabetes: ΔCIF 1.5 pp (0.8–2.2) at 1 year, 2.6 pp (1.5–3.5) at 2 years, and 4.5 pp (3.1–6.0) at 5 years (RRs 4.56, 3.77, and 2.46; A→B > B→A across horizons). This pattern is in consonance with literature linking diabetic retinopathy to future cardiovascular events and mortality, consistent with widespread microvascular disease and shared pathobiology between micro- and macrovascular complications.[38, 39]

Figure 3, shown below, shows the cumulative incidence of the outcome for each of the above disease trajectories compared to the baseline cohort.

**Figure 3.**
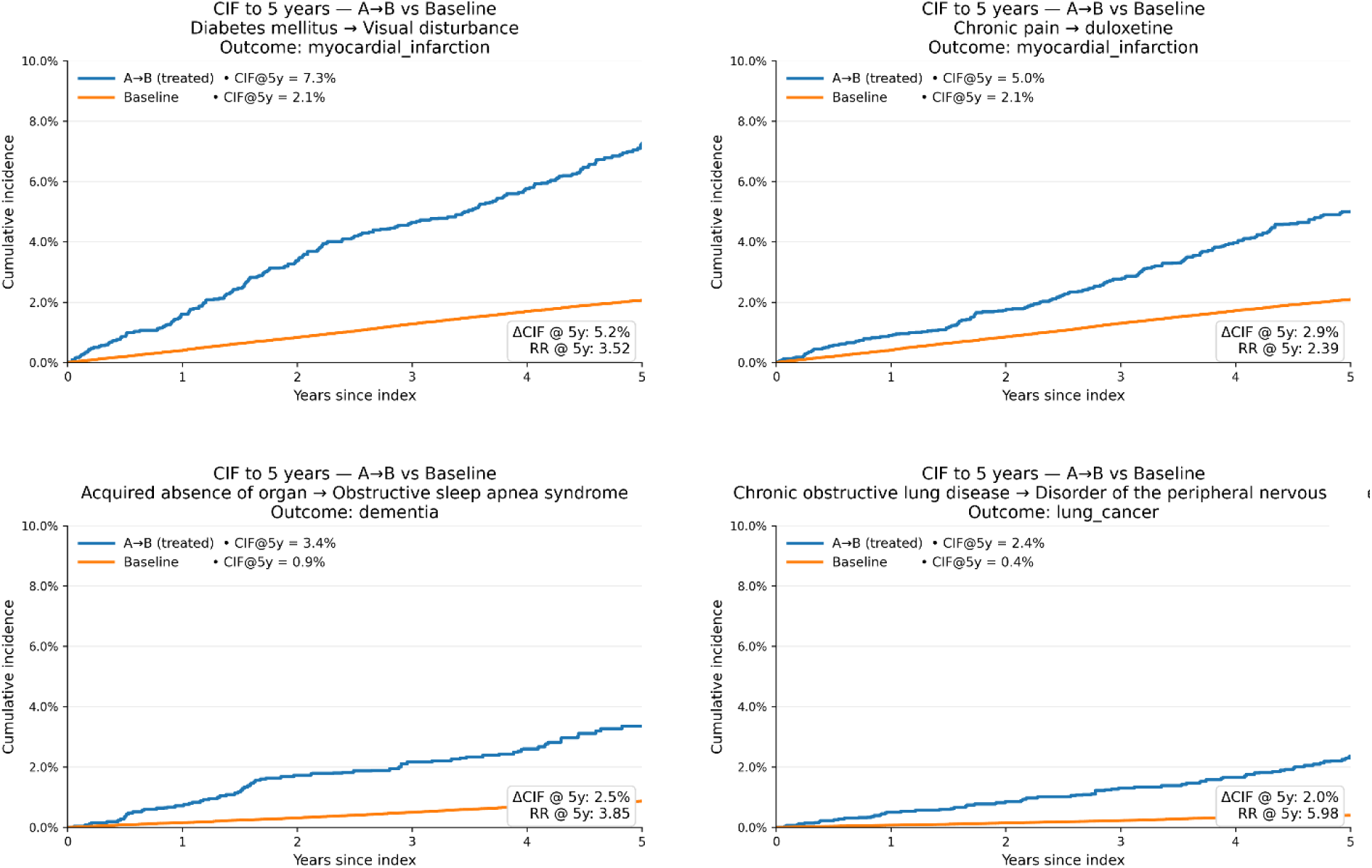
Cumulative incidence curves for the illustrative sequences versus the baseline comparator. Curves display estimated cumulative incidence functions over follow-up. Greater separation indicates larger absolute risk differences between the sequence cohort and its comparator. (CIF, cumulative incidence function; CI, confidence interval.)

### Methodological contributions

Beyond the specific signals, the study contributes a reproducible pipeline for trajectory discovery and validation in a large, diverse cohort. The frequent-pattern mining with support thresholds ensures hypotheses are data-anchored and powered; the discovery→confirmation split with global FDR control limits false positives; and the same-day policies, gap limits, and observation- period–aware censoring provide a principled template other systems can adopt. Importantly, effects were generally attenuated but directionally concordant against other comparators (A-no- B; baseline; reverse), reinforcing that the primary A→B vs B-no-A contrast is not an artifact of a single reference group.

### Limitations

These are observational analyses; residual confounding and phenotype misclassification remain possible. Our AoU-compliant dissemination rules (suppression for small cells; rounding) appropriately constrain granularity but can blur very fine-grained subgroup effects. Episodes rely on coded events; under-ascertainment of symptoms, measurement timing, and care-seeking behavior can shift apparent order. Finally, we restricted ourselves to ordered pairs; some pathways may require higher-order or time-gap–sensitive patterns (A→B→C→D) to fully capture risk propagation.

### Future directions

Two directions are especially promising. First, escalating to higher-order trajectories and variable gap/latency models should uncover deeper structure while preserving interpretability. We then envision assembling the validated sequences into a patient-specific “disease tree”—a directed graph in which early, modifiable events feed branches that culminate in adverse outcomes. Prevention then becomes pruning: intervening on a proximal antecedent within the empirically derived latency window (via treatment, screening, or behavior change) should attenuate risk along downstream branches.

Second, embedding trajectory signals into prospective workflows (e.g., screening eligibility checks, shared-decision aids, post-visit summaries) can translate population-scale patterns into timely, patient-level actions. Because our procedure yields calibrated, horizon-specific absolute incidence, clinicians can use trajectories discovered using this method to make informed risk assessments based on robust quantitative data for individual patients.

## Conclusion

In this retrospective study of 432,617 eligible participants in the All of Us Research Program, we show that temporally ordered clinical sequences mined from routine EHRs capture prognostic information that is largely invisible to unordered “bag-of-codes” representations. From 340,687 frequent A→B pairs discovered under pre-specified support thresholds, we form 3,066,183 A→B→C trajectories ending in adverse health events. Due to compute constraints, we were able to test 20,565 of these trajectories for risk elevation of adverse event C, and find 39 distinct A→B→C trajectories that validated under a separate confirmation analysis with global FDR control, spanning six clinically important endpoints.

Effect sizes were clinically meaningful: at 5 years, the median risk ratio was 2.18 (IQR 1.76–2.65) versus the primary comparator with a median absolute risk difference (“(B without prior A)→C”) of 2.3 percentage-points (IQR 1.4–3.7). Estimates were robust across reverse-order and tertiary comparators, achieved acceptable covariate balance with IPTW, and were consistent across sensitivity analyses varying latency and allowable gaps. Together, these results indicate that the order in which common conditions and exposures occur matters for downstream risk, not merely their presence.

Because the approach is transparent by construction (explicit sequences, observation-period– aware indexing, prespecified covariates, and competing-risk estimation) it complements existing risk calculators and end-to-end sequence models while remaining amenable to clinical audit. The resulting horizon-specific absolute risks can support practical decisions such as targeted screening, medication safety checks, and shared decision-making for patients who traverse high-risk trajectories. While observational limitations and phenotype misclassification remain relevant in this type of analysis, our discovery→confirmation framework, dissemination safeguards, and machine-readable results provide a reproducible template for trajectory-aware risk assessment. Future work should prospectively evaluate clinical utility, extend beyond ordered pairs to higher-order and time-gap–sensitive patterns, and embed these signals into workflows that translate population-scale trajectories into timely, patient-level action.

## Data Availability

All data, results, and code used are available in the supplementary materials.

## Author Contributions

Jeffrey Skolnick originated the idea of studying time dependent disease trajectories to infer a causal sequence of disease events. Brice Edelman and Jeffrey Skolnick collaborated on ideation and design of the data analysis methodology. Brice Edelman wrote the code for the computational analysis and generation of results, tables, and figures. Brice Edelman and Hannah Kim, the clinician contributor, collaborated on the investigation of mechanistic plausibility of illustrative trajectories. Brice Edelman wrote the draft of the manuscript, and Brice Edelman and Jeffrey Skolnick collaborated on revisions.

## Data Availability

The full code used for the analysis, as well as the results table and sensitivity analysis results, are included in the Supplementary Data.

## Patient and Public Involvement

Patients or the public were not involved in the design, or conduct, or reporting, or dissemination plans of this research.

## Supplementary Data

We include the following artefacts in the “SupplementaryData.zip” archive:

- Results.xlsx

`o` The main results table, including the comparator CIFs and Risk Ratios for each trajectory-horizon combination.
- SensitivityAnalysis.xlsx

`o` The results of the sensitivity analysis testing different latency and max gap parameters run for the top validated signals.
- Code.py

`o` The code run in the secure All of Us workspace used to generate the results and sensitivity analysis.

## Acknowledgements

We gratefully acknowledge *All of Us* participants for their contributions, without whom this research would not have been possible. We also thank the National Institutes of Health’s *<u>All of Us</u>* <u>Research Program</u> for making available the participant data examined in this study. We thank Jessica Forness for assistance in preparation of the manuscript.

We leveraged OpenAI’s GPT-5-Pro AI Model for code debugging and final grammar revision of the manuscript, and take full responsibility for the content.

