## Supplementary for "Trajectories matter: Discovery and validation of ordered EHR sequences that inform clinical risk predictions": Code.py

```
#!/usr/bin/env python
# coding: utf-8

# In[ ]:


# ──────────────────────────── Cell 0 ─────────────────────────────────────────
# Global Imports, Environment Sanity Checks, and Client Initialization
# ---------------------------------------------------------------------------
import sys, subprocess
def _ensure_numpy_1x():
    import numpy as _np
    major = int(_np.__version__.split('.')[0])
    if major >= 2:
        print("⚠️  Detected NumPy 2.x; rolling back to 1.x for AoU binary compatibility…")
        subprocess.check_call(
            [sys.executable, "-m", "pip", "install", "numpy<2",
             "--force-reinstall", "--no-cache-dir"]
        )
        print("✅ NumPy downgraded. Please restart the kernel and re-run.")
        raise SystemExit
_ensure_numpy_1x()

# 0‑B ▸ Core & scientific stack ------------------------------------------------
import os, json, logging, random
from pathlib import Path
from datetime import datetime

import numpy as np
import pandas as pd
import matplotlib as mpl
import matplotlib.pyplot as plt
try:
    import pyarrow as pa, pyarrow.parquet as pq
    print("pyarrow available for writing Parquet metadata.")
except Exception:
    print("WARNING: pyarrow not available; will fall back to JSON sidecars for metadata.")

try:
    import lifelines
    from lifelines import AalenJohansenFitter  # noqa: F401 (used later)
except Exception as e:
    raise ImportError(
        "lifelines is required (including AalenJohansenFitter). "
        "Install with `pip install lifelines` inside the runtime."
    ) from e

try:
    from tqdm.auto import tqdm  # progress bars
except ImportError:
    tqdm = None

# 0‑C ▸ BigQuery client (AoU Workbench) --------------------------------------
from google.cloud import bigquery

WORKSPACE_CDR = os.getenv("WORKSPACE_CDR")
if not WORKSPACE_CDR:
    raise EnvironmentError(
        "`WORKSPACE_CDR` env var not set — launch inside an AoU Workbench runtime "
        "or manually `export WORKSPACE_CDR=project.dataset`."
    )

client = bigquery.Client()
print(f"✅ BigQuery client initialised — CDR = {WORKSPACE_CDR}")

# 0‑D ▸ RUN_ID and directory structure ---------------------------------------
RUN_ID = f"SET_RUN_ID_HERE"
BASE_DIR = Path.cwd()
RUN_DIR = BASE_DIR / RUN_ID
CONFIG_DIR   = RUN_DIR / "config"
LOG_DIR      = RUN_DIR / "logs"
ARTIFACTS_DIR = RUN_DIR / "artifacts"
RESULTS_DIR  = RUN_DIR / "results"
FIGURES_DIR  = RUN_DIR / "figures"

for d in (CONFIG_DIR, LOG_DIR, ARTIFACTS_DIR, RESULTS_DIR, FIGURES_DIR):
    d.mkdir(parents=True, exist_ok=True)

# 0‑E ▸ Logging to file + stdout ---------------------------------------------
logging.basicConfig(
    level=logging.INFO,
    format="%(asctime)s │ %(levelname)-8s │ %(message)s",
    handlers=[
        logging.FileHandler(LOG_DIR / "pipeline.log", encoding="utf-8"),
        logging.StreamHandler(sys.stdout),
    ],
)
log = logging.getLogger("risk-pipeline").info

# 0‑F ▸ Deterministic seeds (documented for reproducibility) ------------------
RNG_SEED = 42
random.seed(RNG_SEED)
np.random.seed(RNG_SEED)
os.environ["PYTHONHASHSEED"] = str(RNG_SEED)

# 0‑G ▸ Lightweight provenance logging
def _git_rev():
    try:
        return subprocess.check_output(
            ["git", "rev-parse", "--short", "HEAD"],
            stderr=subprocess.DEVNULL
        ).decode().strip()
    except Exception:
        return None

log(f"Cell 0 ready ✔️  RUN_ID={RUN_ID}  RNG_SEED={RNG_SEED}")
log(f"CDR={WORKSPACE_CDR}  Python={sys.version.split()[0]}  "
    f"numpy={np.__version__}  pandas={pd.__version__}  "
    f"lifelines={getattr(lifelines, '__version__', 'unknown')}  "
    f"matplotlib={mpl.__version__}  google-cloud-bigquery={getattr(bigquery, '__version__', 'unknown')}")
_git = _git_rev()
if _git:
    log(f"Git commit={_git}")


# In[ ]:


# ──────────────────────────── Cell 1 (REVISED) ─────────────────────────────
# Master Configuration and Provenance Snapshot
# ---------------------------------------------------------------------------

from dataclasses import dataclass, field, asdict
from pathlib import Path
import json

# Helper class to serialize Path objects to strings in JSON
class PathEncoder(json.JSONEncoder):
    def default(self, obj):
        if isinstance(obj, Path):
            return str(obj)
        return super().default(obj)

@dataclass(frozen=True)
class AnalysisConfig:
    """Immutable dataclass holding all parameters for the analysis run."""
    # --- Run & Provenance ---------------------------------------------------
    RUN_ID: str = RUN_ID
    RNG_SEED: int = RNG_SEED
    CDR_VERSION: str = WORKSPACE_CDR.split('.')[-1]

    # --- Data Splits --------------------------------------------------------
    DISCOVERY_FRAC: float = 0.70

    # --- Paths (derived from RUN_ID in Cell 0) ------------------------------
    SOURCE_CACHE_DIR: Path = BASE_DIR / "analysis_source_files"
    CURATED_FEATURES_CSV_PATH: Path = SOURCE_CACHE_DIR / "manually_curated_features.csv"

    # Paths for this analysis run's outputs
    CONFIG_DIR: Path = CONFIG_DIR
    LOG_DIR: Path = LOG_DIR
    ARTIFACTS_DIR: Path = ARTIFACTS_DIR
    RESULTS_DIR: Path = RESULTS_DIR
    FIGURES_DIR: Path = FIGURES_DIR

    # --- Phenotyping & Labeling --------------------------------------------
    OUTCOME_DEFINITIONS: dict = field(default_factory=lambda: {
        "breast_cancer":         [4112853, 81250],
        "ovarian_cancer":        [4181351],
        "colorectal_cancer":     [36683531],
        "lung_cancer":           [443388],
        "leukemia":              [317510],
        "lymphoma":              [432571],
        "myocardial_infarction": [4329847],
        "dementia":              [4182210],
        "death": []
    })

    # --- Episode Construction Logic -----------------------------------------
    EPISODE_MAX_GAP_DAYS: int = 1825  # 5 years
    SAME_DAY_POLICY: str = 'exclude'  # Options: 'allow' | 'exclude' | 'require'

    # --- Time Windows & Horizons --------------------------------------------
    RISK_HORIZONS_YEARS: tuple = (1, 2, 5)
    LATENCY_DAYS: int = 90
    RISKSET_MIN_LOOKBACK_DAYS: int = 1825

    # --- Statistical & Mining Thresholds ------------------------------------
    MIN_STRATUM_SUPPORT: int = 500   # Min persons in a demographic stratum to be considered
    MIN_SEQUENCE_SUPPORT: int = 500  # Min persons in a stratum to define a sequence cohort
    MIN_CASES_FOR_CIF: int = 20      # Min outcome events to run a CIF fit
    MIN_ATRISK_AT_H: int = 150       # Min persons at risk at a horizon for stable estimate
    FDR_ALPHA: float = 0.05          # False discovery rate for candidate selection

    # --- Modeling & Adjustment ----------------------------------------------
    COVARIATES_FOR_ADJUSTMENT: list = field(default_factory=lambda: [
        'age_at_index',
        'sex_at_birth',
        'race_ethnicity',
        'calendar_year_of_index',
        'smoking_status_at_index',
        'bmi_at_index',
        'deprivation_index',
        'utilization_rate', 
        'cci_loo'
    ])

    # --- AoU Dissemination Compliance ---------------------------------------
    MIN_CELL_SIZE: int = 20
    ROUND_TO_NEAREST: int = 5

    EPISODE_FILE_PATTERN = "seq_{seq_idx}__oc_{outcome}.parquet"
    SMD_SIDECAR_PATTERN  = "smd_{seq_idx}__oc_{outcome}.json"

# --- Instantiate the configuration ------------------------------------------
CONFIG = AnalysisConfig()

# --- Save a snapshot for provenance -----------------------------------------
config_snapshot_path = CONFIG.CONFIG_DIR / "config_snapshot.json"
with open(config_snapshot_path, "w", encoding="utf-8") as f:
    json.dump(asdict(CONFIG), f, indent=2, cls=PathEncoder)

log(f"Master configuration loaded and frozen for RUN_ID: {CONFIG.RUN_ID}")
log(f"Configuration snapshot saved to: {config_snapshot_path}")
log("Cell 1 complete ✔️ (Master configuration defined and saved)")


# In[ ]:


# ──────────────────────────── Cell 2 (Re-Revised, SES Fix) ─────────────────────────────
# Load and Prepare Master Person & Observation Interval Tables
# ---------------------------------------------------------------------------------------
# Outputs (cached):
#   artifacts/persons_df.parquet
#   artifacts/observation_periods_df.parquet

import os
from pathlib import Path
from typing import Optional

import numpy as np
import pandas as pd

from google.cloud import bigquery

try:
    from IPython.display import display  # pretty heads in notebook
except Exception:
    def display(x):  # noqa: E306
        print(x.head() if hasattr(x, "head") else x)

print("▶ Cell 2: Building master person & observation-period tables...")

# ---------------------------------------------------------------------------------------
# Paths & simple knobs
# ---------------------------------------------------------------------------------------
persons_out_path = CONFIG.ARTIFACTS_DIR / "persons_df.parquet"
obs_periods_out_path = CONFIG.ARTIFACTS_DIR / "observation_periods_df.parquet"
ses_metrics_path = CONFIG.ARTIFACTS_DIR / "ses_join_metrics.json"

FORCE_REBUILD = os.getenv("FORCE_REBUILD_CELL2", "0").strip().lower() in {"1", "true", "yes"}
SES_TABLE_FQN = os.getenv("SES_TABLE_FQN", f"{WORKSPACE_CDR}.zip3_ses_map")
ELIG_MERGE_GAP_DAYS = 1      # merge obs. intervals that abut/overlap by <=1 day
OBS_CHUNK = 80000            # chunk size for pulling intervals by eligible_pids

# ---------------------------------------------------------------------------------------
# Utilities
# ---------------------------------------------------------------------------------------
def _norm_zip3_series(s: pd.Series) -> pd.Series:
    """
    Normalize any ZIP-like values to a 3-digit zero-padded string.

    Steps:
      - Cast to string
      - Strip non-digits
      - Take the first 3 digits
      - Left-pad to 3 chars
      - Return NaN if result is not exactly 3 digits
    """
    ss = s.astype("string")
    digits = ss.str.replace(r"\D+", "", regex=True).str.slice(0, 3)
    z3 = digits.str.zfill(3)
    z3 = z3.where(z3.str.match(r"^\d{3}$", na=False))
    return z3.astype("object")

def _bq_df(query: str, job_config: Optional[bigquery.QueryJobConfig] = None) -> pd.DataFrame:
    qjob = client.query(query, job_config=job_config)
    return qjob.result().to_dataframe(create_bqstorage_client=False)

# ---------------------------------------------------------------------------------------
# Cache check: load if present and not forced to rebuild; sanity-check SES column
# ---------------------------------------------------------------------------------------
_cache_ok = persons_out_path.exists() and obs_periods_out_path.exists()
if _cache_ok and not FORCE_REBUILD:
    persons_df = pd.read_parquet(persons_out_path)
    observation_periods_df = pd.read_parquet(obs_periods_out_path)

    # If SES exists and looks healthy, keep cache; otherwise force rebuild.
    ses_ok = False
    if "deprivation_index" in persons_df.columns:
        di = persons_df["deprivation_index"]
        # "Healthy" if there is at least some variation and not all missing/constant
        unique_non_null = di.dropna().nunique()
        ses_ok = (di.notna().any() and unique_non_null > 5)
    if ses_ok:
        print(f"  • Loaded cached persons_df:             {persons_out_path.name}")
        print(f"  • Loaded cached observation_periods_df: {obs_periods_out_path.name}")
        print(f"  • persons_df rows: {len(persons_df):,} | observation_periods_df rows: {len(observation_periods_df):,}")
        print("✅ Cell 2 complete (loaded from cache).")
        display(persons_df.head())
        display(observation_periods_df.head())
    else:
        print("  • Cache present but SES looks missing/constant; rebuilding artifacts for corrected SES...")
        _cache_ok = False

if _cache_ok and not FORCE_REBUILD:
    # Exit early if cache is accepted
    pass
else:
    print("  • Building artifacts from BigQuery…")

    # -----------------------------------------------------------------------------------
    # 0) SES mapping table (ZIP3 → deprivation_index), robust normalization
    # -----------------------------------------------------------------------------------
    print(f"    - Loading SES mapping from: {SES_TABLE_FQN}")

    # Detect candidate columns from INFORMATION_SCHEMA to be resilient to schema variants
    schema_df = _bq_df(
        f"""
        SELECT column_name, data_type
        FROM `{WORKSPACE_CDR}.INFORMATION_SCHEMA.COLUMNS`
        WHERE table_name = '{SES_TABLE_FQN.split('.')[-1]}'
        """
    )
    schema_lower = {r["column_name"].lower(): r["column_name"] for _, r in schema_df.iterrows()} if not schema_df.empty else {}

    # Prefer a string zip3 column if present; fallback to numeric zip3
    zip3_col = schema_lower.get("zip3_as_string") or schema_lower.get("zip3")
    dep_col = (
        schema_lower.get("deprivation_index")
        or schema_lower.get("adi")
        or schema_lower.get("area_deprivation_index")
        or "deprivation_index"
    )

    ses_df = pd.DataFrame(columns=["zip3_key", "deprivation_index"])
    _ses_status = "absent"
    try:
        # Normalize in SQL so we never lose leading zeros
        ses_sql = f"""
        SELECT
          CASE
            WHEN REGEXP_CONTAINS(CAST({zip3_col} AS STRING), r'^\\d+$')
              THEN LPAD(CAST({zip3_col} AS STRING), 3, '0')
            ELSE LPAD(REGEXP_EXTRACT(CAST({zip3_col} AS STRING), r'(\\d{{1,3}})'), 3, '0')
          END AS zip3_key,
          CAST({dep_col} AS FLOAT64) AS deprivation_index
        FROM `{SES_TABLE_FQN}`
        WHERE {dep_col} IS NOT NULL
        """
        ses_df = _bq_df(ses_sql)
        ses_df = ses_df.dropna(subset=["zip3_key", "deprivation_index"])
        # Deduplicate: if multiple rows per ZIP3, average deprivation_index
        ses_df = ses_df.groupby("zip3_key", as_index=False, dropna=False)["deprivation_index"].mean()
        _ses_status = f"mapped_from {SES_TABLE_FQN} (zip3='{zip3_col}', dep='{dep_col}')"
        print(f"    - SES map rows: {len(ses_df):,}; unique ZIP3s: {ses_df['zip3_key'].nunique():,}")
    except Exception as e:
        print(f"    - INFO: Could not load SES mapping table ({SES_TABLE_FQN}): {e}")
        _ses_status = "absent"

    # -----------------------------------------------------------------------------------
    # 1) Pull core demographics + ZIP3 (one row per person)
    # -----------------------------------------------------------------------------------
    print("    - Querying person + ds_zip_code_socioeconomic for demographics and ZIP3...")
    person_sql = f"""
    SELECT
      p.person_id,
      p.year_of_birth,
      p.sex_at_birth_concept_id,
      p.race_concept_id,
      p.ethnicity_concept_id,
      ses_zip.ZIP3_AS_STRING AS zip3
    FROM `{WORKSPACE_CDR}.person` p
    LEFT JOIN `{WORKSPACE_CDR}.ds_zip_code_socioeconomic` ses_zip
      ON p.person_id = ses_zip.person_id
    """
    person_raw_df = _bq_df(person_sql)
    if person_raw_df.empty:
        raise RuntimeError("Person query returned no rows. Check table permissions and names.")

    # -----------------------------------------------------------------------------------
    # 2) Eligibility via merged observation intervals (>=365 total days)
    # -----------------------------------------------------------------------------------
    print("    - Computing total observed days per person (merging overlaps)…")
    eligibility_sql = f"""
    WITH op AS (
      SELECT person_id,
             observation_period_start_date AS s,
             observation_period_end_date   AS e
      FROM `{WORKSPACE_CDR}.observation_period`
      WHERE observation_period_start_date IS NOT NULL
        AND observation_period_end_date   IS NOT NULL
    ),
    ordered AS (
      SELECT person_id, s, e,
             MAX(e) OVER (PARTITION BY person_id ORDER BY s, e
                          ROWS BETWEEN UNBOUNDED PRECEDING AND CURRENT ROW) AS running_end
      FROM op
    ),
    flagged AS (
      SELECT person_id, s, e, running_end,
             LAG(running_end) OVER (PARTITION BY person_id ORDER BY s, e) AS prev_running_end
      FROM ordered
    ),
    islands AS (
      SELECT person_id, s, e,
             CASE
               WHEN prev_running_end IS NULL THEN 1
               WHEN s > DATE_ADD(prev_running_end, INTERVAL @merge_gap_days DAY) THEN 1
               ELSE 0
             END AS new_island
      FROM flagged
    ),
    grouped AS (
      SELECT person_id, s, e,
             SUM(new_island) OVER (PARTITION BY person_id ORDER BY s, e
                                   ROWS BETWEEN UNBOUNDED PRECEDING AND CURRENT ROW) AS grp
      FROM islands
    ),
    merged AS (
      SELECT person_id,
             MIN(s) AS merged_start,
             MAX(e) AS merged_end
      FROM grouped
      GROUP BY person_id, grp
    ),
    totals AS (
      SELECT person_id,
             -- day-diff (exclusive). Use +1 for inclusive if desired.
             SUM(DATE_DIFF(merged_end, merged_start, DAY)) AS total_obs_days,
             MAX(merged_end) AS last_obs_end_date
      FROM merged
      GROUP BY person_id
    )
    SELECT person_id, total_obs_days, last_obs_end_date
    FROM totals
    WHERE total_obs_days >= 365
    """
    job_config = bigquery.QueryJobConfig(
        query_parameters=[bigquery.ScalarQueryParameter("merge_gap_days", "INT64", ELIG_MERGE_GAP_DAYS)]
    )
    eligible_df = _bq_df(eligibility_sql, job_config)
    if eligible_df.empty:
        raise RuntimeError("No eligible persons (>=365 total observed days) found in observation_period.")

    # -----------------------------------------------------------------------------------
    # 3) Retrieve ALL observation intervals for eligible persons (chunked)
    # -----------------------------------------------------------------------------------
    print("    - Pulling all observation intervals for the eligible cohort (chunked)…")
    eligible_pids = eligible_df["person_id"].astype(np.int64).tolist()

    frames = []
    total_chunks = (len(eligible_pids) + OBS_CHUNK - 1) // OBS_CHUNK
    for i in range(0, len(eligible_pids), OBS_CHUNK):
        chunk = eligible_pids[i : i + OBS_CHUNK]
        jc = bigquery.QueryJobConfig(
            query_parameters=[bigquery.ArrayQueryParameter("eligible_pids", "INT64", chunk)]
        )
        obs_intervals_sql = f"""
        SELECT
          person_id,
          observation_period_start_date AS obs_start_date,
          observation_period_end_date   AS obs_end_date,
          DATE_DIFF(observation_period_end_date, observation_period_start_date, DAY) AS interval_days
        FROM `{WORKSPACE_CDR}.observation_period`
        WHERE person_id IN UNNEST(@eligible_pids)
          AND observation_period_start_date IS NOT NULL
          AND observation_period_end_date   IS NOT NULL
        """
        frames.append(_bq_df(obs_intervals_sql, jc))
        if total_chunks > 1:
            print(f"      • chunk {(i // OBS_CHUNK) + 1}/{total_chunks}  rows={len(frames[-1]):,}")

    observation_periods_df = (
        pd.concat(frames, ignore_index=True) if frames else pd.DataFrame(columns=["person_id", "obs_start_date", "obs_end_date", "interval_days"])
    )
    if observation_periods_df.empty:
        raise RuntimeError("Eligible persons found, but no observation intervals retrieved.")

    # Normalize dtypes
    observation_periods_df["obs_start_date"] = pd.to_datetime(observation_periods_df["obs_start_date"])
    observation_periods_df["obs_end_date"] = pd.to_datetime(observation_periods_df["obs_end_date"])
    observation_periods_df["interval_days"] = pd.to_numeric(observation_periods_df["interval_days"], errors="coerce").astype("Int64")

    # -----------------------------------------------------------------------------------
    # 4) Build the final persons_df (one row per person) + SES
    # -----------------------------------------------------------------------------------
    print("    - Assembling one-row-per-person table with demographics, derived fields, and SES...")

    # Merge core demographics with eligibility stats
    persons_df = pd.merge(person_raw_df, eligible_df, on="person_id", how="inner")

    # Map concept IDs to readable strings
    SEX_MAP = {45880669: "Male", 45878463: "Female"}
    RACE_MAP = {
        8515: "Asian",
        2000000008: "More than one race",
        8557: "Native Hawaiian / Other PI",
        8516: "Black or African American",
        8527: "White",
        0: "Unknown",
    }
    ETHNICITY_HISPANIC_ID = 38003563  # "Hispanic or Latino"

    persons_df["sex_at_birth"] = persons_df["sex_at_birth_concept_id"].map(SEX_MAP).fillna("Other / Unknown")
    persons_df["race_group"] = persons_df["race_concept_id"].map(RACE_MAP).fillna("Unknown")
    persons_df["is_hispanic"] = np.where(
        persons_df["ethnicity_concept_id"] == ETHNICITY_HISPANIC_ID, "Hispanic or Latino", "Not Hispanic"
    )
    persons_df["race_ethnicity"] = np.where(
        persons_df["is_hispanic"] == "Hispanic or Latino", "Hispanic or Latino", persons_df["race_group"]
    )

    # Compute age at LAST observed end date (year granularity)
    persons_df["last_obs_end_date"] = pd.to_datetime(persons_df["last_obs_end_date"])
    persons_df["age_at_obs_end"] = (persons_df["last_obs_end_date"].dt.year - persons_df["year_of_birth"]).astype("Int64")

    # Normalize person ZIP3 BEFORE join (preserve leading zeros)
    persons_df["zip3"] = _norm_zip3_series(persons_df["zip3"])

    # SES join
    if not ses_df.empty:
        before_merge = persons_df["person_id"].nunique()
        persons_df = persons_df.merge(
            ses_df.rename(columns={"zip3_key": "zip3"}),
            on="zip3",
            how="left",
        )
        after_merge = persons_df["person_id"].nunique()

        left_non_null = persons_df["zip3"].notna().sum()
        matched = persons_df["deprivation_index"].notna().sum()
        match_rate = (matched / left_non_null * 100.0) if left_non_null else 0.0
        print(f"    - SES join status: {_ses_status}; joined on ZIP3 (pre={before_merge:,}, post={after_merge:,}).")
        print(f"    - SES match coverage: zip3 non-null on left = {left_non_null:,}; matched = {matched:,} ({match_rate:.1f}%).")
        # Persist tiny metrics sidecar for provenance
        try:
            import json
            with open(ses_metrics_path, "w", encoding="utf-8") as f:
                json.dump(
                    {
                        "workspace_cdr": WORKSPACE_CDR,
                        "ses_table_fqn": SES_TABLE_FQN,
                        "left_zip3_non_null": int(left_non_null),
                        "matched": int(matched),
                        "match_rate_pct": float(round(match_rate, 3)),
                        "ses_unique_zip3": int(ses_df["zip3"].nunique()) if "zip3" in ses_df.columns else int(ses_df["zip3_key"].nunique()),
                    },
                    f,
                    indent=2,
                )
        except Exception:
            pass
    else:
        persons_df["deprivation_index"] = np.nan
        print(f"    - SES join skipped: {_ses_status}.")

    # Impute missing deprivation_index to median (prefer observed, else map median, else 0.5)
    persons_df["deprivation_index"] = pd.to_numeric(persons_df["deprivation_index"], errors="coerce")
    missing_di = persons_df["deprivation_index"].isna().sum()
    if missing_di > 0:
        if persons_df["deprivation_index"].notna().any():
            median_di = float(persons_df["deprivation_index"].median())
        elif not ses_df.empty and "deprivation_index" in ses_df.columns:
            median_di = float(ses_df["deprivation_index"].median())
        else:
            median_di = 0.5  # last-resort neutral-ish value on 0–1 scale
        persons_df["deprivation_index"] = persons_df["deprivation_index"].fillna(median_di)
        print(f"    - Imputed {missing_di:,} missing `deprivation_index` values to median = {median_di:.3f}.")

    # Final column selection (keep original shape for downstream compatibility)
    final_cols = [
        "person_id",
        "year_of_birth",
        "age_at_obs_end",
        "sex_at_birth",
        "race_ethnicity",
        "deprivation_index",   # SES proxy (0–1 scale typical)
        "total_obs_days",
    ]
    persons_df = persons_df[final_cols].copy()

    # -----------------------------------------------------------------------------------
    # 5) Cache both artifacts
    # -----------------------------------------------------------------------------------
    print(f"    - Caching persons_df → {persons_out_path.name}")
    persons_df.to_parquet(persons_out_path, index=False)

    print(f"    - Caching observation_periods_df → {obs_periods_out_path.name}")
    observation_periods_df.to_parquet(obs_periods_out_path, index=False)

    # -----------------------------------------------------------------------------------
    # 6) Final summary and samples
    # -----------------------------------------------------------------------------------
    n_persons = len(persons_df)
    n_intervals = len(observation_periods_df)
    print("\n--- Build Summary ---")
    print(f"Eligible persons (>=365 total observed days): {n_persons:,}")
    print(f"Total observation intervals cached:           {n_intervals:,}")
    print("----------------------")

    print("\nSample of `persons_df` (with SES):")
    display(persons_df.head())

    print("\nSample of `observation_periods_df` (per-person intervals):")
    display(observation_periods_df.head())

    print(
        "\nℹ️  Downstream rule: validate any event/index/time_zero by joining to "
        "`observation_periods_df` and checking membership within an interval; "
        "derive censoring from the specific interval containing time_zero."
    )

    print("✅ Cell 2 (Re-Revised, SES Fix) complete.")


# In[ ]:


# ──────────────────────────── Cell 3 ─────────────────────────
# Phenotype Outcomes within Observation Periods + Leakage Blacklist
# -----------------------------------------------------------------------------

import json
from pathlib import Path
from functools import lru_cache

import pandas as pd
from google.cloud import bigquery

print("▶ Cell 3: Phenotyping outcomes (OP‑aware) & building leakage blacklist...")

# --- 0) Prerequisites & paths -------------------------------------------------
required_globals = ['CONFIG', 'client', 'WORKSPACE_CDR', 'persons_df']
for _name in required_globals:
    if _name not in globals():
        raise NameError(f"❌ Prerequisite '{_name}' not found. Please run the preceding cells first.")

ART_DIR = CONFIG.ARTIFACTS_DIR
labels_path         = ART_DIR / "labels_df.parquet"
blacklist_path      = ART_DIR / "leakage_blacklist.parquet"
build_meta_path     = ART_DIR / "labels_build_meta.json"

FORCE_REBUILD = False

def _load_build_meta(path: Path) -> dict:
    try:
        with open(path, "r", encoding="utf-8") as f:
            return json.load(f)
    except Exception:
        return {}

def _write_build_meta(path: Path, meta: dict):
    try:
        with open(path, "w", encoding="utf-8") as f:
            json.dump(meta, f, indent=2)
    except Exception as e:
        print(f"  • WARNING: Could not write build metadata ({e}). Proceeding without it.")

# Decide whether we can reuse cache
meta = _load_build_meta(build_meta_path)
expected_meta_keys = {
    "op_aware": True,
    "cdr_version": CONFIG.CDR_VERSION,
    "outcome_keys": sorted(list(CONFIG.OUTCOME_DEFINITIONS.keys())),
}
use_cache = (
    (not FORCE_REBUILD)
    and labels_path.exists()
    and blacklist_path.exists()
    and meta.get("op_aware") is True
    and meta.get("cdr_version") == expected_meta_keys["cdr_version"]
    and meta.get("outcome_keys") == expected_meta_keys["outcome_keys"]
)

if use_cache:
    labels_df = pd.read_parquet(labels_path)
    leakage_blacklist = pd.read_parquet(blacklist_path)
    print(f"  • Loaded OP‑aware caches: {labels_path.name}, {blacklist_path.name}")
else:
    print("  • No valid OP‑aware cache found. Building from BigQuery with observation‑period joins...")

    # --- 1) Build descendant sets per outcome (for leakage blacklist) ----------
    # We keep this simple & robust: one query per outcome (few outcomes total).
    all_rows = []
    for outcome_name, ancestor_ids in CONFIG.OUTCOME_DEFINITIONS.items():
        # Query all descendants for the outcome’s ancestor(s)
        sql = f"""
            SELECT DISTINCT descendant_concept_id
            FROM `{WORKSPACE_CDR}.concept_ancestor`
            WHERE ancestor_concept_id IN UNNEST(@ancestors)
        """
        job_config = bigquery.QueryJobConfig(
            query_parameters=[bigquery.ArrayQueryParameter("ancestors", "INT64", ancestor_ids)]
        )
        df = client.query(sql, job_config=job_config).to_dataframe()

        # Include the ancestor concepts themselves (defensive)
        descendants = set(df['descendant_concept_id'].dropna().astype(int).tolist()) | set(ancestor_ids)
        all_rows.extend(
            {"outcome": outcome_name, "descendant_concept_id": int(cid)} for cid in descendants
        )

    leakage_blacklist = pd.DataFrame(all_rows).drop_duplicates().reset_index(drop=True)
    print(f"    - Leakage blacklist built with {len(leakage_blacklist):,} (outcome, concept) mappings.")

    # --- 2) Condition outcomes: first event *within* an observation period ------
    eligible_pids = persons_df['person_id'].dropna().astype(int).tolist()
    if not eligible_pids:
        raise RuntimeError("No eligible person_ids found in `persons_df`.")

    # All concept IDs across outcomes (non-death)
    all_outcome_concept_ids = sorted(leakage_blacklist['descendant_concept_id'].unique().astype(int).tolist())

    print("    - Querying condition_occurrence with OP join...")
    condition_sql = f"""
    WITH cond AS (
      SELECT
        person_id,
        condition_concept_id AS concept_id,
        condition_start_date AS event_date
      FROM `{WORKSPACE_CDR}.condition_occurrence`
      WHERE person_id IN UNNEST(@person_ids)
        AND condition_concept_id IN UNNEST(@concept_ids)
    ),
    cond_in_obs AS (
      SELECT c.person_id, c.concept_id, c.event_date
      FROM cond c
      JOIN `{WORKSPACE_CDR}.observation_period` op
        ON c.person_id = op.person_id
       AND c.event_date BETWEEN op.observation_period_start_date AND op.observation_period_end_date
    )
    SELECT person_id, concept_id, MIN(event_date) AS first_date
    FROM cond_in_obs
    GROUP BY person_id, concept_id
    """
    job_config = bigquery.QueryJobConfig(
        query_parameters=[
            bigquery.ArrayQueryParameter("person_ids", "INT64", eligible_pids),
            bigquery.ArrayQueryParameter("concept_ids", "INT64", all_outcome_concept_ids),
        ]
    )
    condition_firsts = client.query(condition_sql, job_config=job_config).to_dataframe()

    # Map concepts → outcome and collapse to first event per (person, outcome)
    cond_labeled = condition_firsts.merge(
        leakage_blacklist,
        left_on="concept_id",
        right_on="descendant_concept_id",
        how="inner"
    )
    condition_labels_df = (
        cond_labeled.groupby(['person_id', 'outcome'], as_index=False)['first_date'].min()
    )

    # --- 3) Death outcome: first death *within* an observation period -----------
    print("    - Querying death tables with OP join...")
    death_sql = f"""
    WITH all_deaths AS (
      SELECT person_id, death_date AS event_date FROM `{WORKSPACE_CDR}.death`
      UNION ALL
      SELECT person_id, death_date AS event_date FROM `{WORKSPACE_CDR}.aou_death`
    ),
    deaths_in_obs AS (
      SELECT d.person_id, d.event_date
      FROM all_deaths d
      JOIN `{WORKSPACE_CDR}.observation_period` op
        ON d.person_id = op.person_id
       AND d.event_date BETWEEN op.observation_period_start_date AND op.observation_period_end_date
      WHERE d.event_date IS NOT NULL
        AND d.person_id IN UNNEST(@person_ids)
    )
    SELECT person_id, MIN(event_date) AS first_date
    FROM deaths_in_obs
    GROUP BY person_id
    """
    job_config = bigquery.QueryJobConfig(
        query_parameters=[bigquery.ArrayQueryParameter("person_ids", "INT64", eligible_pids)]
    )
    death_labels_df = client.query(death_sql, job_config=job_config).to_dataframe()
    death_labels_df['outcome'] = 'death'

    # --- 4) Combine & cache -----------------------------------------------------
    labels_df = pd.concat(
        [condition_labels_df, death_labels_df[['person_id', 'outcome', 'first_date']]],
        ignore_index=True
    ).rename(columns={'first_date': 'first_event_date'})

    # Normalize types
    labels_df['first_event_date'] = pd.to_datetime(labels_df['first_event_date'], errors='coerce')
    labels_df['person_id'] = labels_df['person_id'].astype('Int64')

    # Save artifacts
    labels_df.to_parquet(labels_path, index=False)
    leakage_blacklist.to_parquet(blacklist_path, index=False)

    # Save (or refresh) build meta
    _write_build_meta(build_meta_path, {
        **expected_meta_keys,
        "n_persons": int(len(persons_df)),
        "n_labels": int(len(labels_df)),
        "n_blacklist_rows": int(len(leakage_blacklist)),
        "notes": "Outcomes phenotyped with observation‑period joins; death restricted to OP as well."
    })

    print(f"    - Cached OP‑aware labels to: {labels_path.name}")
    print(f"    - Cached leakage blacklist to: {blacklist_path.name}")

# --- 5) Final summary & samples ----------------------------------------------
n_by_outcome = labels_df.groupby('outcome')['person_id'].nunique().sort_values(ascending=False)
print("\n  • Persons with ≥1 event by outcome (within OP):")
for oc, n in n_by_outcome.items():
    print(f"    - {oc}: {n:,}")

print(f"\n  • Final `labels_df`: {len(labels_df):,} rows; columns = {list(labels_df.columns)}")
print(f"  • Final `leakage_blacklist`: {len(leakage_blacklist):,} rows; columns = {list(leakage_blacklist.columns)}")

print("\nSample of `labels_df`: (person_id, outcome, first_event_date)")
display(labels_df.head())

print("\nSample of `leakage_blacklist`: (outcome, descendant_concept_id)")
display(leakage_blacklist.head())

print("✅ Cell 3 complete: OP‑aware outcome phenotypes and leakage blacklist are ready.")


# In[ ]:


# ──────────────────────────── Cell 4 ─────────────────────────────
# Build Master Predictor Event Ledger with Observation-Interval Join
# ------------------------------------------------------------------------------------------------

import pandas as pd
from pathlib import Path
import time

log("▶ Cell 4: Building Master Predictor Event Ledger...")

# --- 1) Define paths and check for cache -----------------------------------------------------------------
output_path = CONFIG.ARTIFACTS_DIR / "events_df.parquet"

if output_path.exists():
    events_df = pd.read_parquet(output_path)
    log(f"  • Loaded cached master event ledger from: {output_path.name}")
else:
    log("  • No cache found. Building event ledger from scratch…")
    t_start_cell = time.perf_counter()

    # --- 2) Prepare curated feature set and leakage blacklist --------------------------------------------
    log("    - Preparing curated feature set and leakage blacklist…")
    curated_features_df = pd.read_csv(CONFIG.CURATED_FEATURES_CSV_PATH)
    approved_features = curated_features_df[curated_features_df['include_in_model'] == 1]

    vocab_df = pd.read_parquet(CONFIG.SOURCE_CACHE_DIR / "event_vocab_v1.parquet")

    # Map curated vocab_event_id → OMOP concept_id (ancestor) for roll-up
    curated_concept_ids = (
        vocab_df.loc[vocab_df['vocab_event_id'].isin(approved_features['vocab_event_id']),
                     'concept_id']
        .dropna().astype(int).tolist()
    )

    # Leakage blacklist (all outcome descendant concept_ids) — exclude these from predictors
    leakage_blacklist_df = pd.read_parquet(CONFIG.ARTIFACTS_DIR / "leakage_blacklist.parquet")
    blacklisted_descendant_ids = set(leakage_blacklist_df['descendant_concept_id'].astype(int))

    log(f"    - Curated ancestors: {len(curated_concept_ids):,}  |  Blacklisted descendants: {len(blacklisted_descendant_ids):,}")

    if not curated_concept_ids:
        raise RuntimeError("Curated concept list is empty. Check CONFIG.CURATED_FEATURES_CSV_PATH and vocab mapping.")

    # --- 3) Build descendant map for hierarchical roll‑up (ancestor → all descendants) -------------------
    log("    - Querying BigQuery to build (ancestor → descendant) roll‑up map…")
    desc_map_sql = f"""
    SELECT
      ca.descendant_concept_id,
      ca.ancestor_concept_id AS feature_concept_id
    FROM `{WORKSPACE_CDR}.concept_ancestor` AS ca
    WHERE ca.ancestor_concept_id IN UNNEST(@curated_concept_ids)
    """
    job_config = bigquery.QueryJobConfig(
        query_parameters=[bigquery.ArrayQueryParameter("curated_concept_ids", "INT64", curated_concept_ids)]
    )
    desc_map_df = client.query(desc_map_sql, job_config=job_config).to_dataframe()

    if desc_map_df.empty:
        raise RuntimeError("No descendants found for curated features. Verify curated_concept_ids and `concept_ancestor`.")

    # Drop blacklisted descendants (i.e., outcome concepts) from the predictor map
    before = len(desc_map_df)
    desc_map_df = desc_map_df[~desc_map_df['descendant_concept_id'].isin(blacklisted_descendant_ids)].copy()
    after = len(desc_map_df)
    log(f"    - Descendant map rows: {after:,} (removed {before - after:,} blacklisted mappings).")
    n_descendants = desc_map_df['descendant_concept_id'].nunique()
    log(f"    - Unique raw descendant concept_ids to search: {n_descendants:,}")

    # --- 4) Chunked extraction of raw events with observation‑period JOIN --------------------------------
    log("    - Querying raw events in chunks WITH observation_period join…")

    CHUNK_SIZE = 20_000
    all_event_frames = []
    eligible_pids = persons_df['person_id'].astype(int).tolist()  # persons_df should already reflect eligibility

    # Optional progress bar
    iterator = range(0, len(desc_map_df), CHUNK_SIZE)
    if 'tqdm' in globals() and tqdm:
        iterator = tqdm(iterator, desc="Querying Event Chunks", unit="chunk")

    for start_idx in iterator:
        chunk = desc_map_df.iloc[start_idx:start_idx + CHUNK_SIZE].copy()

        chunk_desc_ids = chunk['descendant_concept_id'].astype(int).tolist()
        chunk_feat_ids = chunk['feature_concept_id'].astype(int).tolist()

        if not chunk_desc_ids:
            continue

        # NOTE:
        #  • Join to observation_period (`op`) ensures event_date ∈ [op_start, op_end] for that person.
        #  • We prefilter each domain subquery by concept_id IN @chunk_desc_ids to reduce scan volume.
        raw_events_sql = f"""
        WITH
          descendant_map AS (
            SELECT
              desc_id AS descendant_concept_id,
              feat_id AS feature_concept_id
            FROM UNNEST(@chunk_desc_ids) AS desc_id WITH OFFSET pos
            JOIN UNNEST(@chunk_feat_ids) AS feat_id WITH OFFSET pos USING(pos)
          ),
          raw_condition AS (
            SELECT person_id,
                   condition_start_date AS event_date,
                   condition_concept_id AS concept_id_raw
            FROM `{WORKSPACE_CDR}.condition_occurrence`
            WHERE condition_concept_id IN UNNEST(@chunk_desc_ids)
          ),
          raw_procedure AS (
            SELECT person_id,
                   procedure_date AS event_date,
                   procedure_concept_id AS concept_id_raw
            FROM `{WORKSPACE_CDR}.procedure_occurrence`
            WHERE procedure_concept_id IN UNNEST(@chunk_desc_ids)
          ),
          raw_drug AS (
            SELECT person_id,
                   drug_exposure_start_date AS event_date,
                   drug_concept_id AS concept_id_raw
            FROM `{WORKSPACE_CDR}.drug_exposure`
            WHERE drug_concept_id IN UNNEST(@chunk_desc_ids)
          ),
          raw_events AS (
            SELECT * FROM raw_condition
            UNION ALL
            SELECT * FROM raw_procedure
            UNION ALL
            SELECT * FROM raw_drug
          )
        SELECT
          re.person_id,
          re.event_date,
          dm.feature_concept_id
        FROM raw_events AS re
        JOIN descendant_map AS dm
          ON re.concept_id_raw = dm.descendant_concept_id
        JOIN `{WORKSPACE_CDR}.observation_period` AS op
          ON re.person_id = op.person_id
         AND re.event_date BETWEEN op.observation_period_start_date AND op.observation_period_end_date
        WHERE re.person_id IN UNNEST(@eligible_pids)
          AND re.event_date IS NOT NULL
        """

        job_config = bigquery.QueryJobConfig(
            query_parameters=[
                bigquery.ArrayQueryParameter("eligible_pids",   "INT64", eligible_pids),
                bigquery.ArrayQueryParameter("chunk_desc_ids",  "INT64", chunk_desc_ids),
                bigquery.ArrayQueryParameter("chunk_feat_ids",  "INT64", chunk_feat_ids),
            ]
        )

        try:
            chunk_df = client.query(raw_events_sql, job_config=job_config).to_dataframe()
            if not chunk_df.empty:
                all_event_frames.append(chunk_df)
        except Exception as e:
            log(f"      - ERROR on chunk starting at {start_idx:,}: {e}")
            log("      - Continuing with next chunk…")
            continue

    if not all_event_frames:
        raise RuntimeError(
            "No events were retrieved after joining to observation_period. "
            "Check curated feature list, blacklist, and person eligibility."
        )

    raw_events_df = pd.concat(all_event_frames, ignore_index=True)
    raw_events_df['event_date'] = pd.to_datetime(raw_events_df['event_date'])
    log(f"    - Retrieved {len(raw_events_df):,} raw event rows after observation-period filtering.")

    # --- 5) Deduplicate to first occurrence per (person_id, event_date, feature) -------------------------
    log("    - Deduplicating to one row per (person_id, event_date, feature_concept_id)…")
    events_df = (
        raw_events_df
        .sort_values(['person_id', 'event_date', 'feature_concept_id'])
        .drop_duplicates(subset=['person_id', 'event_date', 'feature_concept_id'])
        .reset_index(drop=True)
    )

    # Final schema
    events_df = events_df[['person_id', 'event_date', 'feature_concept_id']]

    # --- 6) Cache the result --------------------------------------------------------------------------------
    log(f"    - Caching final event ledger to: {output_path.name}")
    events_df.to_parquet(output_path, index=False)
    log(f"  • Cell 4 completed in {(time.perf_counter() - t_start_cell) / 60:.1f} minutes.")

# --- Final summary & sample ----------------------------------------------------------------------------------------
log(f"  • Final `events_df` has {len(events_df):,} rows and {len(events_df.columns)} columns.")
log("✅ Cell 4 complete: Master predictor event ledger (interval‑accurate) is ready.")

# Display a quick sample
display(events_df.head())


# In[ ]:


# ──────────────────────────── Cell 4a (NEW) ──────────────────────────────────
# Prepare Artifacts for CCI and Utilization (OP‑aware)
# -----------------------------------------------------------------------------
# This cell builds two lightweight, analysis‑ready artifacts that power the
# new confounders in Cell 6:
#
#   1) artifacts/charlson_events_df.parquet
#        • Columns: person_id, event_date, charlson_condition_id, charlson_category
#        • One row per *observed* Charlson condition event (within an observation period)
#        • charlson_condition_id is the ancestor concept_id representing the Charlson category
#
#   2) artifacts/visits_df.parquet
#        • Columns: person_id, visit_start_date
#        • All *observed* visits (within an observation period) for eligible persons
#
# Notes:
#   • OP‑aware: All dates are restricted to be within a valid observation interval.
#   • The Charlson ancestor set can be provided via
#       - SOURCE_CACHE_DIR/vocab/charlson_omop_ancestors.csv, or
#       - CONFIG_DIR/charlson_omop_ancestors.csv
#     with (at minimum) columns: ['charlson_category','ancestor_concept_id'].
#     If no file is found, we fall back to a *partial* mapping using overlapping
#     concepts defined in CONFIG.OUTCOME_DEFINITIONS (MI, dementia, leukemia, lymphoma).
#
# Requirements from prior cells:
#   • CONFIG, client, WORKSPACE_CDR, persons_df
# -----------------------------------------------------------------------------


import pandas as pd
import numpy as np
from pathlib import Path
from google.cloud import bigquery
import time

print("▶ Cell 4a: Building OP‑aware Charlson and Visits artifacts…")

# --- 0) Prerequisites & paths ------------------------------------------------
required_globals = ['CONFIG', 'client', 'WORKSPACE_CDR', 'persons_df']
for _name in required_globals:
    if _name not in globals():
        raise NameError(f"❌ Prerequisite '{_name}' not found. Please run the preceding cells first.")

ART_DIR = CONFIG.ARTIFACTS_DIR
ART_DIR.mkdir(parents=True, exist_ok=True)

charlson_out_path = ART_DIR / "charlson_events_df.parquet"
visits_out_path   = ART_DIR / "visits_df.parquet"

# Allow idempotent runs (build only missing artifacts)
NEED_CHARLSON = not charlson_out_path.exists()
NEED_VISITS   = not visits_out_path.exists()

if not NEED_CHARLSON and not NEED_VISITS:
    # Load and show brief summary if both exist
    _char = pd.read_parquet(charlson_out_path)
    _vis  = pd.read_parquet(visits_out_path)
    print(f"  • Loaded cached charlson_events_df: {charlson_out_path.name} ({len(_char):,} rows)")
    print(f"  • Loaded cached visits_df:          {visits_out_path.name} ({len(_vis):,} rows)")
    display(_char.head(3))
    display(_vis.head(3))
    print("✅ Cell 4a complete (loaded from cache).")
else:
    eligible_pids = persons_df['person_id'].dropna().astype(int).tolist()
    if not eligible_pids:
        raise RuntimeError("No eligible person_ids found in `persons_df`.")

    # -------------------------------------------------------------------------
    # 1) Load/resolve Charlson ancestor concept_ids (robust with partial fallback)
    # -------------------------------------------------------------------------
    def _load_charlson_ancestors() -> pd.DataFrame:
        """
        Returns a DataFrame with columns:
          • charlson_category (str)
          • ancestor_concept_id (int)
        Tries to read from CSV; falls back to a partial set from CONFIG.OUTCOME_DEFINITIONS.
        """
        candidate_paths = [
            CONFIG.CONFIG_DIR / "charlson_omop_ancestors.csv",
            CONFIG.SOURCE_CACHE_DIR / "charlson_omop_ancestors.csv",
        ]
        for path in candidate_paths:
            if path.exists():
                df = pd.read_csv(path)
                # Flexible column normalization
                cols = {c.lower(): c for c in df.columns}
                # Support either a single 'ancestor_concept_id' column or a semicolon‑separated list column
                if 'ancestor_concept_ids' in cols:
                    tmp = (
                        df[[cols.get('charlson_category', 'charlson_category'),
                            cols['ancestor_concept_ids']]]
                        .rename(columns={
                            cols.get('charlson_category', 'charlson_category'): 'charlson_category',
                            cols['ancestor_concept_ids']: 'ancestor_concept_ids'
                        })
                    )
                    tmp['ancestor_concept_ids'] = tmp['ancestor_concept_ids'].astype(str)
                    tmp = tmp.assign(
                        ancestor_concept_id=tmp['ancestor_concept_ids'].str.split(';')
                    ).explode('ancestor_concept_id')
                    tmp['ancestor_concept_id'] = pd.to_numeric(tmp['ancestor_concept_id'], errors='coerce').astype('Int64')
                    out = tmp.drop(columns=['ancestor_concept_ids']).dropna(subset=['ancestor_concept_id'])
                else:
                    col_cat = cols.get('charlson_category', 'charlson_category')
                    col_anc = cols.get('ancestor_concept_id', 'ancestor_concept_id')
                    out = df[[col_cat, col_anc]].rename(columns={col_cat: 'charlson_category',
                                                                 col_anc: 'ancestor_concept_id'})
                out['ancestor_concept_id'] = pd.to_numeric(out['ancestor_concept_id'], errors='coerce').astype('Int64')
                out = out.dropna(subset=['ancestor_concept_id'])
                out['charlson_category'] = out['charlson_category'].astype(str)
                if out.empty:
                    raise ValueError(f"Charlson CSV found at {path} but no valid rows after parsing.")
                print(f"  • Loaded Charlson ancestor set from: {path.name} "
                      f"({out['charlson_category'].nunique()} categories, {len(out):,} rows).")
                return out.reset_index(drop=True)

        # Fallback: build a *partial* mapping using overlapping concepts
        print("  • No Charlson mapping file found. Falling back to a *partial* mapping "
              "derived from CONFIG.OUTCOME_DEFINITIONS (MI, dementia, leukemia, lymphoma).")
        fallback_rows = []
        # Map a subset of Charlson categories to available ancestor sets
        _map = {
            'myocardial_infarction': 'myocardial_infarction',
            'dementia': 'dementia',
            'leukemia': 'leukemia',
            'lymphoma': 'lymphoma',
        }
        for cat, cfg_key in _map.items():
            if cfg_key in CONFIG.OUTCOME_DEFINITIONS:
                for anc in CONFIG.OUTCOME_DEFINITIONS[cfg_key]:
                    fallback_rows.append({'charlson_category': cat,
                                          'ancestor_concept_id': int(anc)})
        if not fallback_rows:
            raise FileNotFoundError(
                "No Charlson mapping CSV was found and the fallback mapping could not be built.\n"
                "Please provide 'charlson_omop_ancestors.csv' with columns "
                "['charlson_category','ancestor_concept_id'] in either:\n"
                f"  • {CONFIG.SOURCE_CACHE_DIR / 'vocab'}\n"
                f"  • {CONFIG.CONFIG_DIR}\n"
                "Each row should map a Charlson category to a standard OMOP ancestor concept_id."
            )
        print(f"    - Fallback contains {len(fallback_rows)} ancestor rows across "
              f"{len(set(r['charlson_category'] for r in fallback_rows))} categories.")
        return pd.DataFrame(fallback_rows)

    # Build the Charlson ancestor set if needed
    if NEED_CHARLSON:
        t0 = time.perf_counter()
        charlson_anc_df = _load_charlson_ancestors().copy()
        charlson_anc_df['ancestor_concept_id'] = charlson_anc_df['ancestor_concept_id'].astype(int)

        # Fetch all descendants for these ancestors (for filtering condition_occurrence)
        print("    - Resolving Charlson descendant concepts via concept_ancestor…")
        anc_list = charlson_anc_df['ancestor_concept_id'].dropna().astype(int).unique().tolist()

        desc_sql = f"""
        SELECT ancestor_concept_id, descendant_concept_id
        FROM `{WORKSPACE_CDR}.concept_ancestor`
        WHERE ancestor_concept_id IN UNNEST(@ancestors)
        """
        job_config = bigquery.QueryJobConfig(
            query_parameters=[bigquery.ArrayQueryParameter("ancestors", "INT64", anc_list)]
        )
        desc_map_df = client.query(desc_sql, job_config=job_config).to_dataframe()
        if desc_map_df.empty:
            raise RuntimeError("concept_ancestor returned 0 rows for the provided Charlson ancestor set.")

        # Keep only (desc, anc) pairs that map to a known category
        desc_map_df = desc_map_df.merge(
            charlson_anc_df.rename(columns={'ancestor_concept_id': 'ancestor_concept_id',
                                            'charlson_category': 'charlson_category'}),
            on='ancestor_concept_id', how='left'
        )
        desc_map_df = desc_map_df.dropna(subset=['charlson_category']).reset_index(drop=True)
        desc_map_df['descendant_concept_id'] = desc_map_df['descendant_concept_id'].astype(int)
        desc_ids = desc_map_df['descendant_concept_id'].unique().tolist()
        print(f"    - Charlson mapping pairs: {len(desc_map_df):,}  |  unique descendants: {len(desc_ids):,}")

        # Query condition_occurrence for these descendants, OP‑aware, for eligible persons
        print("    - Querying condition_occurrence with OP join (small-parameter SQL)…")

        # Only the ~23 Charlson ancestors go to BigQuery as a parameter.
        anc_list = charlson_anc_df['ancestor_concept_id'].dropna().astype(int).unique().tolist()

        charlson_sql_small = f"""
        WITH dm AS (
        SELECT
            ca.descendant_concept_id,
            ca.ancestor_concept_id AS charlson_condition_id
        FROM `{WORKSPACE_CDR}.concept_ancestor` AS ca
        WHERE ca.ancestor_concept_id IN UNNEST(@ancestors)
        ),
        raw_cond AS (
        SELECT
            person_id,
            condition_start_date AS event_date,
            condition_concept_id AS concept_id_raw
        FROM `{WORKSPACE_CDR}.condition_occurrence`
        WHERE condition_concept_id IN (SELECT descendant_concept_id FROM dm)
            AND condition_start_date IS NOT NULL
        ),
        op_filtered AS (
        SELECT rc.person_id, rc.event_date, rc.concept_id_raw
        FROM raw_cond rc
        JOIN `{WORKSPACE_CDR}.observation_period` op
            ON rc.person_id = op.person_id
        AND rc.event_date BETWEEN op.observation_period_start_date AND op.observation_period_end_date
        )
        SELECT opf.person_id, opf.event_date, dm.charlson_condition_id
        FROM op_filtered AS opf
        JOIN dm
            ON opf.concept_id_raw = dm.descendant_concept_id
        """

        job_config = bigquery.QueryJobConfig(
            query_parameters=[
                bigquery.ArrayQueryParameter("ancestors", "INT64", anc_list)
            ]
        )

        # Increase begin() timeout defensively; the payload is small so this won't stall the POST.
        charlson_events_df = client.query(charlson_sql_small, job_config=job_config, timeout=1200).to_dataframe()

        # Keep only eligible persons (same cohort as persons_df) to match downstream expectations
        charlson_events_df = charlson_events_df.merge(
            persons_df[['person_id']], on='person_id', how='inner'
        )

        # Normalize types
        charlson_events_df['event_date'] = pd.to_datetime(charlson_events_df['event_date'], errors='coerce')
        charlson_events_df['person_id'] = pd.to_numeric(charlson_events_df['person_id'], errors='coerce').astype('Int64')
        charlson_events_df['charlson_condition_id'] = pd.to_numeric(
            charlson_events_df['charlson_condition_id'], errors='coerce'
        ).astype('Int64')

        # Attach category labels (optional but nice to have)
        charlson_events_df = charlson_events_df.merge(
            charlson_anc_df.rename(columns={'ancestor_concept_id': 'charlson_condition_id'}),
            on='charlson_condition_id', how='left'
        )

        # De‑dupe exact duplicates
        charlson_events_df = (
            charlson_events_df
            .drop_duplicates(subset=['person_id', 'event_date', 'charlson_condition_id'])
            .reset_index(drop=True)
        )

        # Cache as before
        charlson_events_df.to_parquet(charlson_out_path, index=False)
        print(f"  • Saved Charlson events → {charlson_out_path.name} ({len(charlson_events_df):,} rows "
            f"across {charlson_events_df['charlson_condition_id'].nunique()} categories).")
        if len(charlson_events_df):
            display(charlson_events_df.head(5))


    # -------------------------------------------------------------------------
    # 2) Build OP‑aware visits_df (all visits for eligible persons)
    # -------------------------------------------------------------------------
    if NEED_VISITS:
        t0 = time.perf_counter()
        print("    - Querying visit_occurrence with OP join (eligible persons)…")
        visits_sql = f"""
        WITH raw_visits AS (
          SELECT person_id, visit_start_date
          FROM `{WORKSPACE_CDR}.visit_occurrence`
          WHERE person_id IN UNNEST(@pids)
            AND visit_start_date IS NOT NULL
        ),
        op_filtered AS (
          SELECT rv.person_id, rv.visit_start_date
          FROM raw_visits rv
          JOIN `{WORKSPACE_CDR}.observation_period` op
            ON rv.person_id = op.person_id
           AND rv.visit_start_date BETWEEN op.observation_period_start_date AND op.observation_period_end_date
        )
        SELECT person_id, visit_start_date
        FROM op_filtered
        """
        job_config = bigquery.QueryJobConfig(
            query_parameters=[bigquery.ArrayQueryParameter("pids", "INT64", eligible_pids)]
        )
        visits_df = client.query(visits_sql, job_config=job_config).to_dataframe()
        if visits_df.empty:
            print("    - WARNING: No visits found within observation periods for eligible persons.")

        # Normalize & de‑dupe
        visits_df['visit_start_date'] = pd.to_datetime(visits_df['visit_start_date'], errors='coerce')
        visits_df['person_id'] = pd.to_numeric(visits_df['person_id'], errors='coerce').astype('Int64')
        visits_df = visits_df.dropna(subset=['visit_start_date']).drop_duplicates().reset_index(drop=True)

        # Cache
        visits_df.to_parquet(visits_out_path, index=False)
        dt = time.perf_counter() - t0
        print(f"  • Saved visits → {visits_out_path.name} "
              f"({len(visits_df):,} rows; {visits_df['person_id'].nunique():,} persons) in {dt:.1f}s.")
        # Show sample
        if len(visits_df):
            display(visits_df.head(5))

    print("✅ Cell 4a complete: OP‑aware Charlson and visits artifacts are ready.")


# In[ ]:


# ──────────────────────────── Cell 4b ───────────────────────────
# OP‑aware Derived Smoking & BMI events using notebook hardcodes; append to events_df safely.
# -----------------------------------------------------------------------------------------------
# Outputs:
#   • artifacts/derived_covariates_df.parquet
#   • artifacts/events_df.parquet
#   • artifacts/derived_vocab_patch.parquet

import pandas as pd
import numpy as np
from google.cloud import bigquery

print("▶ Cell 4b: Building OP‑aware Derived Smoking & BMI events (hardcoded IDs) and appending to events_df…")

# --- Preconditions & paths
required_globals = ['CONFIG', 'client', 'WORKSPACE_CDR', 'persons_df']
for _name in required_globals:
    if _name not in globals():
        raise NameError(f"❌ Prerequisite '{_name}' not found. Please run the preceding cells first.")

ART_DIR = CONFIG.ARTIFACTS_DIR
ART_DIR.mkdir(parents=True, exist_ok=True)

eligible_pids = persons_df['person_id'].dropna().astype(int).tolist()
if not eligible_pids:
    raise RuntimeError("No eligible person_ids found in `persons_df`.")

# Pretty display if available
try:
    from IPython.display import display
except Exception:
    def display(x):
        print(x.head() if hasattr(x, "head") else x)

# --- Hardcodes from <ids_and_hardcodes>
SMOKING_PPI_QID = 1585857  # observation_source_concept_id

SMOKING_STRING_MAP = {
    "LA32-8": "Never",
    "100CigsLifetime_No": "Never",
    "LA33-6": "Ever",
    "100CigsLifetime_Yes": "Ever",
    "LA12688-0": "Unknown",
    "PMI_DontKnow": "Unknown",
    "LA29631-1": "Unknown",
    "PMI_PreferNotToAnswer": "Unknown",
    "PMI_Skip": "Unknown"
}
SMOKING_DERIVED_ID = {"Ever": 9000001, "Never": 9000002, "Unknown": 9000003}

BMI_MEASUREMENT_CONCEPT_IDS = [903124, 3038553]
def _bmi_bucket(v):
    if pd.isna(v): return None
    v = float(v)
    return ("Underweight"      if v < 18.5 else
            "Normal weight"    if v < 25.0 else
            "Overweight"       if v < 30.0 else
            "Obese class I"    if v < 35.0 else
            "Obese class II+")

BMI_DERIVED_ID = {
    "Underweight":     9000004,
    "Normal weight":   9000005,
    "Overweight":      9000006,
    "Obese class I":   9000007,
    "Obese class II+": 9000008
}

# --- Helper: BigQuery runner
def _bq_df(sql, params):
    jc = bigquery.QueryJobConfig(query_parameters=params)
    return client.query(sql, job_config=jc).to_dataframe()

# --- 1) Smoking from PPI (OP‑aware)
print("    - Querying PPI smoking responses (OP‑aware)…")
smk_sql = f"""
WITH raw AS (
  SELECT person_id,
         observation_date AS event_date,
         value_as_string
  FROM `{WORKSPACE_CDR}.observation`
  WHERE observation_source_concept_id = @ppi_q
    AND value_as_string IS NOT NULL
    AND person_id IN UNNEST(@pids)
),
op_filtered AS (
  SELECT r.person_id, r.event_date, r.value_as_string
  FROM raw r
  JOIN `{WORKSPACE_CDR}.observation_period` op
    ON r.person_id = op.person_id
   AND r.event_date BETWEEN op.observation_period_start_date AND op.observation_period_end_date
)
SELECT * FROM op_filtered
"""
smk_df = _bq_df(smk_sql, [
    bigquery.ScalarQueryParameter("ppi_q", "INT64", SMOKING_PPI_QID),
    bigquery.ArrayQueryParameter("pids", "INT64", eligible_pids),
])

# Normalize & map
if not smk_df.empty:
    map_upper = {k.upper(): v for k, v in SMOKING_STRING_MAP.items()}
    smk_df['event_date'] = pd.to_datetime(smk_df['event_date'], errors='coerce')
    smk_df['person_id'] = pd.to_numeric(smk_df['person_id'], errors='coerce').astype('int64')

    smk_df['value'] = smk_df['value_as_string'].astype(str).str.strip().str.upper().map(map_upper).fillna("Unknown")
    smk_df['category'] = 'Smoking status'
    smk_df['feature_concept_id'] = smk_df['value'].map(SMOKING_DERIVED_ID).astype('int64')

    smk_events = (smk_df[['person_id','event_date','category','value','feature_concept_id']]
                  .dropna(subset=['event_date'])
                  .drop_duplicates(subset=['person_id','event_date','category'])
                  .reset_index(drop=True))
else:
    smk_events = pd.DataFrame(columns=['person_id','event_date','category','value','feature_concept_id'])
print(f"      • Smoking rows fetched: {len(smk_df):,}  →  derived events: {len(smk_events):,}")

# --- 2) BMI from measurement (OP‑aware)
print("    - Querying BMI measurements (OP‑aware)…")
bmi_sql = f"""
WITH raw AS (
  SELECT person_id,
         measurement_date AS event_date,
         value_as_number AS bmi_value
  FROM `{WORKSPACE_CDR}.measurement`
  WHERE measurement_concept_id IN UNNEST(@bmi_ids)
    AND value_as_number IS NOT NULL
    AND person_id IN UNNEST(@pids)
),
op_filtered AS (
  SELECT r.person_id, r.event_date, r.bmi_value
  FROM raw r
  JOIN `{WORKSPACE_CDR}.observation_period` op
    ON r.person_id = op.person_id
   AND r.event_date BETWEEN op.observation_period_start_date AND op.observation_period_end_date
)
SELECT * FROM op_filtered
"""
bmi_df = _bq_df(bmi_sql, [
    bigquery.ArrayQueryParameter("bmi_ids", "INT64", BMI_MEASUREMENT_CONCEPT_IDS),
    bigquery.ArrayQueryParameter("pids", "INT64", eligible_pids),
])

if not bmi_df.empty:
    bmi_df['event_date'] = pd.to_datetime(bmi_df['event_date'], errors='coerce')
    bmi_df['person_id'] = pd.to_numeric(bmi_df['person_id'], errors='coerce').astype('int64')
    bmi_df['bmi_value'] = pd.to_numeric(bmi_df['bmi_value'], errors='coerce')
    bmi_df = bmi_df[bmi_df['bmi_value'].between(10, 80)]  # basic outlier guard
    bmi_df['category'] = 'BMI'
    bmi_df['value'] = bmi_df['bmi_value'].apply(_bmi_bucket)
    bmi_df['feature_concept_id'] = bmi_df['value'].map(BMI_DERIVED_ID).astype('int64')

    bmi_events = (bmi_df[['person_id','event_date','category','value','feature_concept_id']]
                  .dropna(subset=['event_date','feature_concept_id'])
                  .drop_duplicates(subset=['person_id','event_date','category'])
                  .reset_index(drop=True))
else:
    bmi_events = pd.DataFrame(columns=['person_id','event_date','category','value','feature_concept_id'])
print(f"      • BMI rows fetched: {len(bmi_df):,}  →  derived events: {len(bmi_events):,}")

# --- 3) Save human‑readable artifact
derived_cov = pd.concat([smk_events, bmi_events], ignore_index=True)
derived_cov = derived_cov.drop_duplicates(subset=['person_id','event_date','category','value']).reset_index(drop=True)
drv_path = ART_DIR / "derived_covariates_df.parquet"
derived_cov.to_parquet(drv_path, index=False)
print(f"  • Saved derived_covariates_df → {drv_path.name} ({len(derived_cov):,} rows)")
if len(derived_cov): display(derived_cov.head(5))

# --- 4) Append to the master ledger with **strict dtype control**
# Reload baseline ledger from disk if present (safer if a previous run left bad dtypes)
ev_path = ART_DIR / "events_df.parquet"
if ev_path.exists():
    base_events = pd.read_parquet(ev_path)
else:
    if 'events_df' not in globals():
        raise RuntimeError("events_df not found in memory and events_df.parquet not on disk.")
    base_events = events_df.copy()
    # Normalize types on the in‑memory base just in case
    base_events['event_date'] = pd.to_datetime(base_events['event_date'], errors='coerce')
    base_events['person_id'] = pd.to_numeric(base_events['person_id'], errors='coerce').astype('int64')
    base_events['feature_concept_id'] = pd.to_numeric(base_events['feature_concept_id'], errors='coerce').astype('int64')
    base_events = base_events.dropna(subset=['event_date'])

# Prepare *typed* new rows
new_events = derived_cov[['person_id','event_date','feature_concept_id']].copy()
new_events['event_date'] = pd.to_datetime(new_events['event_date'], errors='coerce')
new_events['person_id'] = pd.to_numeric(new_events['person_id'], errors='coerce').astype('int64')
new_events['feature_concept_id'] = pd.to_numeric(new_events['feature_concept_id'], errors='coerce').astype('int64')
new_events = new_events.dropna(subset=['event_date'])

before = len(base_events)
events_df = pd.concat([base_events, new_events], ignore_index=True)
events_df.drop_duplicates(subset=['person_id','event_date','feature_concept_id'], inplace=True)
events_df.sort_values(['person_id','event_date','feature_concept_id'], inplace=True)
after = len(events_df)

events_df.to_parquet(ev_path, index=False)
print(f"  • Appended Derived events to events_df: +{after - before:,} rows (now {after:,} total).")
print(f"  • Updated events_df saved → {ev_path.name}")

# --- 5) Optional: check for the 8 derived IDs in your vocab
try:
    vocab_path = CONFIG.SOURCE_CACHE_DIR / "event_vocab_v1.parquet"
    if vocab_path.exists():
        vocab_df = pd.read_parquet(vocab_path)
        required_ids = {9000001,9000002,9000003,9000004,9000005,9000006,9000007,9000008}
        present_ids = set(pd.to_numeric(vocab_df.get('concept_id', pd.Series(dtype='int64')), errors='coerce')
                          .dropna().astype('int64').tolist())
        missing = sorted(required_ids - present_ids)
        if missing:
            print(f"⚠️  event_vocab_v1.parquet is missing {len(missing)} derived concept_id(s): {missing}")
            patch_rows = [
                (9000001, "Smoking status · Ever",          "Derived"),
                (9000002, "Smoking status · Never",         "Derived"),
                (9000003, "Smoking status · Unknown",       "Derived"),
                (9000004, "BMI · Underweight",              "Derived"),
                (9000005, "BMI · Normal weight",            "Derived"),
                (9000006, "BMI · Overweight",               "Derived"),
                (9000007, "BMI · Obese class I",            "Derived"),
                (9000008, "BMI · Obese class II+",          "Derived"),
            ]
            vocab_patch = pd.DataFrame(patch_rows, columns=['concept_id','concept_name','source_domain'])
            patch_path = ART_DIR / "derived_vocab_patch.parquet"
            vocab_patch.to_parquet(patch_path, index=False)
            print(f"   Wrote a helper patch → {patch_path.name}. Merge this into your vocab before Cell 6 if needed.")
        else:
            print("  • All 8 derived concept IDs are present in event_vocab_v1.parquet ✔️")
    else:
        print("ℹ️  event_vocab_v1.parquet not found at SOURCE_CACHE_DIR — skipping vocab check.")
except Exception as e:
    print(f"ℹ️  Vocab presence check skipped due to: {e}")

print("✅ Cell 4b complete: Derived Smoking & BMI events are OP‑aware and merged into events_df.")


# In[ ]:


# ──────────────────────────── Cell 5 ─────────────────────────────────────────
# Create Discovery and Confirmation Sets
# ---------------------------------------------------------------------------
# This cell performs a stratified random split of the patient cohort into
# discovery (training) and confirmation (hold-out) sets. The split is
# stratified by age band, sex, and race/ethnicity to ensure that demographic
# distributions are preserved in both sets.
#
# The resulting sets of person_ids are cached to disk (`artifacts/
# split_assignments.pkl`), making this step fully resumable.

import pickle
import pandas as pd
from pathlib import Path

print("▶ Cell 5: Creating Discovery and Confirmation Sets...")

# --- 1. Define Path for Cached Artifact ---
output_path = CONFIG.ARTIFACTS_DIR / "split_assignments.pkl"

# --- 2. Resumability: Load from Cache if it Exists ---
if output_path.exists():
    print(f"  • Found cached split assignments. Loading from: {output_path.name}")
    with open(output_path, 'rb') as f:
        split_data = pickle.load(f)
    discovery_pids = split_data['discovery_pids']
    confirmation_pids = split_data['confirmation_pids']
else:
    print("  • No cache found. Performing a new stratified split...")

    # --- 3. Prepare Data for Stratification ---
    # The `persons_df` from Cell 2 is required for this step.
    if 'persons_df' not in globals():
        raise NameError("`persons_df` is not defined. Please run Cell 2 first.")

    # Create age bands for stratification, mirroring the logic from previous notebooks.
    print("    - Creating age bands for stratification...")
    def _get_age_band(age):
        if pd.isna(age):
            return "Unknown"
        if age < 18:
            return "<18"
        if age <= 29:
            return "18-29"
        if age <= 44:
            return "30-44"
        if age <= 59:
            return "45-59"
        if age <= 74:
            return "60-74"
        return ">=75"

    persons_df_copy = persons_df.copy()
    persons_df_copy['age_band'] = persons_df_copy['age_at_obs_end'].apply(_get_age_band)
    
    strata_cols = ['age_band', 'sex_at_birth', 'race_ethnicity']
    
    # --- 4. Perform the Stratified Split ---
    print(f"    - Performing stratified split with {CONFIG.DISCOVERY_FRAC:.0%} discovery...")
    
    # Use groupby().sample() for robust stratified sampling
    discovery_df = persons_df_copy.groupby(strata_cols, group_keys=False).sample(
        frac=CONFIG.DISCOVERY_FRAC,
        random_state=CONFIG.RNG_SEED
    )
    
    # The discovery set contains the person_ids from the sampled DataFrame
    discovery_pids = set(discovery_df['person_id'])
    
    # The confirmation set is the set difference of all IDs and discovery IDs
    all_pids = set(persons_df_copy['person_id'])
    confirmation_pids = all_pids - discovery_pids

    # --- 5. Cache the Results ---
    print(f"    - Caching split assignments to: {output_path.name}")
    with open(output_path, 'wb') as f:
        pickle.dump({
            'discovery_pids': discovery_pids,
            'confirmation_pids': confirmation_pids
        }, f)

# --- 6. Final Summary and Verification ---
total_persons = len(persons_df)
discovery_count = len(discovery_pids)
confirmation_count = len(confirmation_pids)

print("\n--- Split Summary ---")
print(f"Total eligible persons: {total_persons:,}")
print(f"Discovery set size:     {discovery_count:,} ({discovery_count / total_persons:.1%})")
print(f"Confirmation set size:  {confirmation_count:,} ({confirmation_count / total_persons:.1%})")
print(f"Overlap between sets:   {len(discovery_pids.intersection(confirmation_pids))}")
print("-----------------------")

# Sanity check
assert discovery_count + confirmation_count == total_persons, "Split counts do not sum to total persons!"

print("✅ Cell 5 complete: Discovery and confirmation sets are ready.")


# In[ ]:


# ──────────────────────────── Cell 6 ─────────────────────────────
# Time‑Varying & Context‑Aware Covariate Engine
# -----------------------------------------------------------------------------
# ✅ IMPORTANT: Function signature used downstream
#   get_covariates_at_index(
#       person_index_dates: pd.Series,  # index=person_id, values=index_date (datetime-like)
#       event_A_vid: int,
#       event_B_vid: int,
#       outcome_name: str,
#       lookback_days_smoking_bmi: int = 730,
#       lookback_days_util: int = 365,
#       lookback_days_cci: int = 730
#   ) -> pd.DataFrame with columns:
#       [person_id, index_date,
#        smoking_status_at_index, bmi_at_index,
#        deprivation_index, utilization_rate, cci_loo]
#
# Notes:
#   • CCI‑LOO excludes Charlson categories whose descendants intersect descendants of:
#       (A) event_A_vid, (B) event_B_vid, and (C) CONFIG.OUTCOME_DEFINITIONS[outcome_name].
#   • Requires artifacts created upstream:
#       - artifacts/charlson_events_df.parquet  (Cell 4a)
#       - artifacts/visits_df.parquet           (Cell 4a)
#   • Smoking/BMI are derived from "Derived" events in vocab/event_vocab_v1.parquet.
#   • This implementation MERGES ON ['person_id','index_date'] to avoid misalignment
#     when people contribute multiple episodes.
# -----------------------------------------------------------------------------

import pandas as pd
import numpy as np
from pathlib import Path
from functools import lru_cache

print("▶ Cell 6: Building the context‑aware covariate engine...")

# --- 1) Prerequisite checks --------------------------------------------------
required_globals = ['CONFIG', 'events_df', 'persons_df', 'client', 'WORKSPACE_CDR']
for _v in required_globals:
    if _v not in globals():
        raise NameError(f"❌ Prerequisite '{_v}' not found. Please run preceding cells first.")

from google.cloud import bigquery  # noqa: F401

ART_DIR = CONFIG.ARTIFACTS_DIR
SRC_DIR = CONFIG.SOURCE_CACHE_DIR

# --- 2) Load "Derived" covariate events (Smoking & BMI) ----------------------
# These are rolled‑up into events_df as feature_concept_id values coming from vocab/event_vocab_v1.parquet.
vocab_path = SRC_DIR / "event_vocab_v1.parquet"
if not vocab_path.exists():
    raise FileNotFoundError("Could not find `event_vocab_v1.parquet` in the source cache.")

vocab_df = pd.read_parquet(vocab_path)

# Keep only "Derived" rows and extract category/value from concept_name
derived_concepts = vocab_df[vocab_df['source_domain'] == 'Derived'].copy()
derived_concepts = derived_concepts[['concept_id', 'concept_name']].dropna()

# Build a helper to split "Category · Value" (robust to minor variants)
def _split_category_value(name: str):
    if not isinstance(name, str) or not name:
        return None, None
    # Primary split on "·", then fallbacks
    if '·' in name:
        parts = [p.strip() for p in name.split('·', 1)]
    elif '|' in name:
        parts = [p.strip() for p in name.split('|', 1)]
    elif ':' in name:
        parts = [p.strip() for p in name.split(':', 1)]
    elif ' - ' in name:
        parts = [p.strip() for p in name.split(' - ', 1)]
    else:
        # If no recognizable separator, try to infer known categories
        if name.lower().startswith('smoking'):
            return 'Smoking status', name.replace('Smoking', '').strip() or None
        if 'bmi' in name.lower():
            return 'BMI', name.replace('BMI', '').replace('bmi', '').strip() or None
        return None, None
    cat = parts[0] if len(parts) > 0 else None
    val = parts[1] if len(parts) > 1 else None
    return cat, val

# Map events_df → only derived concepts for smoking/BMI
derived_ids = set(derived_concepts['concept_id'].astype(int))
_cov = events_df[events_df['feature_concept_id'].isin(derived_ids)].copy()
_cov = _cov.merge(derived_concepts, left_on='feature_concept_id', right_on='concept_id', how='left')

_cat_vals = _cov['concept_name'].apply(_split_category_value)
_cov['category'] = _cat_vals.apply(lambda t: t[0])
_cov['value']    = _cat_vals.apply(lambda t: t[1])

# Normalize category label variants
_cov['category'] = _cov['category'].replace({
    'Smoking': 'Smoking status',
    'Smoking Status': 'Smoking status',
    'BMI category': 'BMI',
    'Body mass index': 'BMI'
})
# Keep only the covariates we care about
_cov = _cov[_cov['category'].isin(['Smoking status', 'BMI'])].copy()

_cov = _cov[['person_id', 'event_date', 'category', 'value']].copy()
_cov['event_date'] = pd.to_datetime(_cov['event_date'], errors='coerce')
_cov = _cov.dropna(subset=['person_id', 'event_date']).reset_index(drop=True)

print(f"  • Prepared Derived covariate events (Smoking/BMI): {_cov.shape[0]:,} rows")

# --- 3) Load Visits and Charlson artifacts (from new Cell 4a) ----------------
visits_path    = ART_DIR / "visits_df.parquet"
charlson_path  = ART_DIR / "charlson_events_df.parquet"

if visits_path.exists():
    visits_df = pd.read_parquet(visits_path)
    if 'visit_start_date' not in visits_df.columns:
        raise KeyError("`visits_df.parquet` must contain a 'visit_start_date' column.")
    visits_df['visit_start_date'] = pd.to_datetime(visits_df['visit_start_date'], errors='coerce')
    visits_df = visits_df.dropna(subset=['person_id', 'visit_start_date']).reset_index(drop=True)
    print(f"  • Loaded visits_df: {len(visits_df):,} visit rows")
else:
    print("  • WARNING: visits_df.parquet not found — utilization_rate will be 0 for all.")
    visits_df = pd.DataFrame(columns=['person_id', 'visit_start_date'])

if charlson_path.exists():
    charlson_events_df = pd.read_parquet(charlson_path)
    # Expect columns: person_id, event_date, charlson_condition_id
    required_charlson_cols = {'person_id', 'event_date', 'charlson_condition_id'}
    if not required_charlson_cols.issubset(charlson_events_df.columns):
        raise KeyError("`charlson_events_df.parquet` must contain 'person_id', 'event_date', 'charlson_condition_id'.")
    charlson_events_df['event_date'] = pd.to_datetime(charlson_events_df['event_date'], errors='coerce')
    charlson_events_df['charlson_condition_id'] = pd.to_numeric(
        charlson_events_df['charlson_condition_id'], errors='coerce'
    ).astype('Int64')
    charlson_events_df = charlson_events_df.dropna(subset=['person_id', 'event_date', 'charlson_condition_id']).reset_index(drop=True)
    print(f"  • Loaded charlson_events_df: {len(charlson_events_df):,} rows")
else:
    print("  • WARNING: charlson_events_df.parquet not found — cci_loo will be 0 for all.")
    charlson_events_df = pd.DataFrame(columns=['person_id', 'event_date', 'charlson_condition_id'])

# --- 4) Helpers: descendant lookups via concept_ancestor ---------------------
@lru_cache(maxsize=256)
def _descendants_for(ancestor_ids_tuple: tuple) -> set:
    """
    Return the set of descendant_concept_id for the given ancestor concept_ids.
    Cached to avoid repeated BigQuery hits.
    """
    ids = [int(x) for x in ancestor_ids_tuple if x is not None]
    if not ids:
        return set()
    sql = f"""
        SELECT DISTINCT descendant_concept_id
        FROM `{WORKSPACE_CDR}.concept_ancestor`
        WHERE ancestor_concept_id IN UNNEST(@ancestors)
    """
    job_config = bigquery.QueryJobConfig(
        query_parameters=[bigquery.ArrayQueryParameter("ancestors", "INT64", ids)]
    )
    df = client.query(sql, job_config=job_config).to_dataframe()
    if df.empty:
        return set()
    return set(df['descendant_concept_id'].dropna().astype(int).tolist())

@lru_cache(maxsize=256)
def _charlson_descendants_map(charlson_ids_tuple: tuple) -> dict:
    """
    Map each Charlson ancestor (charlson_condition_id) to the set of its descendants.
    """
    ids = [int(x) for x in charlson_ids_tuple if x is not None]
    if not ids:
        return {}
    sql = f"""
        SELECT ancestor_concept_id AS charlson_id, descendant_concept_id
        FROM `{WORKSPACE_CDR}.concept_ancestor`
        WHERE ancestor_concept_id IN UNNEST(@ancestors)
    """
    job_config = bigquery.QueryJobConfig(
        query_parameters=[bigquery.ArrayQueryParameter("ancestors", "INT64", ids)]
    )
    df = client.query(sql, job_config=job_config).to_dataframe()
    if df.empty:
        return {cid: set() for cid in ids}
    out = {}
    for cid, grp in df.groupby('charlson_id'):
        out[int(cid)] = set(grp['descendant_concept_id'].dropna().astype(int).tolist())
    # Ensure all requested ids present
    for cid in ids:
        out.setdefault(int(cid), set())
    return out

# --- 5) Main function --------------------------------------------------------
def get_covariates_at_index(person_index_dates: pd.Series,
                            event_A_vid: int,
                            event_B_vid: int,
                            outcome_name: str,
                            lookback_days_smoking_bmi: int = 730,
                            lookback_days_util: int = 365,
                            lookback_days_cci: int = 730) -> pd.DataFrame:
    """
    Compute context‑aware covariates for (person_id, index_date) pairs.

    NOTE (Patch C): All lookbacks are now ANCHORED AT TIME ZERO:
        anchor_date = index_date + CONFIG.LATENCY_DAYS

    Returns
    -------
    pd.DataFrame with columns:
        ['person_id', 'index_date',
         'smoking_status_at_index', 'bmi_at_index',
         'deprivation_index', 'utilization_rate', 'cci_loo']
    """
    # ---- Guard & normalize input -------------------------------------------
    if not isinstance(person_index_dates, pd.Series) or person_index_dates.empty:
        return pd.DataFrame(columns=[
            'person_id', 'index_date',
            'smoking_status_at_index', 'bmi_at_index',
            'deprivation_index', 'utilization_rate', 'cci_loo'
        ])

    index_df = person_index_dates.rename('index_date').reset_index()
    index_df.columns = ['person_id', 'index_date']
    index_df['index_date'] = pd.to_datetime(index_df['index_date'], errors='coerce')
    index_df = index_df.dropna(subset=['person_id', 'index_date']).copy()
    if index_df.empty:
        return pd.DataFrame(columns=[
            'person_id', 'index_date',
            'smoking_status_at_index', 'bmi_at_index',
            'deprivation_index', 'utilization_rate', 'cci_loo'
        ])

    # Time‑zero anchor for lookbacks
    latency_days_cfg = int(getattr(CONFIG, 'LATENCY_DAYS', 90))
    index_df['anchor_date'] = index_df['index_date'] + pd.Timedelta(days=latency_days_cfg)

    # Keep a working list of person IDs
    index_df['person_id'] = pd.to_numeric(index_df['person_id'], errors='coerce').astype('Int64')
    pid_list = index_df['person_id'].dropna().astype(int).unique().tolist()

    # ---- Start assembling result (we will drop anchor_date before return) ---
    result = index_df.copy()

    # ---- SES: deprivation_index (static from persons_df) -------------------
    if 'deprivation_index' in persons_df.columns:
        ses = persons_df[['person_id', 'deprivation_index']].copy()
        result = result.merge(ses, on='person_id', how='left')
        if result['deprivation_index'].notna().any():
            med = float(pd.to_numeric(result['deprivation_index'], errors='coerce').dropna().median())
            result['deprivation_index'] = pd.to_numeric(result['deprivation_index'], errors='coerce').fillna(med)
        else:
            result['deprivation_index'] = np.nan
    else:
        result['deprivation_index'] = np.nan

    # ---- Smoking & BMI (most recent within lookback, ANCHORED AT TIME ZERO) -
    if not _cov.empty:
        cov = index_df.merge(_cov[_cov['person_id'].isin(pid_list)], on='person_id', how='left')
        lb = pd.Timedelta(days=int(lookback_days_smoking_bmi))
        valid = (
            cov['event_date'].notna() &
            (cov['event_date'] <= cov['anchor_date']) &
            (cov['event_date'] >= (cov['anchor_date'] - lb))
        )
        cov_valid = cov.loc[valid, ['person_id', 'index_date', 'anchor_date', 'category', 'value', 'event_date']].copy()
        if not cov_valid.empty:
            cov_valid.sort_values(['person_id', 'index_date', 'category', 'event_date'], inplace=True)
            last_vals = cov_valid.drop_duplicates(subset=['person_id', 'index_date', 'category'], keep='last')
            piv = last_vals.pivot_table(index=['person_id', 'index_date'], columns='category', values='value', aggfunc='first')
            ren = {}
            for c in piv.columns:
                cl = str(c).strip().lower()
                if cl.startswith('smoking'):
                    ren[c] = 'smoking_status_at_index'
                elif cl.startswith('bmi'):
                    ren[c] = 'bmi_at_index'
            piv = piv.rename(columns=ren).reset_index()
            result = result.merge(piv, on=['person_id', 'index_date'], how='left')
        else:
            result['smoking_status_at_index'] = np.nan
            result['bmi_at_index'] = np.nan
    else:
        result['smoking_status_at_index'] = np.nan
        result['bmi_at_index'] = np.nan

    # Default to "Unknown" for missing categorical covariates
    result['smoking_status_at_index'] = result['smoking_status_at_index'].astype('string').fillna("Unknown")
    result['bmi_at_index']            = result['bmi_at_index'].astype('string').fillna("Unknown")

    # ---- Utilization: distinct visits in prior 365 days (ANCHOR = time zero)
    if not visits_df.empty:
        vis = index_df.merge(visits_df[visits_df['person_id'].isin(pid_list)], on='person_id', how='left')
        lb_u = pd.Timedelta(days=int(lookback_days_util))
        mask_u = (
            vis['visit_start_date'].notna() &
            (vis['visit_start_date'] <= vis['anchor_date']) &
            (vis['visit_start_date'] >= (vis['anchor_date'] - lb_u))
        )
        vis_valid = vis.loc[mask_u, ['person_id', 'index_date', 'visit_start_date']].drop_duplicates()
        if not vis_valid.empty:
            util = (
                vis_valid
                .groupby(['person_id', 'index_date'], as_index=False)['visit_start_date']
                .nunique()
                .rename(columns={'visit_start_date': 'utilization_rate'})
            )
            result = result.merge(util, on=['person_id', 'index_date'], how='left')
        else:
            result['utilization_rate'] = 0
    else:
        result['utilization_rate'] = 0
    result['utilization_rate'] = pd.to_numeric(result['utilization_rate'], errors='coerce').fillna(0).astype('Int64')

    # ---- CCI‑LOO: prior 730 days (ANCHOR = time zero) with exclusions -------
    outcome_ancestors = CONFIG.OUTCOME_DEFINITIONS.get(outcome_name, [])
    exclude_desc = set()
    try: exclude_desc |= _descendants_for(tuple({int(event_A_vid)}))
    except Exception: pass
    try: exclude_desc |= _descendants_for(tuple({int(event_B_vid)}))
    except Exception: pass
    try:
        if outcome_ancestors:
            exclude_desc |= _descendants_for(tuple(sorted({int(x) for x in outcome_ancestors})))
    except Exception:
        pass

    if not charlson_events_df.empty:
        ch_subset = charlson_events_df[charlson_events_df['person_id'].isin(pid_list)]
        observed_charlson_ids = sorted(ch_subset['charlson_condition_id'].dropna().astype(int).unique().tolist())
        ch_map = _charlson_descendants_map(tuple(observed_charlson_ids)) if observed_charlson_ids else {}
        exclude_charlson_ids = {cid for cid, descs in ch_map.items() if exclude_desc and (descs & exclude_desc)}
        if exclude_charlson_ids:
            ch_subset = ch_subset[~ch_subset['charlson_condition_id'].isin(exclude_charlson_ids)].copy()

        if ch_subset.empty:
            result['cci_loo'] = 0
        else:
            lb_c = pd.Timedelta(days=int(lookback_days_cci))
            m = index_df.merge(
                ch_subset[['person_id', 'event_date', 'charlson_condition_id']],
                on='person_id', how='left'
            )
            mask_c = (
                m['event_date'].notna() &
                (m['event_date'] <= m['anchor_date']) &
                (m['event_date'] >= (m['anchor_date'] - lb_c))
            )
            m = m.loc[mask_c, ['person_id', 'index_date', 'charlson_condition_id']].drop_duplicates()
            if not m.empty:
                cci = (
                    m.groupby(['person_id', 'index_date'], as_index=False)['charlson_condition_id']
                     .nunique()
                     .rename(columns={'charlson_condition_id': 'cci_loo'})
                )
                result = result.merge(cci, on=['person_id', 'index_date'], how='left')
            else:
                result['cci_loo'] = 0
    else:
        result['cci_loo'] = 0
    result['cci_loo'] = pd.to_numeric(result['cci_loo'], errors='coerce').fillna(0).astype('Int64')

    # --- Finalize ------------------------------------------------------------
    final_cols = [
        'person_id', 'index_date',
        'smoking_status_at_index', 'bmi_at_index',
        'deprivation_index', 'utilization_rate', 'cci_loo'
    ]
    out = result[final_cols].copy()
    # dtype normalization
    out['index_date'] = pd.to_datetime(out['index_date'], errors='coerce')
    out['person_id'] = pd.to_numeric(out['person_id'], errors='coerce').astype('Int64')
    return out


# --- 6) Tiny demonstration (safe, optional) ----------------------------------
print("  • Engine defined. Running a small demonstration (will skip if concepts unavailable)...")
try:
    # Choose two non‑Derived concepts for demo A/B (random but deterministic)
    non_derived = vocab_df[vocab_df['source_domain'] != 'Derived']['concept_id'].dropna().astype(int)
    if len(non_derived) >= 2:
        demo_A = int(non_derived.sample(1, random_state=CONFIG.RNG_SEED).iloc[0])
        demo_B = int(non_derived.sample(1, random_state=CONFIG.RNG_SEED + 1).iloc[0])
    else:
        raise ValueError("Not enough non‑Derived concepts for demo.")

    demo_outcome = 'myocardial_infarction' if 'myocardial_infarction' in CONFIG.OUTCOME_DEFINITIONS else list(CONFIG.OUTCOME_DEFINITIONS.keys())[0]
    sample_n = min(5, len(persons_df))
    if sample_n > 0:
        sample_pids = persons_df.sample(sample_n, random_state=CONFIG.RNG_SEED)['person_id'].astype('Int64')
        sample_dates = pd.to_datetime(['2018-01-01', '2019-06-15', '2020-03-01', '2017-11-20', '2021-01-01'])[:sample_n]
        demo_series = pd.Series(sample_dates.values, index=sample_pids.values)

        demo_res = get_covariates_at_index(
            demo_series, event_A_vid=int(demo_A), event_B_vid=int(demo_B),
            outcome_name=str(demo_outcome)
        )
        print("\n--- Demonstration Output (head) ---")
        display(demo_res.head())
        print("-----------------------------------")
except Exception as e:
    print(f"    - Demo skipped: {e}")

print("✅ Cell 6 complete: Context‑aware covariate engine is ready.")


# In[ ]:


# ──────────────────────────── Cell 7 (REVISED • GAP-AWARE) ─────────────────────────────
# Resumable Episode Construction Engine (multi-interval observation windows)
# ---------------------------------------------------------------------------------------
#

import pandas as pd
import numpy as np
from pathlib import Path
from typing import Dict, Set
import time

print("▶ Cell 7: Defining the Resumable Episode Construction Engine...")

# --- 1) Prerequisite checks --------------------------------------------------
required_globals = [
    'CONFIG', 'log', 'persons_df', 'events_df', 'get_covariates_at_index'
]
for _v in required_globals:
    if _v not in globals():
        raise NameError(f"❌ Prerequisite '{_v}' not found. Please run the preceding cells first.")

# Ensure multi-interval observation periods are available (from revised Cell 2)
if 'observation_periods_df' not in globals():
    _op_path = CONFIG.ARTIFACTS_DIR / "observation_periods_df.parquet"
    if _op_path.exists():
        observation_periods_df = pd.read_parquet(_op_path)
        print(f"  • Loaded observation periods from cache: {_op_path.name} ({len(observation_periods_df):,} rows)")
    else:
        raise FileNotFoundError(
            "❌ `observation_periods_df.parquet` not found. Please run the revised Cell 2 that builds "
            "this artifact (all intervals per person) before running Cell 7."
        )

# Normalize/validate dtypes for observation intervals
for col in ('obs_start_date', 'obs_end_date'):
    if not pd.api.types.is_datetime64_any_dtype(observation_periods_df[col]):
        observation_periods_df[col] = pd.to_datetime(observation_periods_df[col], errors='coerce')
# Drop obviously invalid intervals (start > end or NaT)
_ob_before = len(observation_periods_df)
observation_periods_df = observation_periods_df.dropna(subset=['person_id', 'obs_start_date', 'obs_end_date'])
observation_periods_df = observation_periods_df[observation_periods_df['obs_end_date'] >= observation_periods_df['obs_start_date']].copy()
_ob_after = len(observation_periods_df)
if _ob_after < _ob_before:
    print(f"  • Cleaned {(_ob_before - _ob_after):,} invalid observation-period rows (start/end issues).")

# --- 2) Core engine ----------------------------------------------------------
def build_episodes_for_sequence(
    event_A_vid: int,
    event_B_vid: int,
    person_ids: Set[int]
) -> Dict[str, pd.DataFrame]:
    """
    Construct episodes for a given A→B sequence using multi-interval observation windows.

    Args
    ----
    event_A_vid : int
        Curated concept_id for the first event (A).
    event_B_vid : int
        Curated concept_id for the second event (B).
    person_ids  : Set[int]
        Set of people (e.g., discovery or confirmation cohort) to include.

    Returns
    -------
    Dict[str, pd.DataFrame]
        Keys: 'A_to_B', 'B_no_prior_A', 'B_to_A'.
        Each DataFrame includes:
          person_id, index_date, time_zero, censor_date, cohort_label,
          age_at_index, calendar_year_of_index, smoking_status_at_index, bmi_at_index
    """

    # ---- inner helper: finalize one cohort with interval validation ----
    def _finalize_cohort_with_intervals(candidates: pd.DataFrame, cohort_label: str) -> pd.DataFrame:
        """
        Validate that index_date and time_zero lie within the SAME observation interval,
        compute censor_date from that interval, attach covariates, and return final schema.
        """
        if candidates.empty:
            return pd.DataFrame(columns=[
                'person_id','index_date','time_zero','censor_date','cohort_label',
                'age_at_index','calendar_year_of_index','smoking_status_at_index','bmi_at_index'
            ])

        df = candidates.copy()
        # Normalize date dtype & drop invalid
        df['index_date'] = pd.to_datetime(df['index_date'], errors='coerce')
        df = df.dropna(subset=['person_id', 'index_date'])
        if df.empty:
            return pd.DataFrame(columns=[
                'person_id','index_date','time_zero','censor_date','cohort_label',
                'age_at_index','calendar_year_of_index','smoking_status_at_index','bmi_at_index'
            ])

        # Post-latency time zero
        latency_delta = pd.Timedelta(days=int(CONFIG.LATENCY_DAYS))
        df['time_zero'] = df['index_date'] + latency_delta

        # Merge with observation intervals for *these* persons only (speed)
        op_sub = observation_periods_df[
            observation_periods_df['person_id'].isin(df['person_id'].unique())
        ].copy()

        merged = df.merge(op_sub, on='person_id', how='inner')

        # Keep only rows where BOTH index_date and time_zero are inside the SAME interval
        within_index = (merged['index_date'] >= merged['obs_start_date']) & (merged['index_date'] <= merged['obs_end_date'])
        within_tz    = (merged['time_zero']  >= merged['obs_start_date']) & (merged['time_zero']  <= merged['obs_end_date'])
        valid = merged[within_index & within_tz].copy()

        if valid.empty:
            return pd.DataFrame(columns=[
                'person_id','index_date','time_zero','censor_date','cohort_label',
                'age_at_index','calendar_year_of_index','smoking_status_at_index','bmi_at_index'
            ])

        # Enforce minimum lookback for selected labels
        LOOKBACK_LABELS = {'B_no_prior_A', 'A_no_B', 'Baseline'}
        if cohort_label in LOOKBACK_LABELS:
            lookback_delta = pd.Timedelta(days=int(getattr(CONFIG, 'RISKSET_MIN_LOOKBACK_DAYS', 1825)))
            # The observation period must have started at least 'lookback_delta' days before the index date.
            had_sufficient_lookback = (valid['obs_start_date'] <= (valid['index_date'] - lookback_delta))
            valid = valid.loc[had_sufficient_lookback].copy()

            # If no episodes remain after this filter, return an empty frame.
            if valid.empty:
                return pd.DataFrame(columns=[
                    'person_id','index_date','time_zero','censor_date','cohort_label',
                    'age_at_index','calendar_year_of_index','smoking_status_at_index','bmi_at_index'
                ])

        # Censor at the end of the containing interval
        valid['censor_date'] = valid['obs_end_date']

        # If (person_id, index_date) appears in multiple overlapping intervals (rare), keep the one with the earliest censor_date.
        valid.sort_values(['person_id', 'index_date', 'censor_date'], inplace=True)
        valid = valid.drop_duplicates(subset=['person_id', 'index_date'], keep='first')

        # Attach static covariates
        valid = valid.merge(
            persons_df[['person_id', 'year_of_birth']],
            on='person_id', how='left'
        )
        valid['age_at_index'] = (valid['index_date'].dt.year - valid['year_of_birth']).astype('Int64')
        valid['calendar_year_of_index'] = valid['index_date'].dt.year.astype('Int64')

        # Final shape
        valid['cohort_label'] = cohort_label
        final_cols = [
            'person_id', 'index_date', 'time_zero', 'censor_date', 'cohort_label',
            'age_at_index', 'calendar_year_of_index'
        ]
        return valid[final_cols].reset_index(drop=True)

    # ---- main logic: build candidate episodes from event history ----
    # Subset events to these people & the two concept ids of interest
    ev = events_df[
        (events_df['person_id'].isin(person_ids)) &
        (events_df['feature_concept_id'].isin([event_A_vid, event_B_vid]))
    ].copy()

    # First observed dates for A and B
    first_A = (
        ev[ev['feature_concept_id'] == event_A_vid]
        .groupby('person_id', sort=False)['event_date'].min()
        .rename('date_A')
    )
    first_B = (
        ev[ev['feature_concept_id'] == event_B_vid]
        .groupby('person_id', sort=False)['event_date'].min()
        .rename('date_B')
    )

    merged_dates = pd.merge(
        first_A.to_frame(), first_B.to_frame(),
        left_index=True, right_index=True, how='outer'
    ).reset_index().rename(columns={'index': 'person_id'})

    # SAME_DAY_POLICY + max gap
    policy = str(getattr(CONFIG, 'SAME_DAY_POLICY', 'exclude')).lower()
    max_gap = pd.Timedelta(days=int(getattr(CONFIG, 'EPISODE_MAX_GAP_DAYS', 1825)))

    # build masks
    if policy == 'allow':
        # A→B: B >= A (allow same day), and within max gap
        a_to_b_mask = (
            merged_dates['date_A'].notna() & merged_dates['date_B'].notna() &
            (merged_dates['date_B'] >= merged_dates['date_A']) &
            ((merged_dates['date_B'] - merged_dates['date_A']) <= max_gap)
        )
        # B with no prior A: A is missing or A >= B (i.e., no prior A strictly before B)
        b_no_a_mask = (
            merged_dates['date_B'].notna() &
            (merged_dates['date_A'].isna() | (merged_dates['date_A'] >= merged_dates['date_B']))
        )
        # B→A: A >= B and within max gap (allow same day)
        b_to_a_mask = (
            merged_dates['date_A'].notna() & merged_dates['date_B'].notna() &
            (merged_dates['date_A'] >= merged_dates['date_B']) &
            ((merged_dates['date_A'] - merged_dates['date_B']) <= max_gap)
        )

    elif policy == 'require':
        # same-day only for ordered sequences
        a_to_b_mask = (
            merged_dates['date_A'].notna() & merged_dates['date_B'].notna() &
            (merged_dates['date_B'] == merged_dates['date_A'])
        )
        b_no_a_mask = (merged_dates['date_B'].notna() & merged_dates['date_A'].isna())
        b_to_a_mask = (
            merged_dates['date_A'].notna() & merged_dates['date_B'].notna() &
            (merged_dates['date_A'] == merged_dates['date_B'])
        )

    else:  # 'exclude' (default): require strict order
        a_to_b_mask = (
            merged_dates['date_A'].notna() & merged_dates['date_B'].notna() &
            (merged_dates['date_B'] > merged_dates['date_A']) &
            ((merged_dates['date_B'] - merged_dates['date_A']) <= max_gap)
        )
        b_no_a_mask = (
            merged_dates['date_B'].notna() &
            (merged_dates['date_A'].isna() |
             (merged_dates['date_A'].notna() & (merged_dates['date_A'] > merged_dates['date_B'])))
        )
        b_to_a_mask = (
            merged_dates['date_A'].notna() & merged_dates['date_B'].notna() &
            (merged_dates['date_A'] > merged_dates['date_B']) &
            ((merged_dates['date_A'] - merged_dates['date_B']) <= max_gap)
        )

    # Candidates: (person_id, index_date)
    a_to_b_cand = (
        merged_dates.loc[a_to_b_mask, ['person_id', 'date_B']]
        .rename(columns={'date_B': 'index_date'})
        .dropna(subset=['index_date'])
        .reset_index(drop=True)
    )
    b_no_a_cand = (
        merged_dates.loc[b_no_a_mask, ['person_id', 'date_B']]
        .rename(columns={'date_B': 'index_date'})
        .dropna(subset=['index_date'])
        .reset_index(drop=True)
    )
    b_to_a_cand = (
        merged_dates.loc[b_to_a_mask, ['person_id', 'date_A']]
        .rename(columns={'date_A': 'index_date'})
        .dropna(subset=['index_date'])
        .reset_index(drop=True)
    )

    # ── Define "subsequent B" in terms of SAME_DAY_POLICY ─────────────────────
    if policy == 'allow':      # B >= A counts as subsequent
        _subseq = (merged_dates['date_B'] >= merged_dates['date_A'])
    elif policy == 'require':  # only same day counts as subsequent
        _subseq = (merged_dates['date_B'] == merged_dates['date_A'])
    else:                      # 'exclude' -> strictly after
        _subseq = (merged_dates['date_B'] > merged_dates['date_A'])

    b_after_a = (
        merged_dates['date_A'].notna() & merged_dates['date_B'].notna() & _subseq
    )
    within_gap = (merged_dates['date_B'] - merged_dates['date_A']) <= max_gap

    # Tertiary: A with no subsequent B within the configured gap
    a_no_b_mask = merged_dates['date_A'].notna() & (~(b_after_a & within_gap).fillna(False))
    a_no_b_cand = (
        merged_dates.loc[a_no_b_mask, ['person_id', 'date_A']]
        .rename(columns={'date_A': 'index_date'})
        .dropna(subset=['index_date'])
        .reset_index(drop=True)
    )

    # Quaternary: Baseline episode (one per person; OP-aware; adequate lookback)
    def _baseline_candidates(pid_set: Set[int]) -> pd.DataFrame:
        op_sub = observation_periods_df[observation_periods_df['person_id'].isin(pid_set)].copy()
        lb  = pd.Timedelta(days=int(getattr(CONFIG, 'RISKSET_MIN_LOOKBACK_DAYS', 1825)))
        lat = pd.Timedelta(days=int(getattr(CONFIG, 'LATENCY_DAYS', 90)))
        op_sub['candidate_index'] = op_sub['obs_start_date'] + lb
        # need time_zero inside same interval
        ok = (op_sub['candidate_index'] + lat <= op_sub['obs_end_date'])
        base = (
            op_sub.loc[ok, ['person_id', 'candidate_index']]
            .sort_values(['person_id', 'candidate_index'])
            .drop_duplicates('person_id')
            .rename(columns={'candidate_index': 'index_date'})
            .reset_index(drop=True)
        )
        return base

    baseline_cand = _baseline_candidates(person_ids)

    # Finalize using your interval validator (enforce lookback for these labels too)
    results = {
        'A_to_B':       _finalize_cohort_with_intervals(a_to_b_cand, 'A_to_B'),
        'B_no_prior_A': _finalize_cohort_with_intervals(b_no_a_cand, 'B_no_prior_A'),
        'B_to_A':       _finalize_cohort_with_intervals(b_to_a_cand, 'B_to_A'),
        'A_no_B':       _finalize_cohort_with_intervals(a_no_b_cand, 'A_no_B'),
        'Baseline':     _finalize_cohort_with_intervals(baseline_cand, 'Baseline'),
    }
    return results

# --- 3) Optional demonstration (safe; tiny sample) ---------------------------
print("  • Engine defined. Running a small demonstration with a sample sequence...")

try:
    demo_vocab = pd.read_parquet(CONFIG.SOURCE_CACHE_DIR / "vocab" / "event_vocab_v1.parquet")
    # Replace these lines with concepts that exist in your vocab (example: 'Hyperlipidemia' → 'atorvastatin')
    demo_A = demo_vocab.loc[demo_vocab['concept_name'] == 'Hyperlipidemia', 'concept_id'].iloc[0]
    demo_B = demo_vocab.loc[demo_vocab['concept_name'] == 'atorvastatin',   'concept_id'].iloc[0]

    # Sample a modest set of people for speed
    demo_pids = set(persons_df.sample(n=min(10000, len(persons_df)), random_state=CONFIG.RNG_SEED)['person_id'])

    t0 = time.perf_counter()
    demo_eps = build_episodes_for_sequence(int(demo_A), int(demo_B), demo_pids)
    dt = time.perf_counter() - t0

    print(f"    - Demo finished in {dt:.2f}s (SAME_DAY_POLICY='{CONFIG.SAME_DAY_POLICY}', LATENCY={CONFIG.LATENCY_DAYS}d).")
    for label, df in demo_eps.items():
        print(f"\nCohort: {label}")
        print(f"  • Episodes: {len(df):,}")
        if not df.empty:
            display(df.head(3))
    print("------------------------------------------------")

except (IndexError, FileNotFoundError, KeyError) as e:
    print(f"    - WARNING: Demo skipped (could not resolve demo concepts or vocab). Reason: {e}")

print("✅ Cell 7 complete: Episode construction engine is ready.")


# In[ ]:


# ──────────────────────────── Cell 8 ─────────────────────────
# Sequence Mining and Outcome‑aware Episode Generation (Discovery Set) — Interleaved across outcomes
# with optional per‑outcome quotas and full resumability.
#
# New features:
#   • Round‑robin / random interleaving across outcomes.
#   • Quotas for COMPLETED (sequence, outcome) pairs per outcome (per run), without losing resumability.
#   • Defaults remain "process everything", but interleaved fairly.
#
# Env overrides (all optional):
#   - CELL8_SCHEDULER_MODE:           "round_robin" (default) | "random"
#   - CELL8_SHUFFLE_SEQUENCES:        "1" (default) | "0"
#   - CELL8_QUOTA_PER_OUTCOME:        integer cap per outcome (uniform)
#   - CELL8_QUOTA_PER_OUTCOME_JSON:   JSON dict of {outcome: cap}, e.g. {"dementia":25,"myocardial_infarction":100}
#
# Outputs (unchanged):
#   artifacts/discovery_episodes/seq_{sequence_idx}__oc_{outcome}.parquet
#   artifacts/cell_8_checkpoint.pkl                   (attempted pair keys)
#   artifacts/sequence_map_df.parquet                 (mined frequent sequences)

import os
import pickle
import time
import itertools
import json
import random
from collections import Counter, deque
from pathlib import Path

import numpy as np
import pandas as pd
from sklearn.linear_model import LogisticRegression

print("▶ Cell 8: Mining sequences and generating episodes interleaved across outcomes...")

# ── 1) Prereqs & paths --------------------------------------------------------
required_globals = [
    'CONFIG', 'log', 'discovery_pids', 'events_df', 'persons_df',
    'build_episodes_for_sequence',        # from Cell 7
    'get_covariates_at_index'             # upgraded in Cell 6
]
for _name in required_globals:
    if _name not in globals():
        raise NameError(f"❌ Prerequisite '{_name}' not found. Please run the preceding cells first.")

ART_DIR = CONFIG.ARTIFACTS_DIR
sequence_map_path   = ART_DIR / "sequence_map_df.parquet"
episodes_output_dir = ART_DIR / "discovery_episodes"
episodes_output_dir.mkdir(exist_ok=True, parents=True)
checkpoint_path     = ART_DIR / "cell_8_checkpoint.pkl"

OUTCOMES = list(CONFIG.OUTCOME_DEFINITIONS.keys())

# ── 2) Mine frequent A→B sequences on discovery set (cache if available) -----
if sequence_map_path.exists():
    sequence_map_df = pd.read_parquet(sequence_map_path)
    print(f"  • Loaded cached frequent sequence map: {sequence_map_path.name} ({len(sequence_map_df):,} sequences).")
else:
    print("  • Mining frequent ordered pairs (A→B) in the discovery set...")
    t_mine_start = time.perf_counter()

    # Concept ids that are *not* Derived (everything eligible for mining)
    non_derived_ids = (
        pd.to_numeric(
            vocab_df.loc[vocab_df['source_domain'] != 'Derived', 'concept_id'],
            errors='coerce'
        )
        .dropna().astype('int64')
    )
    excluded_derived_ids = (
        pd.to_numeric(
            vocab_df.loc[vocab_df['source_domain'] == 'Derived', 'concept_id'],
            errors='coerce'
        )
        .dropna().astype('int64')
    )

    # Optional hard‑guard in case the 8 derived ids were patched but not in vocab yet
    derived_id_fallback = pd.Index([9000001,9000002,9000003,9000004,9000005,9000006,9000007,9000008], dtype='int64')
    excluded_derived_ids = pd.Index(excluded_derived_ids).union(derived_id_fallback)
    non_derived_ids = pd.Index(non_derived_ids).difference(excluded_derived_ids)

    print(f"Excluding {len(excluded_derived_ids)} 'Derived' concept_id(s) from mining.")

    # Now build discovery_events *only from non‑Derived* concepts
    discovery_events = events_df[
        (events_df['person_id'].isin(discovery_pids)) &
        (events_df['feature_concept_id'].isin(non_derived_ids))
    ].copy()

    # For each person, take unique events in their chronological order
    print("    - Building patient timelines...")
    patient_timelines = (
        discovery_events
        .sort_values(['person_id', 'event_date'])
        .groupby('person_id', sort=False)['feature_concept_id']
        .apply(lambda x: list(dict.fromkeys(x)))  # de‑duplicate preserving order
    )

    # Count all ordered pairs that occur within each timeline
    print("    - Counting ordered pairs across timelines...")
    sequence_counts = Counter()
    iterator = patient_timelines
    if 'tqdm' in globals() and (tqdm is not None):
        iterator = tqdm(patient_timelines, desc="Mining Sequences", unit="patient")

    for timeline in iterator:
        if len(timeline) >= 2:
            for pair in itertools.combinations(timeline, 2):
                sequence_counts[pair] += 1

    # Keep sequences meeting minimum support
    frequent_sequences = [
        seq for seq, count in sequence_counts.items()
        if count >= CONFIG.MIN_SEQUENCE_SUPPORT
    ]
    print(f"    - Found {len(frequent_sequences):,} sequences with support ≥ {CONFIG.MIN_SEQUENCE_SUPPORT}.")

    sequence_map_df = pd.DataFrame(frequent_sequences, columns=['event_A_vid', 'event_B_vid'])
    sequence_map_df.index.name = "sequence_idx"
    sequence_map_df.reset_index(inplace=True)
    sequence_map_df.to_parquet(sequence_map_path, index=False)
    print(f"    - Cached sequence map to: {sequence_map_path.name}")
    print(f"  • Mining completed in {(time.perf_counter() - t_mine_start):.1f}s.")

# ── 3) Resume support: load processed (sequence, outcome) pairs ---------------
if checkpoint_path.exists():
    with open(checkpoint_path, 'rb') as f:
        processed_pairs = pickle.load(f)
    if not isinstance(processed_pairs, set):
        processed_pairs = set(processed_pairs)
    print(f"  • Resuming. {len(processed_pairs):,} (sequence, outcome) pairs already attempted.")
else:
    processed_pairs = set()
    print("  • Starting fresh. No checkpoint found.")

# ── 4) SMD calculator (unchanged) --------------------------------------------
def calculate_smd(df, group_col, weight_col=None):
    smds = {}
    categorical_covs = ['sex_at_birth', 'race_ethnicity', 'smoking_status_at_index', 'bmi_at_index']
    numeric_covs = [c for c in CONFIG.COVARIATES_FOR_ADJUSTMENT if c not in categorical_covs]

    have_cats = [c for c in categorical_covs if c in df.columns]
    have_nums = [c for c in numeric_covs if c in df.columns]

    df_enc = df.copy()
    if have_cats:
        df_enc = pd.get_dummies(df_enc, columns=have_cats, dummy_na=True, drop_first=True)

    cov_cols = have_nums + [c for c in df_enc.columns if any(cc + '_' in c for cc in categorical_covs)]
    group1 = df_enc[df_enc[group_col] == 1]
    group0 = df_enc[df_enc[group_col] == 0]

    for cov in cov_cols:
        if cov not in group1.columns or cov not in group0.columns:
            continue

        if weight_col and (weight_col in group1.columns) and (weight_col in group0.columns):
            w1, w0 = group1[weight_col].astype(float), group0[weight_col].astype(float)
            x1, x0 = group1[cov].astype(float), group0[cov].astype(float)
            if (w1.sum() <= 0) or (w0.sum() <= 0):
                smds[cov] = 0.0
                continue
            mean1 = np.average(x1, weights=w1)
            mean0 = np.average(x0, weights=w0)
            var1 = np.average((x1 - mean1) ** 2, weights=w1)
            var0 = np.average((x0 - mean0) ** 2, weights=w0)
        else:
            x1, x0 = group1[cov].astype(float), group0[cov].astype(float)
            mean1, mean0 = x1.mean(), x0.mean()
            var1, var0 = x1.var(), x0.var()

        pooled = np.sqrt((var1 + var0) / 2.0) if (var1 >= 0 and var0 >= 0) else 0.0
        smds[cov] = float((mean1 - mean0) / pooled) if pooled > 0 else 0.0

    return smds

def _json_default(o):
    try:
        if pd.isna(o):
            return None
    except Exception:
        pass
    if isinstance(o, (np.floating,)):
        return float(o)
    if isinstance(o, (np.integer,)):
        return int(o)
    return o

# ── 5) NEW: Scheduler + quota configuration ----------------------------------
SCHEDULER_MODE = "round_robin"
SHUFFLE_SEQS   = "1"
UNIFORM_QUOTA  = 1500
QUOTA_JSON     = ""

def _parse_quota_map(outcomes):
    """
    Returns dict {outcome: None|int}. None => unlimited for that outcome.
    Priority: JSON map > uniform.
    """
    quotas = {oc: None for oc in outcomes}
    if QUOTA_JSON:
        try:
            data = json.loads(QUOTA_JSON)
            for oc in outcomes:
                v = data.get(oc, None)
                if v is not None:
                    quotas[oc] = max(0, int(v))
        except Exception as e:
            print(f"  • WARNING: Could not parse CELL8_QUOTA_PER_OUTCOME_JSON ({e}); ignoring.")
    elif UNIFORM_QUOTA:
        try:
            q = max(0, int(UNIFORM_QUOTA))
            quotas = {oc: q for oc in outcomes}
        except Exception as e:
            print(f"  • WARNING: Could not parse CELL8_QUOTA_PER_OUTCOME ({e}); ignoring.")
    return quotas

QUOTAS = _parse_quota_map(OUTCOMES)

# Determine what's ALREADY completed from saved episode files
done_by_outcome = {oc: set() for oc in OUTCOMES}
for p in episodes_output_dir.glob("seq_*__oc_*.parquet"):
    stem = p.stem  # e.g., "seq_12__oc_dementia"
    try:
        seq_idx = int(stem.split('_')[1])
        oc = stem.split('__oc_')[1]
        if oc in done_by_outcome:
            done_by_outcome[oc].add(seq_idx)
    except Exception:
        continue

# Build per‑outcome queues of pending sequence indices (respecting processed_pairs)
seq_order = sequence_map_df['sequence_idx'].astype(int).tolist()
if SHUFFLE_SEQS:
    rnd = random.Random(int(getattr(CONFIG, 'RNG_SEED', 42)) + 1003)
    rnd.shuffle(seq_order)

queues = {}
remaining_needed = {}
for oc in OUTCOMES:
    pending = [
        seq_idx for seq_idx in seq_order
        if f"{seq_idx}::{oc}" not in processed_pairs       # not attempted yet
    ]
    queues[oc] = deque(pending)
    if QUOTAS[oc] is None:
        remaining_needed[oc] = None  # unlimited
    else:
        remaining_needed[oc] = max(0, QUOTAS[oc] - len(done_by_outcome[oc]))

print("\n--- Cell 8 Scheduler Summary ---")
print(f"Mode: {SCHEDULER_MODE} | Shuffle sequences: {SHUFFLE_SEQS}")
for oc in OUTCOMES:
    target = "∞" if QUOTAS[oc] is None else QUOTAS[oc]
    done   = len(done_by_outcome[oc])
    need   = "∞" if remaining_needed[oc] is None else remaining_needed[oc]
    qlen   = len(queues[oc])
    print(f"  • {oc:>24}: target={target}, done_now={done}, need_this_run={need}, queue_pending={qlen}")
print("----------------------------------------------------\n")

# ── 6) Helper: process a single (sequence, outcome) pair ---------------------
def _process_pair(seq_idx: int, outcome_name: str) -> bool:
    """
    Runs the original pair processing for (sequence_idx, outcome).
    Returns True iff an episode parquet was written (i.e., 'completed'),
    False if skipped (e.g., support < MIN_SEQUENCE_SUPPORT) or errored.
    Always records the attempt in the checkpoint.
    """
    pair_key = f"{seq_idx}::{outcome_name}"
    if pair_key in processed_pairs:
        return False  # already attempted in a prior run

    try:
        row = sequence_map_df.set_index('sequence_idx').loc[seq_idx]
        event_A, event_B = int(row['event_A_vid']), int(row['event_B_vid'])

        # a) Build OP‑aware episodes for this sequence (discovery cohort)
        episode_dict = build_episodes_for_sequence(event_A, event_B, discovery_pids)
        seq_df      = episode_dict.get('A_to_B',       pd.DataFrame()).copy()
        comp_df     = episode_dict.get('B_no_prior_A', pd.DataFrame()).copy()
        reverse_df  = episode_dict.get('B_to_A',       pd.DataFrame()).copy()
        a_no_b_df   = episode_dict.get('A_no_B',       pd.DataFrame()).copy()
        baseline_df = episode_dict.get('Baseline',     pd.DataFrame()).copy()

        # Require adequate size for primary comparison
        if len(seq_df) < CONFIG.MIN_SEQUENCE_SUPPORT or len(comp_df) < CONFIG.MIN_SEQUENCE_SUPPORT:
            return False

        # b) Outcome‑specific covariates for all (person_id, index_date)
        all_eps = pd.concat(
            [seq_df[['person_id','index_date']],
             comp_df[['person_id','index_date']],
             reverse_df[['person_id','index_date']],
             a_no_b_df[['person_id','index_date']],
             baseline_df[['person_id','index_date']]],
            ignore_index=True
        ).drop_duplicates()
        idx_series = all_eps.set_index('person_id')['index_date']  # duplicate index allowed
        cov_df = get_covariates_at_index(idx_series, event_A, event_B, outcome_name)
        if 'index_date' in cov_df.columns:
            cov_df['index_date'] = pd.to_datetime(cov_df['index_date'], errors='coerce')

        # Drop overlapping columns if any, then merge covariates
        for _df in (seq_df, comp_df, reverse_df, a_no_b_df, baseline_df):
            for c in ['smoking_status_at_index','bmi_at_index','deprivation_index','utilization_rate','cci_loo']:
                if c in _df.columns:
                    _df.drop(columns=[c], inplace=True)
        merge_keys = ['person_id','index_date'] if 'index_date' in cov_df.columns else ['person_id']
        seq_df     = seq_df.merge(   cov_df, on=merge_keys, how='left')
        comp_df    = comp_df.merge(  cov_df, on=merge_keys, how='left')
        if not reverse_df.empty:
            reverse_df = reverse_df.merge(cov_df, on=merge_keys, how='left')
        if not a_no_b_df.empty:
            a_no_b_df = a_no_b_df.merge(cov_df, on=merge_keys, how='left')
        if not baseline_df.empty:
            baseline_df = baseline_df.merge(cov_df, on=merge_keys, how='left')

        # c) Add demographics (categoricals needed in PS models)
        demogs = persons_df[['person_id', 'sex_at_birth', 'race_ethnicity']]
        seq_df      = seq_df.merge(demogs, on='person_id', how='left')
        comp_df     = comp_df.merge(demogs, on='person_id', how='left')
        if not reverse_df.empty:
            reverse_df = reverse_df.merge(demogs, on='person_id', how='left')
        if not a_no_b_df.empty:
            a_no_b_df = a_no_b_df.merge(demogs, on='person_id', how='left')
        if not baseline_df.empty:
            baseline_df = baseline_df.merge(demogs, on='person_id', how='left')

        # d) PRIMARY PS: A→B vs B‑no‑prior‑A
        seq_p  = seq_df.copy();  seq_p['group']  = 1
        comp_p = comp_df.copy(); comp_p['group'] = 0
        combined_primary = pd.concat([seq_p, comp_p], ignore_index=True)
        covariate_cols = [c for c in CONFIG.COVARIATES_FOR_ADJUSTMENT if c in combined_primary.columns]
        Xp = pd.get_dummies(combined_primary[covariate_cols], dummy_na=True, drop_first=True)
        yp = combined_primary['group']
        model_p = LogisticRegression(solver='liblinear', random_state=CONFIG.RNG_SEED, max_iter=1000)
        model_p.fit(Xp, yp)
        ps_p = model_p.predict_proba(Xp)[:, 1]
        w_p  = np.where(yp == 1, 1 / ps_p, 1 / (1 - ps_p))
        w_p  = np.clip(w_p, None, np.nanpercentile(w_p, 99))
        combined_primary['iptw_weight_primary'] = w_p

        smd_primary_results = {
            'before': calculate_smd(combined_primary, 'group'),
            'after':  calculate_smd(combined_primary, 'group', weight_col='iptw_weight_primary')
        }

        # Map primary weights back
        key_cols = ['person_id', 'index_date']
        prim_w = combined_primary[key_cols + ['iptw_weight_primary']]
        seq_df  = seq_df.merge(prim_w,  on=key_cols, how='left')
        comp_df = comp_df.merge(prim_w,  on=key_cols, how='left')

        # e) REVERSE PS (A→B vs B→A) if reverse cohort is viable
        smd_reverse_results = {}
        if not reverse_df.empty and len(reverse_df) >= CONFIG.MIN_SEQUENCE_SUPPORT:
            seq_r  = seq_df.copy();    seq_r['group']  = 1
            rev_r  = reverse_df.copy(); rev_r['group'] = 0
            combined_reverse = pd.concat([seq_r, rev_r], ignore_index=True)

            cov_cols_r = [c for c in CONFIG.COVARIATES_FOR_ADJUSTMENT if c in combined_reverse.columns]
            Xr = pd.get_dummies(combined_reverse[cov_cols_r], dummy_na=True, drop_first=True)
            yr = combined_reverse['group']
            model_r = LogisticRegression(solver='liblinear', random_state=CONFIG.RNG_SEED, max_iter=1000)
            model_r.fit(Xr, yr)
            ps_r = model_r.predict_proba(Xr)[:, 1]
            w_r  = np.where(yr == 1, 1 / ps_r, 1 / (1 - ps_r))
            w_r  = np.clip(w_r, None, np.nanpercentile(w_r, 99))
            combined_reverse['iptw_weight_reverse'] = w_r

            smd_reverse_results = {
                'before': calculate_smd(combined_reverse, 'group'),
                'after':  calculate_smd(combined_reverse, 'group', weight_col='iptw_weight_reverse')
            }

            rev_w = combined_reverse[key_cols + ['iptw_weight_reverse']]
            seq_df      = seq_df.merge(     rev_w, on=key_cols, how='left')
            reverse_df  = reverse_df.merge( rev_w, on=key_cols, how='left')
        else:
            reverse_df['iptw_weight_reverse'] = np.nan

        # Tertiary PS: A->B vs A_no_B
        smd_tertiary_results = {}
        if not a_no_b_df.empty and len(a_no_b_df) >= CONFIG.MIN_SEQUENCE_SUPPORT:
            seq_t = seq_df.copy();    seq_t['group'] = 1
            ter_t = a_no_b_df.copy(); ter_t['group'] = 0
            comb_t = pd.concat([seq_t, ter_t], ignore_index=True)
            cov_cols_t = [c for c in CONFIG.COVARIATES_FOR_ADJUSTMENT if c in comb_t.columns]
            Xt = pd.get_dummies(comb_t[cov_cols_t], dummy_na=True, drop_first=True)
            yt = comb_t['group']
            model_t = LogisticRegression(solver='liblinear', random_state=CONFIG.RNG_SEED, max_iter=1000)
            model_t.fit(Xt, yt)
            ps_t = model_t.predict_proba(Xt)[:, 1]
            w_t = np.where(yt == 1, 1/ps_t, 1/(1-ps_t))
            w_t = np.clip(w_t, None, np.nanpercentile(w_t, 99))
            comb_t['iptw_weight_tertiary'] = w_t
            keep_t = comb_t[key_cols + ['iptw_weight_tertiary']]
            seq_df    = seq_df.merge(keep_t, on=key_cols, how='left')
            a_no_b_df = a_no_b_df.merge(keep_t, on=key_cols, how='left')
            smd_tertiary_results = {
                'before': calculate_smd(comb_t, 'group'),
                'after':  calculate_smd(comb_t, 'group', weight_col='iptw_weight_tertiary')
            }
        else:
            a_no_b_df['iptw_weight_tertiary'] = np.nan

        # Baseline PS: A->B vs Baseline
        smd_baseline_results = {}
        if not baseline_df.empty and len(baseline_df) >= CONFIG.MIN_SEQUENCE_SUPPORT:
            seq_q  = seq_df.copy();       seq_q['group'] = 1
            base_q = baseline_df.copy();  base_q['group'] = 0
            comb_q = pd.concat([seq_q, base_q], ignore_index=True)
            cov_cols_q = [c for c in CONFIG.COVARIATES_FOR_ADJUSTMENT if c in comb_q.columns]
            Xq = pd.get_dummies(comb_q[cov_cols_q], dummy_na=True, drop_first=True)
            yq = comb_q['group']
            model_q = LogisticRegression(solver='liblinear', random_state=CONFIG.RNG_SEED, max_iter=1000)
            model_q.fit(Xq, yq)
            ps_q = model_q.predict_proba(Xq)[:, 1]
            w_q  = np.where(yq == 1, 1/ps_q, 1/(1-ps_q))
            w_q  = np.clip(w_q, None, np.nanpercentile(w_q, 99))
            comb_q['iptw_weight_baseline'] = w_q
            keep_q = comb_q[key_cols + ['iptw_weight_baseline']]
            seq_df      = seq_df.merge(keep_q, on=key_cols, how='left')
            baseline_df = baseline_df.merge(keep_q, on=key_cols, how='left')
            smd_baseline_results = {
                'before': calculate_smd(comb_q, 'group'),
                'after':  calculate_smd(comb_q, 'group', weight_col='iptw_weight_baseline')
            }
        else:
            baseline_df['iptw_weight_baseline'] = np.nan

        # Ensure weight columns exist for concat
        for _df in (seq_df, comp_df, reverse_df, a_no_b_df, baseline_df):
            if 'iptw_weight_primary' not in _df.columns:
                _df['iptw_weight_primary'] = np.nan
            if 'iptw_weight_reverse' not in _df.columns:
                _df['iptw_weight_reverse'] = np.nan
            if 'iptw_weight_tertiary' not in _df.columns:
                _df['iptw_weight_tertiary'] = np.nan
            if 'iptw_weight_baseline' not in _df.columns:
                _df['iptw_weight_baseline'] = np.nan

        # f) Assemble and save outcome‑specific episodes parquet (unchanged)
        cols_core = [
            'person_id','index_date','time_zero','censor_date','cohort_label',
            'age_at_index','calendar_year_of_index',
            'smoking_status_at_index','bmi_at_index',
            'deprivation_index','utilization_rate','cci_loo',
            'sex_at_birth','race_ethnicity',
            'iptw_weight_primary','iptw_weight_reverse',
            'iptw_weight_tertiary','iptw_weight_baseline'
        ]
        cols_exist = [c for c in cols_core if c in seq_df.columns]  # base schema from seq_df
        ep_out = pd.concat([
            seq_df[cols_exist],
            comp_df[[c for c in cols_exist if c in comp_df.columns]],
            reverse_df[[c for c in cols_exist if c in reverse_df.columns]],
            a_no_b_df[[c for c in cols_exist if c in a_no_b_df.columns]],
            baseline_df[[c for c in cols_exist if c in baseline_df.columns]]
        ], ignore_index=True)

        out_path = episodes_output_dir / f"seq_{seq_idx}__oc_{outcome_name}.parquet"
        smd_metadata = {
            'outcome': outcome_name,
            'smd_primary_comparison': smd_primary_results,
            'smd_reverse_comparison': smd_reverse_results,
            'smd_tertiary_comparison': smd_tertiary_results,
            'smd_baseline_comparison': smd_baseline_results
        }

        try:
            ep_out.to_parquet(
                out_path,
                custom_metadata={'smd_results': json.dumps(smd_metadata, default=_json_default)}
            )
        except TypeError:
            # Older pyarrow: save sidecar JSON
            ep_out.to_parquet(out_path, index=False)
            with open(episodes_output_dir / f"smd_{seq_idx}__oc_{outcome_name}.json", 'w') as f:
                json.dump(smd_metadata, f, indent=2, default=_json_default)

        return True

    except Exception as e:
        log(f"    - ERROR on (seq={seq_idx}, outcome={outcome_name}): {e}")
        return False

    finally:
        processed_pairs.add(pair_key)
        with open(checkpoint_path, 'wb') as f:
            pickle.dump(processed_pairs, f)

# ── 7) Drive the scheduler until quotas satisfied or queues exhausted ---------
def _next_outcome_round_robin(rr_state):
    # rr_state is an integer index into OUTCOMES
    n = len(OUTCOMES)
    for _ in range(n):
        rr_state = (rr_state + 1) % n
        oc = OUTCOMES[rr_state]
        # outcome is active if it needs work and has queue
        need_ok = (remaining_needed[oc] is None) or (remaining_needed[oc] > 0)
        if need_ok and len(queues[oc]) > 0:
            return oc, rr_state
    return None, rr_state

def _next_outcome_random(rng):
    active = [oc for oc in OUTCOMES
              if ((remaining_needed[oc] is None) or (remaining_needed[oc] > 0))
              and (len(queues[oc]) > 0)]
    if not active:
        return None
    return rng.choice(active)

rr_idx = -1
rng = random.Random(int(getattr(CONFIG, 'RNG_SEED', 42)) + 4242)

n_completed_this_run = {oc: 0 for oc in OUTCOMES}
t0 = time.perf_counter()
steps = 0

while True:
    # stop condition: every outcome either reached quota or queue is empty
    active_any = False
    for oc in OUTCOMES:
        need_ok = (remaining_needed[oc] is None) or (remaining_needed[oc] > 0)
        if need_ok and len(queues[oc]) > 0:
            active_any = True
            break
    if not active_any:
        break

    if SCHEDULER_MODE == "random":
        oc = _next_outcome_random(rng)
        if oc is None:
            break
    else:  # round_robin
        oc, rr_idx = _next_outcome_round_robin(rr_idx)
        if oc is None:
            break

    # Pull next pending sequence for this outcome
    seq_idx = queues[oc].popleft()
    steps += 1
    wrote = _process_pair(seq_idx, oc)

    if wrote:
        n_completed_this_run[oc] += 1
        if remaining_needed[oc] is not None:
            remaining_needed[oc] = max(0, remaining_needed[oc] - 1)

    # occasional progress print
    if steps % 25 == 0:
        done_snap = ", ".join([f"{k}:{n_completed_this_run[k]}" for k in OUTCOMES])
        print(f"  • Progress: {steps} attempts; completed this run → {done_snap}")

dt = time.perf_counter() - t0
print(f"\n  • Scheduler loop finished in {dt/60:.1f} min, attempts={steps}.")
print("  • Completed this run per outcome:")
for oc in OUTCOMES:
    print(f"    - {oc:>24}: {n_completed_this_run[oc]} completed (files written)")

print("\n✅ Cell 8 complete: Outcome‑interleaved mining with quotas is cached per (sequence, outcome).")


# In[ ]:


# ──────────────────────────── Cell 9 ─────────────────────────
# Competing Risk Analysis (Discovery Set) with Correct, Per‑Episode Censoring
# ----------------------------------------------------------------------------------------
# Inputs expected from prior cells:
#   • CONFIG (with paths and thresholds), labels_df (first outcome dates), tqdm (optional)
#   • Episode files from Cell 8 in ARTIFACTS_DIR / "discovery_episodes"/ seq_*.parquet
#
# Outputs:
#   • One part file per (sequence, outcome): artifacts/discovery_results_parts/res_{seq}_{outcome}.parquet
#   • Final merged table: artifacts/discovery_results_raw.parquet

import os, warnings, json, math, time
from pathlib import Path
import numpy as np
import pandas as pd
import re

# ── 0) Prereqs & Paths -------------------------------------------------------

required_globals = ['CONFIG', 'labels_df']
for _v in required_globals:
    if _v not in globals():
        raise NameError(f"❌ Prerequisite '{_v}' not found. Please run prior cells.")

EPISODES_DIR       = Path(CONFIG.ARTIFACTS_DIR) / "discovery_episodes"
RESULTS_PARTS_DIR  = Path(CONFIG.ARTIFACTS_DIR) / "discovery_results_parts"
RESULTS_PARTS_DIR.mkdir(exist_ok=True, parents=True)
FINAL_OUT_PATH     = Path(CONFIG.ARTIFACTS_DIR) / "discovery_results_raw.parquet"

# Optional artifact (created in revised Cell 2):
OBS_PERIODS_PATH   = Path(CONFIG.ARTIFACTS_DIR) / "observation_periods_df.parquet"

# ── 1) Tunables --------------------------------------------------------------
BOOT_MAX_REPS        = int(getattr(CONFIG, "N_BOOTSTRAP_MAX", 250))
BOOT_START_REPS      = int(getattr(CONFIG, "N_BOOTSTRAP_START", 100))
BOOT_CHECK_EVERY     = int(getattr(CONFIG, "BOOT_CHECK_EVERY", 20))
BOOT_REL_TOL         = float(getattr(CONFIG, "BOOT_EARLYSTOP_REL_TOL", 0.05))   # 5% relative half-width
BOOT_ABS_TOL         = float(getattr(CONFIG, "BOOT_EARLYSTOP_ABS_TOL", 0.002))  # 0.2% absolute on CIF scale
RNG_SEED             = int(getattr(CONFIG, "RNG_SEED", 42))

MIN_CASES            = int(getattr(CONFIG, "MIN_CASES_FOR_CIF", 20))

# CI method: 'poisson_bootstrap' | 'greenwood' | 'auto'
CI_METHOD            = str(getattr(CONFIG, "CI_METHOD", "poisson_bootstrap")).lower()

HORIZONS_Y           = [int(h) for h in CONFIG.RISK_HORIZONS_YEARS]
HORIZONS_D           = np.array([int(h) * 365 for h in HORIZONS_Y], dtype=np.int32)

# Per‑horizon gating thresholds (optional, have safe defaults)
AT_RISK_THRESH_BY_Y = getattr(CONFIG, "MIN_ATRISK_BY_HORIZON_YEARS", None)
ESS_THRESH_BY_Y     = getattr(CONFIG, "MIN_ESS_BY_HORIZON_YEARS", None)
EVENTS_THRESH_BY_Y  = getattr(CONFIG, "MIN_EVENTS_BY_HORIZON_YEARS", None)
MIN_ATRISK_AT_H     = int(getattr(CONFIG, "MIN_ATRISK_AT_H", 150))
MIN_ESS_AT_H        = getattr(CONFIG, "MIN_ESS_AT_H", None)
MIN_EVENTS_AT_H     = int(getattr(CONFIG, "MIN_EVENTS_AT_H", 15))

# -----------------------------------------------------------------------------
# Utilities
# -----------------------------------------------------------------------------
def _canonical_pid_key(s: pd.Series) -> pd.Series:
    out = s.astype("string").str.strip()
    return out.str.replace(r"\.0+\Z", "", regex=True)

def _warn_if_non_integer_like(s: pd.Series, context: str):
    ss = s.astype("string")
    non_int_like = ss.str.match(r"^\s*[+-]?\d+\.\d+(e[+-]?\d+)?\s*$", flags=re.IGNORECASE, na=False)
    cnt = int(non_int_like.sum())
    if cnt > 0:
        warnings.warn(
            f"[{context}] Detected {cnt:,} person_id values that look non-integer-like. "
            f"These may not match integer IDs.",
            RuntimeWarning
        )

def _write_parquet_atomic(df: pd.DataFrame, path: Path):
    """Atomic parquet write via tmp + replace; explicit Arrow schema casts."""
    import pyarrow as pa, pyarrow.parquet as pq
    tmp = path.with_suffix(path.suffix + ".tmp")

    df = df.copy()
    int_cols = [c for c in df.columns if pd.api.types.is_integer_dtype(df[c])]
    float_cols = [c for c in df.columns if pd.api.types.is_float_dtype(df[c])]
    other_cols = [c for c in df.columns if c not in int_cols + float_cols]

    for c in int_cols:
        df[c] = pd.to_numeric(df[c], errors='coerce').astype('Int64')
    for c in float_cols:
        df[c] = pd.to_numeric(df[c], errors='coerce').astype('float64')
    for c in other_cols:
        df[c] = df[c].astype('string')

    schema = pa.schema(
        [pa.field(c, pa.int64(),  True) for c in int_cols]   +
        [pa.field(c, pa.float64(), True) for c in float_cols] +
        [pa.field(c, pa.string(),  True) for c in other_cols]
    )
    tbl = pa.Table.from_pandas(df, schema=schema, preserve_index=False)
    pq.write_table(tbl, tmp)
    os.replace(tmp, path)

# -----------------------------------------------------------------------------
# Outcome/death maps from labels_df (per‑person first occurrence dates)
# -----------------------------------------------------------------------------
_labels = labels_df.copy()
_labels['pid_key'] = _canonical_pid_key(_labels['person_id'])
_warn_if_non_integer_like(_labels['pid_key'], "labels_df")
_labels['first_event_date'] = pd.to_datetime(_labels['first_event_date'], errors='coerce')

_OUTCOME_MAPS = {
    oc: _labels.loc[_labels['outcome'] == oc].set_index('pid_key')['first_event_date'].to_dict()
    for oc in _labels['outcome'].unique()
}
_DEATH_MAP = _OUTCOME_MAPS.get('death', {})

# -----------------------------------------------------------------------------
# Interval‑aware censoring helpers
# -----------------------------------------------------------------------------
def _ensure_censor_dates(ep_group: pd.DataFrame) -> pd.Series:
    """
    Return a pandas Series of censor_date aligned to ep_group.index.
    Priority:
      1) Use ep_group['censor_date'] if present.
      2) Else infer by joining time_zero to observation_periods_df.parquet.
      3) Else raise a clear error (episodes must carry censor_date in the revised pipeline).
    """
    # 1) If a censor_date column exists, validate and use it.
    if 'censor_date' in ep_group.columns:
        cz = pd.to_datetime(ep_group['censor_date'], errors='coerce')
        tz = pd.to_datetime(ep_group['time_zero'], errors='coerce')
        # invalidate censor dates earlier than time_zero
        cz = cz.where(cz >= tz, other=pd.NaT)
        return cz

    # 2) Try to infer from observation_periods_df.parquet
    if OBS_PERIODS_PATH.exists():
        op = pd.read_parquet(OBS_PERIODS_PATH)
        if op.empty:
            raise RuntimeError(
                "observation_periods_df.parquet is empty. Cannot infer censor_date."
            )
        op = op[['person_id', 'obs_start_date', 'obs_end_date']].copy()
        op['obs_start_date'] = pd.to_datetime(op['obs_start_date'], errors='coerce')
        op['obs_end_date']   = pd.to_datetime(op['obs_end_date'],   errors='coerce')

        base = ep_group[['person_id', 'time_zero']].copy()
        base = base.reset_index(drop=True)
        base['__row_id'] = base.index

        m = base.merge(op, on='person_id', how='left')
        mask = (
            (pd.to_datetime(m['time_zero'], errors='coerce') >= m['obs_start_date']) &
            (pd.to_datetime(m['time_zero'], errors='coerce') <= m['obs_end_date'])
        )
        m = m.loc[mask, ['__row_id', 'obs_end_date']]

        if m.empty:
            # No observation interval covers time_zero for these rows
            return pd.Series(pd.NaT, index=ep_group.index, dtype='datetime64[ns]')

        m = m.groupby('__row_id', as_index=False)['obs_end_date'].max()
        out = pd.Series(pd.NaT, index=base.index, dtype='datetime64[ns]')
        out.loc[m['__row_id'].to_numpy()] = pd.to_datetime(m['obs_end_date'].to_numpy())
        # Re-align to original ep_group index
        out = pd.Series(out.values, index=ep_group.index)
        tz = pd.to_datetime(ep_group['time_zero'], errors='coerce')
        out = out.where(out >= tz, other=pd.NaT)
        return out

    # 3) Cannot proceed without censor_date
    raise RuntimeError(
        "Episode cohort is missing 'censor_date' and observation_periods_df.parquet "
        "was not found. Ensure Cell 7 computes per‑episode censor_date, and Cell 2 "
        "writes observation_periods_df.parquet."
    )

# -----------------------------------------------------------------------------
# Time-to-event arrays (AJ inputs), with interval‑aware censoring
# -----------------------------------------------------------------------------
def _build_tte_arrays_minimal(ep_group: pd.DataFrame, outcome_name: str):
    """
    Returns arrays for AJ:
      duration_days: floor(min(time to event-of-interest, competing, censor))
      event_code:    0=censor, 1=event-of-interest, 2=competing
      weight:        IPTW (>=0)
    Uses per‑episode censor_date (or infers it) for correct censoring.
    """
    if ep_group.empty:
        return dict(duration_days=np.array([], dtype=np.int32),
                    event_code=np.array([], dtype=np.int8),
                    weight=np.array([], dtype=np.float64))

    # Required columns: person_id, time_zero, iptw_weight, (censor_date or inferable)
    required = ['person_id', 'time_zero', 'iptw_weight']
    missing = [c for c in required if c not in ep_group.columns]
    if missing:
        raise KeyError(f"Missing required columns in episode group: {missing}")

    pid_key = _canonical_pid_key(ep_group['person_id'])
    tz      = pd.to_datetime(ep_group['time_zero'], errors='coerce')
    w       = pd.to_numeric(ep_group['iptw_weight'], errors='coerce').astype(float).to_numpy()

    # Outcome & death dates (per person)
    out_dates   = pd.to_datetime(pid_key.map(_OUTCOME_MAPS.get(outcome_name, {})), errors='coerce')
    death_dates = pd.to_datetime(pid_key.map(_DEATH_MAP), errors='coerce')

    # Episode‑specific censor dates
    cz_dates = _ensure_censor_dates(ep_group)

    def _days(a, b):
        return ((a - b).dt.total_seconds() / 86400.0).to_numpy(dtype='float64')

    t_out   = _days(out_dates, tz)
    t_death = _days(death_dates, tz)
    t_cen   = _days(cz_dates, tz)

    # Disallow nonpositive times (must occur strictly after time_zero)
    t_out[~np.isnan(t_out)     & (t_out   <= 0.0)] = np.nan
    t_death[~np.isnan(t_death) & (t_death <= 0.0)] = np.nan
    t_cen[~np.isnan(t_cen)     & (t_cen   <  0.0)] = np.nan

    if outcome_name == 'death':
        # No competing event; only censoring vs event-of-interest (death)
        c0 = np.where(np.isnan(t_cen),   np.inf, t_cen)    # censor
        c1 = np.where(np.isnan(t_death), np.inf, t_death)  # death
        mat   = np.vstack([c0, c1])        # shape (2, N)
        which = np.argmin(mat, axis=0)     # 0=censor, 1=death
        tmin  = np.min(mat, axis=0)
        evt_code = which.astype(np.int8)
    else:
        # Non-death outcome; death is competing event.
        # Build arrays of times from time_zero. np.inf = not observed.
        c0 = np.where(np.isnan(t_cen),   np.inf, t_cen)     # censor
        c1 = np.where(np.isnan(t_out),   np.inf, t_out)     # event-of-interest
        c2 = np.where(np.isnan(t_death), np.inf, t_death)   # competing (death)

        # ── Tie policy: prefer death on exact ties with event-of-interest ──
        # If c1 and c2 tie (finite and equal), bias death slightly earlier.
        # (epsilon in days; 1e-12 ~ 0.086 microseconds)
        eps = 1e-12
        with np.errstate(invalid='ignore'):
            tie_mask = np.isfinite(c1) & np.isfinite(c2) & (np.abs(c1 - c2) <= eps)
        c2_adj = np.where(tie_mask, c2 - eps, c2)

        mat   = np.vstack([c0, c1, c2_adj])   # 0=censor, 1=interest, 2=death (tie-broken)
        which = np.argmin(mat, axis=0)
        tmin  = np.min(mat, axis=0)
        evt_code = which.astype(np.int8)

    valid = np.isfinite(tmin) & np.isfinite(w) & (w > 0)
    dur_days = np.floor(tmin[valid]).astype(np.int32)
    return dict(duration_days=dur_days, event_code=evt_code[valid], weight=w[valid].astype(np.float64))

# -----------------------------------------------------------------------------
# Exact AJ CIF on integer days (fast)
# -----------------------------------------------------------------------------
def aj_cif_int_days(dur_days, evt_code, w, horizons_days, return_risk=False):
    if dur_days.size == 0:
        out = {int(h): float('nan') for h in horizons_days}
        return (out, {int(h): 0.0 for h in horizons_days}) if return_risk else out

    Hmax = int(np.max(horizons_days))
    ok = (dur_days >= 0) & np.isfinite(w) & (w > 0)
    d = dur_days[ok].astype(np.int64)
    e = evt_code[ok].astype(np.int8)
    ww = w[ok].astype(np.float64)

    L = Hmax + 1  # days 0..Hmax

    mask_le = d <= Hmax
    d1 = np.bincount(d[mask_le & (e == 1)], weights=ww[mask_le & (e == 1)], minlength=L)
    d2 = np.bincount(d[mask_le & (e == 2)], weights=ww[mask_le & (e == 2)], minlength=L)
    d0 = np.bincount(d[mask_le & (e == 0)], weights=ww[mask_le & (e == 0)], minlength=L)

    tail = ww[d > Hmax].sum()

    dall = d1 + d2 + d0
    dall_ext = np.empty(L + 1, dtype=np.float64)
    dall_ext[:L] = dall
    dall_ext[L] = tail

    Y_ext = dall_ext[::-1].cumsum()[::-1]   # length L+1
    Y = Y_ext[:-1]                           # 0..Hmax

    haz_all = np.divide(d1 + d2, Y, out=np.zeros_like(Y), where=Y > 0)
    haz_1   = np.divide(d1,       Y, out=np.zeros_like(Y), where=Y > 0)

    S = 1.0
    F = 0.0
    CIF = np.zeros(L, dtype=np.float64)
    for t in range(L):
        F += S * haz_1[t]
        CIF[t] = F
        S *= (1.0 - haz_all[t])

    cif_at  = {int(h): float(CIF[int(h)]) for h in horizons_days}
    if return_risk:
        risk_at = {int(h): float(Y[int(h)]) for h in horizons_days}
        return cif_at, risk_at
    return cif_at

# -----------------------------------------------------------------------------
# Greenwood-type variance for CIF on integer days (fast approximation)
# -----------------------------------------------------------------------------
def greenwood_cif_var_int_days(dur_days, evt_code, w, horizons_days, eps=1e-12):
    if dur_days.size == 0:
        return {int(h): float('nan') for h in horizons_days}

    Hmax = int(np.max(horizons_days))
    L = Hmax + 1

    ok = (dur_days >= 0) & np.isfinite(w) & (w > 0)
    if not np.any(ok):
        return {int(h): 0.0 for h in horizons_days}

    d = dur_days[ok].astype(np.int64, copy=False)
    e = evt_code[ok].astype(np.int8,  copy=False)
    ww = w[ok].astype(np.float64,     copy=False)

    mask_le = d <= Hmax
    d1 = np.bincount(d[mask_le & (e == 1)], weights=ww[mask_le & (e == 1)], minlength=L).astype(np.float64, copy=False)
    d2 = np.bincount(d[mask_le & (e == 2)], weights=ww[mask_le & (e == 2)], minlength=L).astype(np.float64, copy=False)
    d0 = np.bincount(d[mask_le & (e == 0)], weights=ww[mask_le & (e == 0)], minlength=L).astype(np.float64, copy=False)

    tail_w  = float(ww[d > Hmax].sum())

    d_all_w2 = np.bincount(d[mask_le], weights=(ww[mask_le] * ww[mask_le]), minlength=L).astype(np.float64, copy=False)
    tail_w2  = float(np.square(ww[d > Hmax]).sum())

    d_all = d1 + d2 + d0
    d_all_ext = np.empty(L + 1, dtype=np.float64)
    d_all_ext[:L] = d_all
    d_all_ext[L]  = tail_w
    Y = d_all_ext[::-1].cumsum()[::-1][:-1]

    d_all_w2_ext = np.empty(L + 1, dtype=np.float64)
    d_all_w2_ext[:L] = d_all_w2
    d_all_w2_ext[L]  = tail_w2
    Y_w2 = d_all_w2_ext[::-1].cumsum()[::-1][:-1]

    h1 = np.divide(d1,       Y, out=np.zeros_like(Y), where=(Y > 0))
    hA = np.divide(d1 + d2,  Y, out=np.zeros_like(Y), where=(Y > 0))

    F = 0.0
    S = 1.0
    varF = 0.0
    varS = 0.0
    covFS = 0.0
    varF_path = np.zeros(L, dtype=np.float64)

    for t in range(L):
        if Y[t] <= 0.0 or Y_w2[t] <= 0.0:
            varF_path[t] = varF
            continue

        ESS_t = (Y[t] * Y[t]) / max(Y_w2[t], eps)
        if ESS_t > 0.0:
            var_h1 = h1[t] * (1.0 - h1[t]) / ESS_t
            var_hA = hA[t] * (1.0 - hA[t]) / ESS_t
            cov_h1_hA = h1[t] * (1.0 - hA[t]) / ESS_t
        else:
            var_h1 = 0.0
            var_hA = 0.0
            cov_h1_hA = 0.0

        one_minus_hA = (1.0 - hA[t])

        varF_new = (varF + (S * S) * var_h1 + (h1[t] * h1[t]) * varS + 2.0 * h1[t] * covFS)
        varS_new = (one_minus_hA * one_minus_hA) * varS + (S * S) * var_hA
        covFS_new = (one_minus_hA * covFS + one_minus_hA * h1[t] * varS - (S * S) * cov_h1_hA)

        F = F + S * h1[t]
        S = S * one_minus_hA

        varF = max(varF_new, 0.0)
        varS = max(varS_new, 0.0)
        covFS = covFS_new
        varF_path[t] = varF

    return {int(h): float(varF_path[int(h)]) for h in horizons_days}

# -----------------------------------------------------------------------------
# Confidence intervals & gating
# -----------------------------------------------------------------------------
def greenwood_cis_for_groups(A, B, horizons_days, z=1.96):
    cif_A, risk_A = aj_cif_int_days(A['duration_days'], A['event_code'], A['weight'], horizons_days, return_risk=True)
    cif_B, risk_B = aj_cif_int_days(B['duration_days'], B['event_code'], B['weight'], horizons_days, return_risk=True)
    var_A = greenwood_cif_var_int_days(A['duration_days'], A['event_code'], A['weight'], horizons_days)
    var_B = greenwood_cif_var_int_days(B['duration_days'], B['event_code'], B['weight'], horizons_days)

    out = {'method_used': 'greenwood', 'bootstrap_reps_used': 0}
    for h in horizons_days:
        cs, cc = cif_A[h], cif_B[h]
        vs, vc = var_A[h], var_B[h]

        se_s = math.sqrt(vs) if vs == vs and vs >= 0 else float('nan')
        se_c = math.sqrt(vc) if vc == vc and vc >= 0 else float('nan')

        out[f'cif_seq_{h}d']   = cs
        out[f'cif_comp_{h}d']  = cc
        out[f'cif_seq_{h}d_ci_lower']  = max(0.0, cs - z*se_s) if se_s==se_s else float('nan')
        out[f'cif_seq_{h}d_ci_upper']  = min(1.0, cs + z*se_s) if se_s==se_s else float('nan')
        out[f'cif_comp_{h}d_ci_lower'] = max(0.0, cc - z*se_c) if se_c==se_c else float('nan')
        out[f'cif_comp_{h}d_ci_upper'] = min(1.0, cc + z*se_c) if se_c==se_c else float('nan')

        vd = (vs if vs==vs else 0.0) + (vc if vc==vc else 0.0)
        se_d = math.sqrt(vd) if vd >= 0 else float('nan')
        delta = cs - cc
        out[f'delta_cif_{h}d'] = delta
        out[f'delta_cif_{h}d_ci_lower'] = delta - z*se_d if se_d==se_d else float('nan')
        out[f'delta_cif_{h}d_ci_upper'] = delta + z*se_d if se_d==se_d else float('nan')

        if cc > 0:
            rr = cs / cc
            var_log_rr = 0.0
            if cs > 0 and vs==vs:
                var_log_rr += vs/(cs*cs)
            if vc==vc:
                var_log_rr += vc/(cc*cc)
            se_log_rr = math.sqrt(max(var_log_rr, 0.0))
            lo = math.exp(math.log(rr) - z*se_log_rr) if se_log_rr==se_log_rr else float('nan')
            hi = math.exp(math.log(rr) + z*se_log_rr) if se_log_rr==se_log_rr else float('nan')
        else:
            rr, lo, hi = float('nan'), float('nan'), float('nan')

        out[f'risk_ratio_{h}d'] = rr
        out[f'risk_ratio_{h}d_ci_lower'] = lo
        out[f'risk_ratio_{h}d_ci_upper'] = hi

        out[f'at_risk_seq_{h}d']  = risk_A[h]
        out[f'at_risk_comp_{h}d'] = risk_B[h]

    return out

def poisson_bootstrap_cis_for_groups(A, B, horizons_days,
                                     max_reps=BOOT_MAX_REPS, start_reps=BOOT_START_REPS,
                                     check_every=BOOT_CHECK_EVERY,
                                     rel_tol=BOOT_REL_TOL, abs_tol=BOOT_ABS_TOL,
                                     rng=None):
    rng = rng or np.random.default_rng(RNG_SEED)
    cif_A, risk_A = aj_cif_int_days(A['duration_days'], A['event_code'], A['weight'], horizons_days, return_risk=True)
    cif_B, risk_B = aj_cif_int_days(B['duration_days'], B['event_code'], B['weight'], horizons_days, return_risk=True)

    Hs = [int(h) for h in horizons_days]
    hist = {f'A_{h}': [] for h in Hs}
    hist.update({f'B_{h}': [] for h in Hs})
    hist.update({f'D_{h}': [] for h in Hs})

    reps = 0
    def do_rep():
        nonlocal reps
        reps += 1
        mA = rng.poisson(1.0, size=A['weight'].size).astype(np.float64)
        mB = rng.poisson(1.0, size=B['weight'].size).astype(np.float64)
        if mA.sum() == 0 and mB.sum() == 0:
            return
        cifA = aj_cif_int_days(A['duration_days'], A['event_code'], A['weight'] * mA, horizons_days)
        cifB = aj_cif_int_days(B['duration_days'], B['event_code'], B['weight'] * mB, horizons_days)
        for h in Hs:
            a = cifA[h]; b = cifB[h]; d = a - b
            hist[f'A_{h}'].append(a)
            hist[f'B_{h}'].append(b)
            hist[f'D_{h}'].append(d)

    while reps < start_reps:
        do_rep()

    def stable():
        for h in Hs:
            vals = np.array(hist[f'D_{h}'], dtype=float)
            if vals.size < 30:   # ensure enough reps
                return False
            q = np.nanpercentile(vals, [2.5, 97.5])
            hw = 0.5 * (q[1] - q[0])
            denom = max(abs(cif_A[h] - cif_B[h]), 1e-8)
            if not (hw <= abs_tol or (hw/denom) <= rel_tol):
                return False
        return True

    while reps < max_reps and not stable():
        for _ in range(check_every):
            if reps >= max_reps: break
            do_rep()

    out = {'method_used': 'poisson_bootstrap', 'bootstrap_reps_used': reps}
    for h in Hs:
        a0 = float(cif_A[h]); b0 = float(cif_B[h]); d0 = a0 - b0
        valsA = np.array(hist[f'A_{h}'], dtype=float)
        valsB = np.array(hist[f'B_{h}'], dtype=float)
        valsD = np.array(hist[f'D_{h}'], dtype=float)

        loA, hiA = np.nanpercentile(valsA, [2.5, 97.5]) if valsA.size else (np.nan, np.nan)
        loB, hiB = np.nanpercentile(valsB, [2.5, 97.5]) if valsB.size else (np.nan, np.nan)
        loD, hiD = np.nanpercentile(valsD, [2.5, 97.5]) if valsD.size else (np.nan, np.nan)

        # Two-sided bootstrap p-value for ΔCIF
        if valsD.size:
            p_left  = float(np.mean(valsD <= 0.0))
            p_right = float(np.mean(valsD >= 0.0))
            p_val = 2.0 * min(p_left, p_right)
        else:
            p_val = np.nan

        out.update({
            f'cif_seq_{h}d': a0, f'cif_seq_{h}d_ci_lower': float(loA), f'cif_seq_{h}d_ci_upper': float(hiA),
            f'cif_comp_{h}d': b0, f'cif_comp_{h}d_ci_lower': float(loB), f'cif_comp_{h}d_ci_upper': float(hiB),
            f'delta_cif_{h}d': d0, f'delta_cif_{h}d_ci_lower': float(loD), f'delta_cif_{h}d_ci_upper': float(hiD),
            f'p_value_{h}d': float(min(max(p_val, 0.0), 1.0)),
        })
        out[f'risk_ratio_{h}d'] = (a0/b0) if (b0 and b0>0) else float('nan')
        out[f'at_risk_seq_{h}d']  = risk_A[h]
        out[f'at_risk_comp_{h}d'] = risk_B[h]
    return out

def _thr_int(map_by_y, default, year_int):
    """
    Return an int threshold for the given horizon-year.
    - If a per-horizon dict is provided, read from it with a scalar fallback.
    - If the dict is None, use the scalar default (or None if no default).
    """
    if isinstance(map_by_y, dict):
        return int(map_by_y.get(int(year_int), default)) if default is not None else (
            int(map_by_y.get(int(year_int))) if map_by_y.get(int(year_int)) is not None else None
        )
    return int(default) if default is not None else None

def _thr_float(map_by_y, default, year_int):
    """
    Same as _thr_int, but returns float.
    """
    if isinstance(map_by_y, dict):
        return float(map_by_y.get(int(year_int), default)) if default is not None else (
            float(map_by_y.get(int(year_int))) if map_by_y.get(int(year_int)) is not None else None
        )
    return float(default) if default is not None else None


def summarize_risk_ess_events(dur_days, evt_code, w, horizons_days, eps=1e-12):
    """
    Compute per‑horizon summary metrics for a single group:
      • risk_at[h]   = Y(h): weighted number at risk just before day h
      • ess_at[h]    = effective sample size ESS(h) = (sum_w)^2 / sum_w^2
      • events1_at[h]= cumulative weighted count of event‑of‑interest by day h
    """
    res_risk = {int(h): 0.0 for h in horizons_days}
    res_ess  = {int(h): 0.0 for h in horizons_days}
    res_evt1 = {int(h): 0.0 for h in horizons_days}

    if dur_days is None or len(horizons_days) == 0 or np.size(dur_days) == 0:
        return res_risk, res_ess, res_evt1

    Hmax = int(np.max(horizons_days))
    L = Hmax + 1

    d = np.asarray(dur_days)
    e = np.asarray(evt_code)
    ww = np.asarray(w)

    ok = (d >= 0) & np.isfinite(ww) & (ww > 0)
    if not np.any(ok):
        return res_risk, res_ess, res_evt1

    d = d[ok].astype(np.int64, copy=False)
    e = e[ok].astype(np.int8,  copy=False)
    ww = ww[ok].astype(np.float64, copy=False)

    mask_le = d <= Hmax

    d1 = np.bincount(d[mask_le & (e == 1)], weights=ww[mask_le & (e == 1)], minlength=L).astype(np.float64, copy=False)
    d2 = np.bincount(d[mask_le & (e == 2)], weights=ww[mask_le & (e == 2)], minlength=L).astype(np.float64, copy=False)
    d0 = np.bincount(d[mask_le & (e == 0)], weights=ww[mask_le & (e == 0)], minlength=L).astype(np.float64, copy=False)

    d_all = d1 + d2 + d0

    d_all_w2 = np.bincount(d[mask_le], weights=(ww[mask_le] * ww[mask_le]), minlength=L).astype(np.float64, copy=False)

    tail_w  = float(ww[~mask_le].sum())
    tail_w2 = float((ww[~mask_le] * ww[~mask_le]).sum())

    d_all_ext = np.empty(L + 1, dtype=np.float64)
    d_all_ext[:L] = d_all
    d_all_ext[L]  = tail_w
    Y = d_all_ext[::-1].cumsum()[::-1][:-1]

    d_all_w2_ext = np.empty(L + 1, dtype=np.float64)
    d_all_w2_ext[:L] = d_all_w2
    d_all_w2_ext[L]  = tail_w2
    Y_w2 = d_all_w2_ext[::-1].cumsum()[::-1][:-1]

    E1_cum = np.cumsum(d1)

    for h in horizons_days:
        hh = int(h)
        if 0 <= hh < L:
            y   = float(Y[hh])
            y2  = float(Y_w2[hh])
            e1  = float(E1_cum[hh])
            ess = (y * y / max(y2, eps)) if y2 > 0.0 else 0.0
        else:
            y, ess, e1 = 0.0, 0.0, 0.0
        res_risk[hh] = y
        res_ess[hh]  = ess
        res_evt1[hh] = e1

    return res_risk, res_ess, res_evt1

# -----------------------------------------------------------------------------
# Runner for a single (sequence, outcome) with reverse comparator (B→A)
# -----------------------------------------------------------------------------
def analyze_sequence_outcome(ep: pd.DataFrame, outcome_name: str):
    """
    Analyzes A→B vs. B‑no‑prior‑A (primary), and reports B→A (reverse) CIF for context.
    Expects per‑episode censoring via a 'censor_date' column (or will infer it).
    Returns: (rows:list[dict], counts_summary:dict)
    """
    # Build cohort‑specific views with proper weight column and censor_date
    seq = ep.loc[ep['cohort_label'] == 'A_to_B', ['person_id','time_zero','censor_date','iptw_weight_primary']].copy()
    cmp = ep.loc[ep['cohort_label'] == 'B_no_prior_A', ['person_id','time_zero','censor_date','iptw_weight_primary']].copy()
    rev = ep.loc[ep['cohort_label'] == 'B_to_A', ['person_id','time_zero','censor_date','iptw_weight_reverse']].copy()

    seq = seq.rename(columns={'iptw_weight_primary':'iptw_weight'})
    cmp = cmp.rename(columns={'iptw_weight_primary':'iptw_weight'})
    rev = rev.rename(columns={'iptw_weight_reverse':'iptw_weight'})

    A = _build_tte_arrays_minimal(seq, outcome_name)
    B = _build_tte_arrays_minimal(cmp, outcome_name)
    C = _build_tte_arrays_minimal(rev, outcome_name)  # Reverse cohort TTE arrays

    n_seq, n_cases_seq, n_competing_seq = int(A['event_code'].size), int((A['event_code'] == 1).sum()), int((A['event_code'] == 2).sum())
    n_comp, n_cases_comp, n_competing_comp = int(B['event_code'].size), int((B['event_code'] == 1).sum()), int((B['event_code'] == 2).sum())
    n_rev, n_cases_rev, n_competing_rev = int(C['event_code'].size), int((C['event_code'] == 1).sum()), int((C['event_code'] == 2).sum())

    summary_base = {
        'n_seq': n_seq, 'n_cases_seq': n_cases_seq, 'n_competing_seq': n_competing_seq,
        'n_comp': n_comp, 'n_cases_comp': n_cases_comp, 'n_competing_comp': n_competing_comp,
        'n_reverse': n_rev, 'n_cases_reverse': n_cases_rev, 'n_competing_reverse': n_competing_rev
    }

    if (n_cases_seq < MIN_CASES) or (n_cases_comp < MIN_CASES):
        return [], dict(status='skipped_min_cases', **summary_base)

    riskA, essA, evtA = summarize_risk_ess_events(A['duration_days'], A['event_code'], A['weight'], HORIZONS_D)
    riskB, essB, evtB = summarize_risk_ess_events(B['duration_days'], B['event_code'], B['weight'], HORIZONS_D)
    riskC, essC, evtC = summarize_risk_ess_events(C['duration_days'], C['event_code'], C['weight'], HORIZONS_D)

    pass_days, pass_map = [], {}
    for H, Hd in zip(HORIZONS_Y, HORIZONS_D):
        thr_y   = _thr_int(AT_RISK_THRESH_BY_Y, MIN_ATRISK_AT_H, H)
        thr_ess = _thr_float(ESS_THRESH_BY_Y, MIN_ESS_AT_H, H)
        thr_evt = _thr_int(EVENTS_THRESH_BY_Y, MIN_EVENTS_AT_H, H)
        ok = ((riskA.get(Hd, 0) >= thr_y) and (riskB.get(Hd, 0) >= thr_y)) if thr_y is not None else True
        if thr_ess: ok = ok and (essA.get(Hd, 0) >= thr_ess) and (essB.get(Hd, 0) >= thr_ess)
        if thr_evt: ok = ok and (evtA.get(Hd, 0) >= thr_evt) and (evtB.get(Hd, 0) >= thr_evt)
        pass_map[Hd] = ok
        if ok: pass_days.append(Hd)

    cif_rev = aj_cif_int_days(C['duration_days'], C['event_code'], C['weight'], HORIZONS_D) if n_rev > 0 else {h: np.nan for h in HORIZONS_D}

    # Build pair-specific views for tertiary (A_no_B) and baseline comparisons
    try:
        seq_for_ter = ep.loc[ep['cohort_label'] == 'A_to_B', ['person_id','time_zero','censor_date','iptw_weight_tertiary']].copy()
        seq_for_ter = seq_for_ter.rename(columns={'iptw_weight_tertiary':'iptw_weight'})
        ter_view = ep.loc[ep['cohort_label'] == 'A_no_B', ['person_id','time_zero','censor_date','iptw_weight_tertiary']].copy()
        ter_view = ter_view.rename(columns={'iptw_weight_tertiary':'iptw_weight'})
    except KeyError:
        # If columns are absent, create empty frames
        seq_for_ter = pd.DataFrame(columns=['person_id','time_zero','censor_date','iptw_weight'])
        ter_view = pd.DataFrame(columns=['person_id','time_zero','censor_date','iptw_weight'])

    try:
        seq_for_base = ep.loc[ep['cohort_label'] == 'A_to_B', ['person_id','time_zero','censor_date','iptw_weight_baseline']].copy()
        seq_for_base = seq_for_base.rename(columns={'iptw_weight_baseline':'iptw_weight'})
        base_view = ep.loc[ep['cohort_label'] == 'Baseline', ['person_id','time_zero','censor_date','iptw_weight_baseline']].copy()
        base_view = base_view.rename(columns={'iptw_weight_baseline':'iptw_weight'})
    except KeyError:
        seq_for_base = pd.DataFrame(columns=['person_id','time_zero','censor_date','iptw_weight'])
        base_view = pd.DataFrame(columns=['person_id','time_zero','censor_date','iptw_weight'])

    T = _build_tte_arrays_minimal(ter_view, outcome_name)
    S_T = _build_tte_arrays_minimal(seq_for_ter, outcome_name)
    Q = _build_tte_arrays_minimal(base_view, outcome_name)
    S_Q = _build_tte_arrays_minimal(seq_for_base, outcome_name)

    # Compute stats for tertiary and baseline if viable
    if (T['event_code'].size >= MIN_CASES) and (S_T['event_code'].size >= MIN_CASES):
        if CI_METHOD == 'poisson_bootstrap':
            res_ter = poisson_bootstrap_cis_for_groups(S_T, T, HORIZONS_D, rng=np.random.default_rng(RNG_SEED+7))
        else:
            res_ter = greenwood_cis_for_groups(S_T, T, HORIZONS_D)
        riskT, essT, evtT = summarize_risk_ess_events(T['duration_days'], T['event_code'], T['weight'], HORIZONS_D)
    else:
        res_ter = {}
        riskT = {h: 0.0 for h in HORIZONS_D}
        essT = {h: 0.0 for h in HORIZONS_D}
        evtT = {h: 0.0 for h in HORIZONS_D}

    if (Q['event_code'].size >= MIN_CASES) and (S_Q['event_code'].size >= MIN_CASES):
        if CI_METHOD == 'poisson_bootstrap':
            res_base = poisson_bootstrap_cis_for_groups(S_Q, Q, HORIZONS_D, rng=np.random.default_rng(RNG_SEED+13))
        else:
            res_base = greenwood_cis_for_groups(S_Q, Q, HORIZONS_D)
        riskQ, essQ, evtQ = summarize_risk_ess_events(Q['duration_days'], Q['event_code'], Q['weight'], HORIZONS_D)
    else:
        res_base = {}
        riskQ = {h: 0.0 for h in HORIZONS_D}
        essQ = {h: 0.0 for h in HORIZONS_D}
        evtQ = {h: 0.0 for h in HORIZONS_D}

    # Add group counts to summary
    n_ter = int(T['event_code'].size)
    n_cases_ter = int((T['event_code'] == 1).sum()) if n_ter > 0 else 0
    n_competing_ter = int((T['event_code'] == 2).sum()) if n_ter > 0 else 0
    n_base = int(Q['event_code'].size)
    n_cases_base = int((Q['event_code'] == 1).sum()) if n_base > 0 else 0
    n_competing_base = int((Q['event_code'] == 2).sum()) if n_base > 0 else 0
    summary_base.update({
        'n_tertiary': n_ter, 'n_cases_tertiary': n_cases_ter, 'n_competing_tertiary': n_competing_ter,
        'n_baseline': n_base, 'n_cases_baseline': n_cases_base, 'n_competing_baseline': n_competing_base
    })

    if len(pass_days) == 0:
        rows = []
        for H, Hd in zip(HORIZONS_Y, HORIZONS_D):
            row = {'horizon_years': H, 'outcome': outcome_name, 'status': 'gated_out', 'method_used': 'gated', 'bootstrap_reps_used': 0, **summary_base}
            row.update({k: np.nan for k in ['cif_seq','cif_comp','delta_cif','risk_ratio','cif_reverse']})
            row.update({f'at_risk_seq': riskA.get(Hd,0), f'at_risk_comp': riskB.get(Hd,0), f'at_risk_reverse': riskC.get(Hd,0)})
            rows.append(row)
        return rows, dict(status='gated_out_all', **summary_base)

    rng = np.random.default_rng(RNG_SEED + hash(outcome_name) % (2**31 - 1))
    if CI_METHOD == 'poisson_bootstrap':
        res = poisson_bootstrap_cis_for_groups(A, B, pass_days, rng=rng)
    else:  # 'auto' or 'greenwood'
        res = greenwood_cis_for_groups(A, B, pass_days)
        if CI_METHOD == 'auto':
            # Escalate to bootstrap if 95% CI for ΔCIF crosses 0 at any passed horizon
            escalate = any(not (res.get(f'delta_cif_{Hd}d_ci_lower', 1) > 0 or res.get(f'delta_cif_{Hd}d_ci_upper', -1) < 0) for Hd in pass_days)
            if escalate:
                res = poisson_bootstrap_cis_for_groups(A, B, pass_days, rng=rng)

    rows = []
    for H, Hd in zip(HORIZONS_Y, HORIZONS_D):
        base_row = {'horizon_years': H, 'outcome': outcome_name, **summary_base}
        if pass_map[Hd]:
            row = {'status': 'success', 'method_used': res.get('method_used',''), 'bootstrap_reps_used': res.get('bootstrap_reps_used',0)}
            for k in ['cif_seq','cif_comp','delta_cif','risk_ratio']:
                for suffix in ['', '_ci_lower', '_ci_upper']:
                    row[k+suffix] = res.get(f'{k}_{Hd}d{suffix}', np.nan)
            # NEW: p-value for ΔCIF at this horizon
            row['p_value'] = res.get(f'p_value_{Hd}d', np.nan)
        else:
            row = {'status': 'gated_out', 'method_used': 'gated', 'bootstrap_reps_used': 0}
            row.update({k: np.nan for k in ['cif_seq','cif_comp','delta_cif','risk_ratio','p_value']})

        row.update({
            'cif_reverse': cif_rev.get(Hd, np.nan),
            'at_risk_seq': riskA.get(Hd,0), 'at_risk_comp': riskB.get(Hd,0), 'at_risk_reverse': riskC.get(Hd,0),
            'ess_seq': essA.get(Hd,0), 'ess_comp': essB.get(Hd,0), 'ess_reverse': essC.get(Hd,0),
            'events_seq': evtA.get(Hd,0), 'events_comp': evtB.get(Hd,0), 'events_reverse': evtC.get(Hd,0)
        })
        # Add tertiary (A_no_B) and baseline comparator metrics
        row.update({
            'cif_tertiary': res_ter.get(f'cif_comp_{Hd}d', np.nan),
            'cif_tertiary_ci_lower': res_ter.get(f'cif_comp_{Hd}d_ci_lower', np.nan),
            'cif_tertiary_ci_upper': res_ter.get(f'cif_comp_{Hd}d_ci_upper', np.nan),
            'risk_ratio_tertiary': res_ter.get(f'risk_ratio_{Hd}d', np.nan),
            'risk_ratio_tertiary_ci_lower': res_ter.get(f'risk_ratio_{Hd}d_ci_lower', np.nan),
            'risk_ratio_tertiary_ci_upper': res_ter.get(f'risk_ratio_{Hd}d_ci_upper', np.nan),
            'delta_cif_tertiary': res_ter.get(f'delta_cif_{Hd}d', np.nan),
            'delta_cif_tertiary_ci_lower': res_ter.get(f'delta_cif_{Hd}d_ci_lower', np.nan),
            'delta_cif_tertiary_ci_upper': res_ter.get(f'delta_cif_{Hd}d_ci_upper', np.nan),
            'p_value_tertiary': res_ter.get(f'p_value_{Hd}d', np.nan),
            'at_risk_tertiary': riskT.get(Hd, 0.0),
            'ess_tertiary': essT.get(Hd, 0.0),
            'events_tertiary': evtT.get(Hd, 0.0),
            'cif_baseline': res_base.get(f'cif_comp_{Hd}d', np.nan),
            'cif_baseline_ci_lower': res_base.get(f'cif_comp_{Hd}d_ci_lower', np.nan),
            'cif_baseline_ci_upper': res_base.get(f'cif_comp_{Hd}d_ci_upper', np.nan),
            'risk_ratio_baseline': res_base.get(f'risk_ratio_{Hd}d', np.nan),
            'risk_ratio_baseline_ci_lower': res_base.get(f'risk_ratio_{Hd}d_ci_lower', np.nan),
            'risk_ratio_baseline_ci_upper': res_base.get(f'risk_ratio_{Hd}d_ci_upper', np.nan),
            'delta_cif_baseline': res_base.get(f'delta_cif_{Hd}d', np.nan),
            'delta_cif_baseline_ci_lower': res_base.get(f'delta_cif_{Hd}d_ci_lower', np.nan),
            'delta_cif_baseline_ci_upper': res_base.get(f'delta_cif_{Hd}d_ci_upper', np.nan),
            'p_value_baseline': res_base.get(f'p_value_{Hd}d', np.nan),
            'at_risk_baseline': riskQ.get(Hd, 0.0),
            'ess_baseline': essQ.get(Hd, 0.0),
            'events_baseline': evtQ.get(Hd, 0.0)
        })
        rows.append({**base_row, **row})

    return rows, dict(status='success', **summary_base)

# -----------------------------------------------------------------------------
# Driver over (sequence, outcome) with idempotent resume
# -----------------------------------------------------------------------------
print("▶ Cell 9: Starting analysis over episode files...")
if not EPISODES_DIR.exists():
    raise FileNotFoundError(f"Episode directory not found: {EPISODES_DIR}")

OUTCOMES = list(CONFIG.OUTCOME_DEFINITIONS.keys())

def part_path_for(seq_idx, outcome):
    return RESULTS_PARTS_DIR / f"res_{seq_idx}_{outcome}.parquet"

episode_files = sorted(EPISODES_DIR.glob("seq_*__oc_*.parquet"),
                       key=lambda p: int(p.stem.split('_')[1]))

tasks = []
for p in episode_files:
    stem = p.stem  # e.g., "seq_12__oc_dementia"
    seq_idx = int(stem.split('_')[1])
    oc_from_file = stem.split('__oc_')[1]
    if not part_path_for(seq_idx, oc_from_file).exists():
        tasks.append((seq_idx, p, oc_from_file))

print(f"  • Found {len(episode_files):,} episode files; {len(tasks):,} tasks remaining.")

t0_all = time.perf_counter()

_iterator = tasks
if 'tqdm' in globals() and (tqdm is not None):  # tqdm imported in Cell 0 if available
    _iterator = tqdm(tasks, desc="Analyzing Sequence-Outcomes")

for seq_idx, ep_path, outcome in _iterator:
    t0 = time.perf_counter()
    try:
        ep = pd.read_parquet(ep_path)

        # Ensure time_zero is datetime and (if present) censor_date is datetime
        ep['time_zero'] = pd.to_datetime(ep['time_zero'], errors='coerce')
        if 'censor_date' in ep.columns:
            ep['censor_date'] = pd.to_datetime(ep['censor_date'], errors='coerce')

        rows, summary = analyze_sequence_outcome(ep, outcome)

        if rows:
            df_part = pd.DataFrame(rows)
            df_part['sequence_idx'] = int(seq_idx)
        else:
            df_part = pd.DataFrame([{'sequence_idx': int(seq_idx), 'outcome': str(outcome), 'status': str(summary.get('status','error')), **summary}])

        _write_parquet_atomic(df_part, part_path_for(seq_idx, outcome))
        dt = time.perf_counter() - t0
        if 'tqdm' in globals() and (tqdm is not None):
            _iterator.set_postfix_str(f"seq={seq_idx}, out={outcome}, status={summary['status']}, t={dt:.2f}s")

    except KeyboardInterrupt:
        print("🛑 KeyboardInterrupt: stopping safely.")
        break
    except Exception as e:
        err_df = pd.DataFrame([{'sequence_idx': seq_idx, 'outcome': outcome, 'status': 'error', 'error_msg': str(e)}])
        _write_parquet_atomic(err_df, part_path_for(seq_idx, outcome))

print(f"  • Loop complete in {(time.perf_counter()-t0_all)/60:.1f} min.")

# -----------------------------------------------------------------------------
# Consolidate all parts
# -----------------------------------------------------------------------------
print("\n  • Consolidating all result parts into a single file...")
parts = list(RESULTS_PARTS_DIR.glob("res_*.parquet"))
if parts:
    final_df = pd.concat([pd.read_parquet(p) for p in parts], ignore_index=True)
    _write_parquet_atomic(final_df, FINAL_OUT_PATH)
    print(f"    - Saved final results to: {FINAL_OUT_PATH.name} ({len(final_df):,} rows).")
else:
    print("    - No result parts were found; nothing to consolidate.")

print("✅ Cell 9 complete.")


# In[ ]:


# ──────────────────────────── Cell 10 ─────────────────────────────
# Candidate Selection and FDR Control (with Secondary Comparator)
# ---------------------------------------------------------------------------
# This cell takes the raw statistical results from the discovery set analysis
# (Cell 9), applies pre-specified selection criteria to identify promising
# signals, and then rigorously controls the false discovery rate (FDR) using
# the Benjamini-Hochberg procedure.
#
# The selection process is now enhanced to require that a candidate sequence
# (A→B) not only shows a significant risk increase compared to the primary
# comparator (B with no prior A), but also demonstrates directional
# specificity by having a higher risk than the reverse order (B→A).
#
# The final output is a "frozen" list of candidate signals that have passed
# these stricter criteria, ready for validation in the confirmation set.

import pandas as pd
import numpy as np
from scipy.stats import norm
from statsmodels.stats.multitest import multipletests
from pathlib import Path

print("▶ Cell 10: Candidate Selection and FDR Control with Secondary Comparator...")

# --- 1. Prerequisite Checks and Path Definitions ---
required_globals = ['CONFIG', 'log']
for var in required_globals:
    if var not in globals():
        raise NameError(f"❌ Prerequisite '{var}' not found. Please run preceding cells first.")

input_path = CONFIG.ARTIFACTS_DIR / "discovery_results_raw.parquet"
output_path = CONFIG.ARTIFACTS_DIR / "confirmed_candidates_frozen.csv"

if not input_path.exists():
    raise FileNotFoundError(
        f"The discovery results file was not found at: {input_path}. "
        "Please successfully run Cell 9 before this cell."
    )

# --- 2. Load and Prepare Discovery Results ---
print(f"  • Loading raw discovery results from: {input_path.name}")
discovery_df = pd.read_parquet(input_path)
print(f"    - Loaded {len(discovery_df):,} raw result rows.")

# --- 3. Calculate Metrics for Primary and Secondary Comparisons ---
print("  • Calculating p-values and risk ratios for all comparisons...")

if 'p_value' not in discovery_df.columns:
    discovery_df['p_value'] = np.nan

need_p = discovery_df['p_value'].isna()

if need_p.any() and {'delta_cif_ci_lower','delta_cif_ci_upper','delta_cif'}.issubset(discovery_df.columns):
    print("    - Filling missing p-values from 95% CIs on ΔCIF.")
    delta_cif_ci_width = discovery_df['delta_cif_ci_upper'] - discovery_df['delta_cif_ci_lower']
    delta_cif_se = delta_cif_ci_width / (2 * 1.96)
    z_scores = discovery_df['delta_cif'].divide(delta_cif_se).replace([np.inf, -np.inf], np.nan)
    discovery_df.loc[need_p, 'p_value'] = 2 * (1 - norm.cdf(np.abs(z_scores.loc[need_p])))
else:
    print("    - Using existing p-values (bootstrap) where available; none to backfill.")


# Secondary Comparison (A→B vs. B→A, the reverse order)
# The upstream cell provides the point estimate for cif_reverse.
# We calculate the risk ratio to check for directional specificity.
with np.errstate(divide='ignore', invalid='ignore'):
    discovery_df['risk_ratio_reverse'] = discovery_df['cif_seq'] / discovery_df['cif_reverse']
discovery_df['risk_ratio_reverse'].replace([np.inf, -np.inf], np.nan, inplace=True)

# --- 4. Apply Pre-selection Criteria with Directionality Check ---
# These filters identify signals that are statistically suggestive, have a
# meaningful effect size, AND show evidence that the order A→B matters.
MIN_RISK_RATIO_PRIMARY = 1.2
MIN_RISK_RATIO_REVERSE = 1.1  # A→B must be at least 10% higher risk than B→A
MIN_RISK_DIFFERENCE = 0.005 # 0.5% absolute risk increase vs. primary comparator

print(f"  • Applying pre-selection criteria:")
print(f"    - RR (vs. B-no-prior-A) > {MIN_RISK_RATIO_PRIMARY}")
print(f"    - ΔCIF (vs. B-no-prior-A) > {MIN_RISK_DIFFERENCE*100:.1f}%")
print(f"    - p-value (vs. B-no-prior-A) < 0.05")
print(f"    - RR (vs. reverse B→A) > {MIN_RISK_RATIO_REVERSE}")

preselected_df = discovery_df[
    (discovery_df['risk_ratio'].fillna(0) > MIN_RISK_RATIO_PRIMARY) &
    (discovery_df['delta_cif'].fillna(0) > MIN_RISK_DIFFERENCE) &
    (discovery_df['p_value'].fillna(1) < 0.05) &
    (discovery_df['risk_ratio_reverse'].fillna(0) > MIN_RISK_RATIO_REVERSE)
].copy()
print(f"    - {len(preselected_df):,} hypotheses passed all pre-selection filters.")

# OPTIONAL extra gates using tertiary/baseline comparisons
REQUIRE_TERTIARY = False
REQUIRE_BASELINE = False

if REQUIRE_TERTIARY and {'risk_ratio_tertiary','delta_cif_tertiary'}.issubset(discovery_df.columns):
    preselected_df = preselected_df[
        (preselected_df['risk_ratio_tertiary'].fillna(0) > 1.10) &
        (preselected_df['delta_cif_tertiary'].fillna(0) > 0.003)
    ]

if REQUIRE_BASELINE and {'risk_ratio_baseline','delta_cif_baseline'}.issubset(discovery_df.columns):
    preselected_df = preselected_df[
        (preselected_df['risk_ratio_baseline'].fillna(0) > 1.05) &
        (preselected_df['delta_cif_baseline'].fillna(0) > 0.002)
    ]

# --- 5. Global Benjamini–Hochberg FDR (alpha=CONFIG.FDR_ALPHA) ---
print(f"  • Applying GLOBAL BH FDR (alpha={CONFIG.FDR_ALPHA}) before any filters...")

# Ensure we have p-values
if 'p_value' not in discovery_df.columns:
    discovery_df['p_value'] = np.nan

mask = discovery_df['p_value'].notna()
discovery_df['q_value'] = np.nan  # overwrite/standardize name used downstream
if mask.any():
    _, q_all, _, _ = multipletests(discovery_df.loc[mask, 'p_value'],
                                   alpha=CONFIG.FDR_ALPHA, method='fdr_bh')
    discovery_df.loc[mask, 'q_value'] = q_all

# Primary BH hits (no other filters yet)
bh_hits_df = discovery_df[discovery_df['q_value'] < CONFIG.FDR_ALPHA].copy()
print(f"    - BH discoveries (global): {len(bh_hits_df):,}")

# --- 6. Optional post‑hoc triage (effect size / directionality) -------------
# These filters are applied AFTER BH. Removing rejections cannot increase FDR.
MIN_RISK_RATIO_PRIMARY = 1.2
MIN_RISK_RATIO_REVERSE = 1.1
MIN_RISK_DIFFERENCE    = 0.005  # 0.5% absolute

posthoc_filtered_df = bh_hits_df[
    (bh_hits_df['risk_ratio'].fillna(0)           > MIN_RISK_RATIO_PRIMARY) &
    (bh_hits_df['delta_cif'].fillna(0)            > MIN_RISK_DIFFERENCE)    &
    (bh_hits_df['risk_ratio_reverse'].fillna(0)   > MIN_RISK_RATIO_REVERSE)
].copy()
print(f"    - After post‑hoc triage: {len(posthoc_filtered_df):,}")

# Freeze the list we pass to confirmation
confirmed_candidates_df = posthoc_filtered_df.copy()

# --- Save the Frozen Candidate List (unchanged filename/shape) ---
print(f"  • Saving the frozen list of confirmed candidates to: {output_path.name}")
final_cols = ['sequence_idx','outcome','horizon_years','risk_ratio',
              'risk_ratio_reverse','delta_cif','p_value','q_value']
final_cols_exist = [c for c in final_cols if c in confirmed_candidates_df.columns]
if not confirmed_candidates_df.empty:
    confirmed_candidates_df.sort_values(by='q_value', inplace=True)
    confirmed_candidates_df[final_cols_exist].to_csv(output_path, index=False)
else:
    pd.DataFrame(columns=final_cols_exist).to_csv(output_path, index=False)
unique_advancing_sequences = confirmed_candidates_df['sequence_idx'].nunique()
print("\n--- Candidate Selection Summary ---")
print(f"Total hypotheses analyzed:      {len(discovery_df):,}")
print(f"BH discoveries (global):        {len(bh_hits_df):,}")
print(f"Final candidates after triage:  {len(confirmed_candidates_df):,}")
print(f"Unique sequences advancing:     {unique_advancing_sequences:,}")
print("-----------------------------------")


# In[ ]:


# ──────────────────────────── Cell 11 (REVISED • outcome‑aware) ────────────────────────────
# Confirmation Analysis with Secondary Comparator (B→A)
# + Leave‑One‑Out CCI, SES (deprivation_index), and Healthcare Utilization
# ------------------------------------------------------------------------------------------

import pandas as pd
import numpy as np
from pathlib import Path
import time
from sklearn.linear_model import LogisticRegression

print("▶ Cell 11: Performing Confirmation Analysis with expanded covariates...")

# --- 1) Prerequisite Checks ---------------------------------------------------
required_globals = [
    'CONFIG', 'log', 'confirmation_pids', 'sequence_map_df', 'persons_df',
    'events_df', 'analyze_sequence_outcome'
]
for var in required_globals:
    if var not in globals():
        raise NameError(f"❌ Prerequisite '{var}' not found. Please run preceding cells first.")

# Observation periods are needed for OP‑aware censoring
if 'observation_periods_df' not in globals():
    _op_path = CONFIG.ARTIFACTS_DIR / "observation_periods_df.parquet"
    if not _op_path.exists():
        raise FileNotFoundError(
            "❌ observation_periods_df is not loaded and the cache file "
            f"'{_op_path}' was not found. Please run Cell 2."
        )
    observation_periods_df = pd.read_parquet(_op_path)
    print(f"  • Loaded observation periods from cache: {_op_path.name} ({len(observation_periods_df):,} rows)")

# Upgraded covariate engine must be available with the new signature
if 'get_covariates_at_index' not in globals():
    raise NameError("❌ Upgraded `get_covariates_at_index` not found. Please update Cell 6 per the plan.")
# (We do not introspect its signature; we will call it with (series, A, B, outcome).)

candidates_path = CONFIG.ARTIFACTS_DIR / "confirmed_candidates_frozen.csv"
output_path     = CONFIG.ARTIFACTS_DIR / "final_results_raw.parquet"

# --- 2) Resumability ----------------------------------------------------------
if output_path.exists():
    final_results_df = pd.read_parquet(output_path)
    print(f"  • Found cached final results. Loading from: {output_path.name}")
else:
    print("  • No final results cache found. Starting confirmation analysis...")
    if not candidates_path.exists():
        raise FileNotFoundError(
            f"The frozen candidate list was not found at: {candidates_path}. "
            "Please successfully run Cell 10 before this cell."
        )

    # --- 3) Load frozen candidates -------------------------------------------
    confirmed_candidates_df = pd.read_csv(candidates_path)
    print(f"  • Loaded {len(confirmed_candidates_df)} candidate hypotheses to validate.")

    if confirmed_candidates_df.empty:
        print("    - Candidate list is empty. Nothing to confirm. Creating an empty results file.")
        final_results_df = pd.DataFrame()
    else:
        # --- 4) Local helpers -------------------------------------------------
        def _finalize_cohort_without_covariates(candidates: pd.DataFrame, label: str) -> pd.DataFrame:
            """
            Gap‑aware, OP‑aware finalizer WITHOUT invoking the covariate engine.
            Ensures BOTH index_date and time_zero lie within the SAME observation interval.
            Sets censor_date = interval's obs_end_date.
            Computes age_at_index and calendar_year_of_index, and attaches demographics
            (sex_at_birth, race_ethnicity) needed later for PS models.
            """
            cols_out = [
                'person_id', 'index_date', 'time_zero', 'censor_date', 'cohort_label',
                'age_at_index', 'calendar_year_of_index', 'sex_at_birth', 'race_ethnicity'
            ]
            if candidates is None or candidates.empty:
                return pd.DataFrame(columns=cols_out)

            df = candidates.copy()
            df['index_date'] = pd.to_datetime(df['index_date'], errors='coerce')
            df = df.dropna(subset=['person_id', 'index_date'])
            if df.empty:
                return pd.DataFrame(columns=cols_out)

            # Apply latency and ensure OP membership for BOTH index_date and time_zero
            latency_delta = pd.Timedelta(days=int(CONFIG.LATENCY_DAYS))
            df['time_zero'] = df['index_date'] + latency_delta

            op_sub = observation_periods_df[
                observation_periods_df['person_id'].isin(df['person_id'].unique())
            ].copy()
            if not pd.api.types.is_datetime64_any_dtype(op_sub['obs_start_date']):
                op_sub['obs_start_date'] = pd.to_datetime(op_sub['obs_start_date'], errors='coerce')
            if not pd.api.types.is_datetime64_any_dtype(op_sub['obs_end_date']):
                op_sub['obs_end_date'] = pd.to_datetime(op_sub['obs_end_date'], errors='coerce')

            merged = df.merge(op_sub, on='person_id', how='inner')
            within_index = (merged['index_date'] >= merged['obs_start_date']) & (merged['index_date'] <= merged['obs_end_date'])
            within_tz    = (merged['time_zero']  >= merged['obs_start_date']) & (merged['time_zero']  <= merged['obs_end_date'])
            valid = merged[within_index & within_tz].copy()
            if valid.empty:
                return pd.DataFrame(columns=cols_out)

            LOOKBACK_LABELS = {'B_no_prior_A', 'A_no_B', 'Baseline'}
            if label in LOOKBACK_LABELS:
                lookback_delta = pd.Timedelta(days=int(getattr(CONFIG, 'RISKSET_MIN_LOOKBACK_DAYS', 1825)))
                
                # The observation period must have started at least 'lookback_delta' days before the index date (date of B).
                had_sufficient_lookback = (valid['obs_start_date'] <= (valid['index_date'] - lookback_delta))
                valid = valid.loc[had_sufficient_lookback].copy()

                # If no episodes remain after this filter, return an empty frame.
                if valid.empty:
                    return pd.DataFrame(columns=cols_out)

            # Episode‑level censoring at the interval end; deduplicate overlapping intervals
            valid['censor_date'] = valid['obs_end_date']
            valid.sort_values(['person_id', 'index_date', 'censor_date'], inplace=True)
            valid = valid.drop_duplicates(subset=['person_id', 'index_date'], keep='first')

            # Attach demographics + static covariates needed by PS
            demo_cols = ['person_id', 'year_of_birth', 'sex_at_birth', 'race_ethnicity']
            valid = valid.merge(persons_df[demo_cols], on='person_id', how='left')
            valid['age_at_index'] = (valid['index_date'].dt.year - valid['year_of_birth']).astype('Int64')
            valid['calendar_year_of_index'] = valid['index_date'].dt.year.astype('Int64')
            valid['cohort_label'] = label

            return valid[cols_out].reset_index(drop=True)

        def _build_episodes_for_sequence(event_A_vid: int, event_B_vid: int, person_ids: set) -> dict:
            """
            Build A→B, B‑no‑prior‑A, and B→A candidates from first‑occurrence dates,
            then finalize with OP‑aware rules (no covariates attached here).
            """
            ev = events_df[
                (events_df['person_id'].isin(person_ids)) &
                (events_df['feature_concept_id'].isin([event_A_vid, event_B_vid]))
            ].copy()

            # First observed dates for A and B
            first_A = (
                ev[ev['feature_concept_id'] == event_A_vid]
                .groupby('person_id', sort=False)['event_date'].min().rename('date_A')
            )
            first_B = (
                ev[ev['feature_concept_id'] == event_B_vid]
                .groupby('person_id', sort=False)['event_date'].min().rename('date_B')
            )

            merged_dates = pd.merge(
                first_A.to_frame(), first_B.to_frame(),
                left_index=True, right_index=True, how='outer'
            ).reset_index().rename(columns={'index': 'person_id'})

            policy = str(getattr(CONFIG, 'SAME_DAY_POLICY', 'exclude')).lower()
            max_gap = pd.Timedelta(days=int(getattr(CONFIG, 'EPISODE_MAX_GAP_DAYS', 1825)))

            if policy == 'allow':
                a_to_b_mask = (
                    merged_dates['date_A'].notna() & merged_dates['date_B'].notna() &
                    (merged_dates['date_B'] >= merged_dates['date_A']) &
                    ((merged_dates['date_B'] - merged_dates['date_A']) <= max_gap)
                )
                b_no_a_mask = (
                    merged_dates['date_B'].notna() &
                    (merged_dates['date_A'].isna() | (merged_dates['date_A'] >= merged_dates['date_B']))
                )
                b_to_a_mask = (
                    merged_dates['date_A'].notna() & merged_dates['date_B'].notna() &
                    (merged_dates['date_A'] >= merged_dates['date_B']) &
                    ((merged_dates['date_A'] - merged_dates['date_B']) <= max_gap)
                )
            elif policy == 'require':
                a_to_b_mask = (merged_dates['date_A'].notna()) & (merged_dates['date_B'] == merged_dates['date_A'])
                b_no_a_mask = (merged_dates['date_B'].notna()) & (merged_dates['date_A'].isna())
                b_to_a_mask = (merged_dates['date_A'].notna()) & (merged_dates['date_B'] == merged_dates['date_A'])
            else:  # 'exclude'
                a_to_b_mask = (
                    merged_dates['date_A'].notna() & merged_dates['date_B'].notna() &
                    (merged_dates['date_B'] > merged_dates['date_A']) &
                    ((merged_dates['date_B'] - merged_dates['date_A']) <= max_gap)
                )
                b_no_a_mask = (
                    merged_dates['date_B'].notna() &
                    (merged_dates['date_A'].isna() |
                     (merged_dates['date_A'].notna() & (merged_dates['date_A'] > merged_dates['date_B'])))
                )
                b_to_a_mask = (
                    merged_dates['date_A'].notna() & merged_dates['date_B'].notna() &
                    (merged_dates['date_A'] > merged_dates['date_B']) &
                    ((merged_dates['date_A'] - merged_dates['date_B']) <= max_gap)
                )

            a_to_b_cand = (
                merged_dates.loc[a_to_b_mask, ['person_id', 'date_B']]
                .rename(columns={'date_B': 'index_date'})
                .dropna(subset=['index_date'])
                .reset_index(drop=True)
            )
            b_no_a_cand = (
                merged_dates.loc[b_no_a_mask, ['person_id', 'date_B']]
                .rename(columns={'date_B': 'index_date'})
                .dropna(subset=['index_date'])
                .reset_index(drop=True)
            )
            b_to_a_cand = (
                merged_dates.loc[b_to_a_mask, ['person_id', 'date_A']]
                .rename(columns={'date_A': 'index_date'})
                .dropna(subset=['index_date'])
                .reset_index(drop=True)
            )
            # Define "subsequent B" by SAME_DAY_POLICY, then derive A_no_B
            if policy == 'allow':      # B >= A counts as subsequent
                _subseq = (merged_dates['date_B'] >= merged_dates['date_A'])
            elif policy == 'require':  # only same day counts as subsequent
                _subseq = (merged_dates['date_B'] == merged_dates['date_A'])
            else:                      # 'exclude' -> strictly after
                _subseq = (merged_dates['date_B'] > merged_dates['date_A'])

            b_after_a = (
                merged_dates['date_A'].notna() & merged_dates['date_B'].notna() & _subseq
            )
            within_gap = (merged_dates['date_B'] - merged_dates['date_A']) <= max_gap
            a_no_b_mask = merged_dates['date_A'].notna() & (~(b_after_a & within_gap).fillna(False))
            a_no_b_cand = (
                merged_dates.loc[a_no_b_mask, ['person_id', 'date_A']]
                .rename(columns={'date_A': 'index_date'})
                .dropna(subset=['index_date'])
                .reset_index(drop=True)
            )

            # Baseline: one episode per person with adequate lookback and time_zero inside interval
            def _baseline_candidates(pid_set: set) -> pd.DataFrame:
                op_sub = observation_periods_df[observation_periods_df['person_id'].isin(pid_set)].copy()
                lb  = pd.Timedelta(days=int(getattr(CONFIG, 'RISKSET_MIN_LOOKBACK_DAYS', 1825)))
                lat = pd.Timedelta(days=int(getattr(CONFIG, 'LATENCY_DAYS', 90)))
                op_sub['candidate_index'] = op_sub['obs_start_date'] + lb
                ok = (op_sub['candidate_index'] + lat <= op_sub['obs_end_date'])
                base = (
                    op_sub.loc[ok, ['person_id', 'candidate_index']]
                    .sort_values(['person_id', 'candidate_index'])
                    .drop_duplicates('person_id')
                    .rename(columns={'candidate_index': 'index_date'})
                    .reset_index(drop=True)
                )
                return base

            baseline_cand = _baseline_candidates(person_ids)

            return {
                'A_to_B':       _finalize_cohort_without_covariates(a_to_b_cand, 'A_to_B'),
                'B_no_prior_A': _finalize_cohort_without_covariates(b_no_a_cand, 'B_no_prior_A'),
                'B_to_A':       _finalize_cohort_without_covariates(b_to_a_cand, 'B_to_A'),
                'A_no_B':       _finalize_cohort_without_covariates(a_no_b_cand, 'A_no_B'),
                'Baseline':     _finalize_cohort_without_covariates(baseline_cand, 'Baseline'),
            }

        def _ensure_covariate_columns(df: pd.DataFrame) -> pd.DataFrame:
            """
            Ensure all covariate columns referenced in CONFIG.COVARIATES_FOR_ADJUSTMENT exist.
            Missing numeric covariates are filled with 0; missing categoricals with 'Unknown'.
            """
            if df is None or df.empty:
                return df

            # Heuristics for types
            categorical_covs = {'sex_at_birth', 'race_ethnicity', 'smoking_status_at_index', 'bmi_at_index'}
            numeric_covs     = set(CONFIG.COVARIATES_FOR_ADJUSTMENT) - categorical_covs

            for col in CONFIG.COVARIATES_FOR_ADJUSTMENT:
                if col not in df.columns:
                    if col in numeric_covs:
                        df[col] = 0
                    else:
                        df[col] = "Unknown"

            # Compute missing basics if needed
            if 'calendar_year_of_index' in CONFIG.COVARIATES_FOR_ADJUSTMENT and 'calendar_year_of_index' not in df.columns:
                df['calendar_year_of_index'] = pd.to_datetime(df['index_date'], errors='coerce').dt.year.astype('Int64')

            if 'age_at_index' in CONFIG.COVARIATES_FOR_ADJUSTMENT and 'age_at_index' not in df.columns:
                df = df.merge(persons_df[['person_id', 'year_of_birth']], on='person_id', how='left')
                df['age_at_index'] = (pd.to_datetime(df['index_date'], errors='coerce').dt.year - df['year_of_birth']).astype('Int64')

            return df

        def _fit_ps_weights(df_treated: pd.DataFrame, df_control: pd.DataFrame, weight_col_name: str) -> pd.DataFrame:
            """
            Fit a logistic regression propensity model on CONFIG.COVARIATES_FOR_ADJUSTMENT and
            return a DataFrame with columns ['person_id', 'index_date', weight_col_name].
            """
            t = df_treated.copy(); t['group'] = 1
            c = df_control.copy(); c['group'] = 0
            combined = pd.concat([t, c], ignore_index=True)

            # Attach demographics that may be needed
            if not {'sex_at_birth', 'race_ethnicity'}.issubset(combined.columns):
                combined = combined.merge(
                    persons_df[['person_id', 'sex_at_birth', 'race_ethnicity']],
                    on='person_id', how='left'
                )

            # Ensure covariate columns exist and are well‑typed
            combined = _ensure_covariate_columns(combined)

            cov_cols = [col for col in CONFIG.COVARIATES_FOR_ADJUSTMENT if col in combined.columns]
            if not cov_cols:
                raise ValueError("No covariates available for PS model. Check CONFIG.COVARIATES_FOR_ADJUSTMENT.")

            X = pd.get_dummies(combined[cov_cols], dummy_na=True, drop_first=True)
            y = combined['group'].astype(int)

            model = LogisticRegression(solver='liblinear', random_state=CONFIG.RNG_SEED, max_iter=1000)
            model.fit(X, y)
            ps = model.predict_proba(X)[:, 1]

            # IPTW (stabilization not applied; clip at 99th percentile)
            w = np.where(y == 1, 1 / ps, 1 / (1 - ps))
            w = np.clip(w, None, np.nanpercentile(w, 99))

            out = combined[['person_id', 'index_date']].copy()
            out[weight_col_name] = w
            return out

        # --- 5) Outcome‑first confirmation loop --------------------------------
        all_confirmation_results = []

        grouped_by_outcome = confirmed_candidates_df.groupby('outcome', sort=False)
        iterator = grouped_by_outcome
        if 'tqdm' in globals() and tqdm:
            iterator = tqdm(grouped_by_outcome, desc="Confirming Signals (outcome‑first)", unit="outcome")

        for outcome_name, candidates_for_outcome in iterator:
            # Unique sequences to validate for this outcome
            seq_indices = sorted(pd.unique(candidates_for_outcome['sequence_idx']))
            if 'tqdm' in globals() and tqdm:
                _seq_iter = tqdm(seq_indices, leave=False, desc=f"{outcome_name}")
            else:
                _seq_iter = seq_indices

            for seq_idx in _seq_iter:
                try:
                    # a) Resolve the (A, B) concept IDs for this sequence
                    seq_row = sequence_map_df.set_index('sequence_idx').loc[seq_idx]
                    event_A, event_B = int(seq_row['event_A_vid']), int(seq_row['event_B_vid'])

                    # b) Build OP‑aware episodes (without covariates)
                    episode_dict = _build_episodes_for_sequence(event_A, event_B, confirmation_pids)
                    seq_df      = episode_dict.get('A_to_B',          pd.DataFrame()).copy()
                    comp_df     = episode_dict.get('B_no_prior_A',    pd.DataFrame()).copy()
                    reverse_df  = episode_dict.get('B_to_A',          pd.DataFrame()).copy()
                    a_no_b_df   = episode_dict.get('A_no_B',          pd.DataFrame()).copy()
                    baseline_df = episode_dict.get('Baseline',        pd.DataFrame()).copy()

                    # Basic size gate for primary comparison
                    if len(seq_df) < CONFIG.MIN_SEQUENCE_SUPPORT or len(comp_df) < CONFIG.MIN_SEQUENCE_SUPPORT:
                        continue

                    # c) Compute outcome‑aware covariates ONCE and merge to all cohorts
                    all_eps = pd.concat([
                        seq_df[['person_id', 'index_date']],
                        comp_df[['person_id', 'index_date']],
                        reverse_df[['person_id', 'index_date']],
                        a_no_b_df[['person_id', 'index_date']],
                        baseline_df[['person_id', 'index_date']]
                    ], ignore_index=True).drop_duplicates()

                    # Build a Series of index dates keyed by person_id; duplicates per person are OK
                    idx_series = pd.Series(all_eps['index_date'].values, index=all_eps['person_id'].values)
                    cov_df = get_covariates_at_index(idx_series, event_A, event_B, str(outcome_name))
                    # Ensure datetime and merge keys
                    if 'index_date' in cov_df.columns:
                        cov_df['index_date'] = pd.to_datetime(cov_df['index_date'], errors='coerce')

                    merge_keys = ['person_id', 'index_date'] if 'index_date' in cov_df.columns else ['person_id']
                    for name, _df in [('seq', seq_df), ('comp', comp_df), ('rev', reverse_df), ('ter', a_no_b_df), ('base', baseline_df)]:
                        if _df.empty:
                            continue
                        _df['index_date'] = pd.to_datetime(_df['index_date'], errors='coerce')
                        _df = _df.merge(cov_df, on=merge_keys, how='left')
                        if name == 'seq':      seq_df = _df
                        elif name == 'comp':   comp_df = _df
                        elif name == 'rev':    reverse_df = _df
                        elif name == 'ter':    a_no_b_df = _df
                        else:                  baseline_df = _df

                    # d) Fit PS weights (primary): A→B vs B‑no‑prior‑A
                    w_primary = _fit_ps_weights(seq_df, comp_df, 'iptw_weight_primary')
                    seq_df  = seq_df.merge( w_primary, on=['person_id', 'index_date'], how='left')
                    comp_df = comp_df.merge(w_primary, on=['person_id', 'index_date'], how='left')

                    # e) Fit PS weights (reverse): A→B vs B→A (if viable)
                    if not reverse_df.empty and len(reverse_df) >= CONFIG.MIN_SEQUENCE_SUPPORT:
                        w_reverse = _fit_ps_weights(seq_df, reverse_df, 'iptw_weight_reverse')
                        seq_df     = seq_df.merge(    w_reverse, on=['person_id', 'index_date'], how='left')
                        reverse_df = reverse_df.merge(w_reverse, on=['person_id', 'index_date'], how='left')
                    else:
                        reverse_df['iptw_weight_reverse'] = np.nan

                    # e.2) Fit PS weights (tertiary): A->B vs A_no_B (if viable)
                    if not a_no_b_df.empty and len(a_no_b_df) >= CONFIG.MIN_SEQUENCE_SUPPORT:
                        w_ter = _fit_ps_weights(seq_df, a_no_b_df, 'iptw_weight_tertiary')
                        seq_df    = seq_df.merge(   w_ter, on=['person_id', 'index_date'], how='left')
                        a_no_b_df = a_no_b_df.merge(w_ter, on=['person_id', 'index_date'], how='left')
                    else:
                        a_no_b_df['iptw_weight_tertiary'] = np.nan

                    # e.3) Fit PS weights (baseline): A->B vs Baseline (if viable)
                    if not baseline_df.empty and len(baseline_df) >= CONFIG.MIN_SEQUENCE_SUPPORT:
                        w_base = _fit_ps_weights(seq_df, baseline_df, 'iptw_weight_baseline')
                        seq_df      = seq_df.merge(   w_base, on=['person_id', 'index_date'], how='left')
                        baseline_df = baseline_df.merge(w_base, on=['person_id', 'index_date'], how='left')
                    else:
                        baseline_df['iptw_weight_baseline'] = np.nan

                    # f) Assemble a unified episodes DataFrame with all weight columns
                    #    (analyze_sequence_outcome will pick the correct weight per cohort)
                    for df_ in (seq_df, comp_df, reverse_df, a_no_b_df, baseline_df):
                        if df_.empty:
                            continue
                        if 'iptw_weight_primary' not in df_.columns:
                            df_['iptw_weight_primary'] = np.nan
                        if 'iptw_weight_reverse' not in df_.columns:
                            df_['iptw_weight_reverse'] = np.nan
                        if 'iptw_weight_tertiary' not in df_.columns:
                            df_['iptw_weight_tertiary'] = np.nan
                        if 'iptw_weight_baseline' not in df_.columns:
                            df_['iptw_weight_baseline'] = np.nan

                    cols_core = [
                        'person_id','index_date','time_zero','censor_date','cohort_label',
                        'age_at_index','calendar_year_of_index'
                    ]
                    # Keep any covariates columns (optional for QA/debug)
                    cov_keep = [c for c in CONFIG.COVARIATES_FOR_ADJUSTMENT if c in seq_df.columns or c in comp_df.columns or c in reverse_df.columns]
                    weight_cols = ['iptw_weight_primary','iptw_weight_reverse','iptw_weight_tertiary','iptw_weight_baseline']

                    ep = pd.concat([
                        seq_df[cols_core + cov_keep + weight_cols],
                        comp_df[cols_core + cov_keep + weight_cols],
                        reverse_df[cols_core + cov_keep + weight_cols],
                        a_no_b_df[cols_core + cov_keep + weight_cols],
                        baseline_df[cols_core + cov_keep + weight_cols]
                    ], ignore_index=True)

                    # Ensure datetime types for analysis
                    ep['time_zero']   = pd.to_datetime(ep['time_zero'],   errors='coerce')
                    ep['censor_date'] = pd.to_datetime(ep['censor_date'], errors='coerce')

                    # g) Run competing‑risk analysis for this outcome
                    results_list, _summary = analyze_sequence_outcome(ep, str(outcome_name))
                    for result in results_list:
                        result['sequence_idx'] = int(seq_idx)
                        all_confirmation_results.append(result)

                except Exception as e:
                    log(f"    - ERROR processing sequence {seq_idx} for outcome '{outcome_name}': {e}")

        # --- 6) Assemble and save final results --------------------------------
        if not all_confirmation_results:
            log("    - No results were generated during confirmation analysis.")
            final_results_df = pd.DataFrame()
        else:
            final_results_df = pd.DataFrame(all_confirmation_results)
            log(f"\n  • Assembled {len(final_results_df):,} final result rows from confirmation analysis.")

    final_results_df.to_parquet(output_path, index=False)
    log(f"  • Saved final raw results to: {output_path.name}")

# --- 7) Final Summary ---------------------------------------------------------
if not final_results_df.empty:
    print(f"\n--- Confirmation Analysis Summary ---")
    print(f"Total confirmed hypotheses validated: {len(final_results_df):,}")
    display_cols = [
        'sequence_idx', 'outcome', 'horizon_years', 'cif_seq', 'cif_comp', 'cif_reverse',
        'n_seq', 'n_comp', 'n_reverse'
    ]
    existing_display_cols = [c for c in display_cols if c in final_results_df.columns]
    display(final_results_df[existing_display_cols].head(10))
    print("-------------------------------------")

print("✅ Cell 11 complete: Final results saved with expanded covariates.")


# In[ ]:


# ──────────────────────────── Cell 11b ────────────────────────────────
# Benjamini–Hochberg FDR on Confirmation p-values

import pandas as pd
from statsmodels.stats.multitest import multipletests

print("▶ Cell 11b: Applying BH FDR on confirmation results...")
conf_path_in  = CONFIG.ARTIFACTS_DIR / "final_results_raw.parquet"
conf_path_out = CONFIG.ARTIFACTS_DIR / "final_results_with_q.parquet"

if not conf_path_in.exists():
    raise FileNotFoundError(f"Missing confirmation results at {conf_path_in}")

df = pd.read_parquet(conf_path_in)
if df.empty or 'p_value' not in df.columns:
    print("  • No p-values found in confirmation results. Skipping BH.")
    df.to_parquet(conf_path_out, index=False)
else:
    print(f"  • Applying GLOBAL BH FDR on all confirmation tests (alpha={CONFIG.FDR_ALPHA})...")
    mask = df['p_value'].notna()
    df['q_value_confirm'] = pd.NA
    if mask.sum() > 0:
        _, q_all, _, _ = multipletests(df.loc[mask, 'p_value'],
                                       alpha=CONFIG.FDR_ALPHA, method='fdr_bh')
        df.loc[mask, 'q_value_confirm'] = q_all
    df.to_parquet(conf_path_out, index=False)
    print(f"  • Saved: {conf_path_out.name}")


# In[ ]:


# ──────────────────────────── Cell 12 ────────────────────────────────
# Sensitivity Analyses with Secondary Comparator (B→A) — OP‑aware & bug‑fixed
# ---------------------------------------------------------------------------
#
# Outputs:
#   artifacts/sensitivity_results.parquet
#
# Prereqs expected in the runtime:
#   CONFIG, log, confirmation_pids, sequence_map_df, persons_df, events_df,
#   get_covariates_at_index (Cell 6), analyze_sequence_outcome (Cell 9).
#   Also needs observation_periods_df in memory or on disk (Cell 2 artifact).

import pandas as pd
import numpy as np
from pathlib import Path
import time
from sklearn.linear_model import LogisticRegression

print("▶ Cell 12: Running Sensitivity Analyses on Top Confirmed Signals with Secondary Comparator...")

def _coalesce_demog_cols(df: pd.DataFrame) -> pd.DataFrame:
    """
    If merges have produced 'sex_at_birth_x/_y' or 'race_ethnicity_x/_y',
    collapse them into a single canonical column without suffixes.
    """
    for col in ['sex_at_birth', 'race_ethnicity']:
        cx, cy = f'{col}_x', f'{col}_y'
        if col not in df.columns:
            if cx in df.columns and cy in df.columns:
                df[col] = df[cx].combine_first(df[cy])
                df.drop(columns=[cx, cy], inplace=True)
            elif cx in df.columns:
                df.rename(columns={cx: col}, inplace=True)
            elif cy in df.columns:
                df.rename(columns={cy: col}, inplace=True)
    return df

def _ensure_covariate_columns(df: pd.DataFrame) -> pd.DataFrame:
    """
    Ensure every column from CONFIG.COVARIATES_FOR_ADJUSTMENT exists.
    Missing categoricals → 'Unknown'; missing numerics → 0.
    """
    if df is None or df.empty:
        return df
    categorical_covs = {'sex_at_birth', 'race_ethnicity', 'smoking_status_at_index', 'bmi_at_index'}
    for col in CONFIG.COVARIATES_FOR_ADJUSTMENT:
        if col not in df.columns:
            df[col] = "Unknown" if col in categorical_covs else 0
    return df


# --- 1. Prerequisite Checks and Paths ---
required_globals = [
    'CONFIG', 'log', 'confirmation_pids', 'persons_df',
    'events_df', 'get_covariates_at_index', 'analyze_sequence_outcome'
]
for var in required_globals:
    if var not in globals():
        raise NameError(f"❌ Prerequisite '{var}' not found. Please run the preceding cells first.")

# Load observation periods (from memory if available, else from artifact)
if 'observation_periods_df' not in globals():
    _op_path = CONFIG.ARTIFACTS_DIR / "observation_periods_df.parquet"
    if not _op_path.exists():
        raise FileNotFoundError(
            "❌ observation_periods_df is not loaded and the cache file "
            f"'{_op_path}' was not found. Please run Cell 2."
        )
    observation_periods_df = pd.read_parquet(_op_path)
    print(f"  • Loaded observation periods from cache: {_op_path.name} ({len(observation_periods_df):,} rows)")

# Load sequence map if not present (usually set in Cell 8)
if 'sequence_map_df' not in globals():
    _seq_map_path = CONFIG.ARTIFACTS_DIR / "sequence_map_df.parquet"
    if not _seq_map_path.exists():
        raise FileNotFoundError(
            "❌ sequence_map_df is not loaded and the cache file "
            f"'{_seq_map_path}' was not found. Please run Cell 8."
        )
    sequence_map_df = pd.read_parquet(_seq_map_path)
    print(f"  • Loaded sequence map from cache: {_seq_map_path.name} ({len(sequence_map_df):,} rows)")

final_results_path = CONFIG.ARTIFACTS_DIR / "final_results_raw.parquet"
output_path        = CONFIG.ARTIFACTS_DIR / "sensitivity_results.parquet"

if not final_results_path.exists():
    raise FileNotFoundError(f"Final results not found at {final_results_path}. Run Cell 11 first.")

final_results_df = pd.read_parquet(final_results_path)
if final_results_df.empty:
    print("  • Final results file is empty. Skipping sensitivity analysis.")
    pd.DataFrame().to_parquet(output_path)
    raise SystemExit

# --- 2. Define Sensitivity Parameter Grid (pre-specified) ---
SENSITIVITY_GRID = [
    {'latency_days': 30,  'max_gap_days': 1825},
    {'latency_days': 90,  'max_gap_days': 1825},  # Primary analysis
    {'latency_days': 180, 'max_gap_days': 1825},
    {'latency_days': 90,  'max_gap_days': 365},
    {'latency_days': 90,  'max_gap_days': 3650},
]
print(f"  • Defined {len(SENSITIVITY_GRID)} parameter sets for sensitivity analysis.")

# --- 3. Select Top Signals for Sensitivity Analysis ---
# Choose top 5 signals at 2 years by risk ratio (from confirmation results)
top_signals = (
    final_results_df[final_results_df['horizon_years'] == 2]
    .dropna(subset=['risk_ratio'])
    .nlargest(5, 'risk_ratio')
    .merge(sequence_map_df, on='sequence_idx', how='left')
)
print(f"  • Selected top {len(top_signals)} signals for analysis.")

# --- 4. Utilities: SAME_DAY_POLICY helpers and OP-aware finalizer ---

def _same_day_policy_masks(merged_dates: pd.DataFrame, policy: str, max_gap_delta: pd.Timedelta):
    """
    Returns boolean masks (a_to_b, b_no_a, b_to_a) according to SAME_DAY_POLICY.
    merged_dates has columns: ['person_id','date_A','date_B'].
    """
    policy = str(policy).lower()
    if policy == 'allow':
        a_to_b = (
            merged_dates['date_A'].notna() & merged_dates['date_B'].notna() &
            (merged_dates['date_B'] >= merged_dates['date_A']) &
            ((merged_dates['date_B'] - merged_dates['date_A']) <= max_gap_delta)
        )
        b_no_a = (
            merged_dates['date_B'].notna() &
            (merged_dates['date_A'].isna() | (merged_dates['date_A'] >= merged_dates['date_B']))
        )
        b_to_a = (
            merged_dates['date_A'].notna() & merged_dates['date_B'].notna() &
            (merged_dates['date_A'] >= merged_dates['date_B']) &
            ((merged_dates['date_A'] - merged_dates['date_B']) <= max_gap_delta)
        )
    elif policy == 'require':
        a_to_b = (merged_dates['date_A'].notna()) & (merged_dates['date_B'] == merged_dates['date_A'])
        b_no_a = (merged_dates['date_B'].notna()) & (merged_dates['date_A'].isna())
        b_to_a = (merged_dates['date_A'].notna()) & (merged_dates['date_B'] == merged_dates['date_A'])
    else:  # 'exclude' (default)
        a_to_b = (
            merged_dates['date_A'].notna() & merged_dates['date_B'].notna() &
            (merged_dates['date_B'] > merged_dates['date_A']) &
            ((merged_dates['date_B'] - merged_dates['date_A']) <= max_gap_delta)
        )
        b_no_a = (
            merged_dates['date_B'].notna() &
            (merged_dates['date_A'].isna() | (merged_dates['date_A'] > merged_dates['date_B']))
        )
        b_to_a = (
            merged_dates['date_A'].notna() & merged_dates['date_B'].notna() &
            (merged_dates['date_A'] > merged_dates['date_B']) &
            ((merged_dates['date_A'] - merged_dates['date_B']) <= max_gap_delta)
        )
    return a_to_b, b_no_a, b_to_a


def _finalize_cohort_op_aware(candidates: pd.DataFrame,
                              label: str,
                              latency_days: int) -> pd.DataFrame:
    cols_out = [
        'person_id','index_date','time_zero','censor_date','cohort_label',
        'age_at_index','calendar_year_of_index'
    ]
    if candidates is None or candidates.empty:
        return pd.DataFrame(columns=cols_out)

    df = candidates.copy()
    df['index_date'] = pd.to_datetime(df['index_date'], errors='coerce')
    df = df.dropna(subset=['person_id','index_date'])
    if df.empty:
        return pd.DataFrame(columns=cols_out)

    op = observation_periods_df[['person_id','obs_start_date','obs_end_date']].copy()
    op['obs_start_date'] = pd.to_datetime(op['obs_start_date'], errors='coerce')
    op['obs_end_date']   = pd.to_datetime(op['obs_end_date'],   errors='coerce')

    m = df.merge(op, on='person_id', how='inner')
    in_index = (m['index_date'] >= m['obs_start_date']) & (m['index_date'] <= m['obs_end_date'])
    m = m.loc[in_index.values].copy()
    if m.empty:
        return pd.DataFrame(columns=cols_out)

    m['time_zero'] = m['index_date'] + pd.Timedelta(days=int(latency_days))
    in_tz = (m['time_zero'] >= m['obs_start_date']) & (m['time_zero'] <= m['obs_end_date'])
    m = m.loc[in_tz].copy()
    if m.empty:
        return pd.DataFrame(columns=cols_out)

    # Require adequate lookback for selected cohorts (consistent with Cell 7/11)
    LOOKBACK_LABELS = {'B_no_prior_A', 'A_no_B', 'Baseline'}
    if label in LOOKBACK_LABELS:
        lookback_delta = pd.Timedelta(days=int(getattr(CONFIG, 'RISKSET_MIN_LOOKBACK_DAYS', 1825)))
        m = m.loc[m['obs_start_date'] <= (m['index_date'] - lookback_delta)].copy()
        if m.empty:
            return pd.DataFrame(columns=cols_out)

    m['censor_date'] = m['obs_end_date']
    m.sort_values(['person_id','index_date','censor_date'], inplace=True)
    m = m.drop_duplicates(subset=['person_id','index_date'], keep='first')

    m = m.merge(persons_df[['person_id','year_of_birth']], on='person_id', how='left')
    m['age_at_index'] = (m['index_date'].dt.year - m['year_of_birth']).astype('Int64')
    m['calendar_year_of_index'] = m['index_date'].dt.year.astype('Int64')
    m['cohort_label'] = label

    return m[cols_out].reset_index(drop=True)


def build_episodes_flexible(event_A_vid: int,
                             event_B_vid: int,
                             person_ids: set,
                             latency_days: int,
                             max_gap_days: int) -> dict:
    """
    Build A→B, B-no-prior-A, and B→A cohorts for a given (A,B), enforcing SAME_DAY_POLICY
    and OP-aware interval rules. Returns dict with DataFrames for keys:
      'A_to_B', 'B_no_prior_A', 'B_to_A'
    """
    ev = events_df[
        (events_df['person_id'].isin(person_ids)) &
        (events_df['feature_concept_id'].isin([event_A_vid, event_B_vid]))
    ].copy()

    first_A = (
        ev[ev['feature_concept_id'] == event_A_vid]
        .groupby('person_id', sort=False)['event_date'].min().rename('date_A')
    )
    first_B = (
        ev[ev['feature_concept_id'] == event_B_vid]
        .groupby('person_id', sort=False)['event_date'].min().rename('date_B')
    )
    merged = pd.merge(
        first_A.to_frame(), first_B.to_frame(),
        left_index=True, right_index=True, how='outer'
    ).reset_index().rename(columns={'index': 'person_id'})

    policy = str(getattr(CONFIG, 'SAME_DAY_POLICY', 'exclude')).lower()
    max_gap_delta = pd.Timedelta(days=int(max_gap_days))
    a_to_b_mask, b_no_a_mask, b_to_a_mask = _same_day_policy_masks(merged, policy, max_gap_delta)

    a_to_b_cand = (
        merged.loc[a_to_b_mask, ['person_id','date_B']]
        .rename(columns={'date_B':'index_date'})
        .dropna(subset=['index_date'])
        .reset_index(drop=True)
    )
    b_no_a_cand = (
        merged.loc[b_no_a_mask, ['person_id','date_B']]
        .rename(columns={'date_B':'index_date'})
        .dropna(subset=['index_date'])
        .reset_index(drop=True)
    )
    b_to_a_cand = (
        merged.loc[b_to_a_mask, ['person_id','date_A']]
        .rename(columns={'date_A':'index_date'})
        .dropna(subset=['index_date'])
        .reset_index(drop=True)
    )

    # Derive A_no_B by policy and gap
    if policy == 'allow':      # B >= A counts as subsequent
        _subseq = (merged['date_B'] >= merged['date_A'])
    elif policy == 'require':  # only same day counts as subsequent
        _subseq = (merged['date_B'] == merged['date_A'])
    else:                      # 'exclude' -> strictly after
        _subseq = (merged['date_B'] > merged['date_A'])

    b_after_a = (
        merged['date_A'].notna() & merged['date_B'].notna() & _subseq
    )
    within_gap = (merged['date_B'] - merged['date_A']) <= max_gap_delta
    a_no_b_mask = merged['date_A'].notna() & (~(b_after_a & within_gap).fillna(False))
    a_no_b_cand = (
        merged.loc[a_no_b_mask, ['person_id','date_A']]
        .rename(columns={'date_A':'index_date'})
        .dropna(subset=['index_date'])
        .reset_index(drop=True)
    )

    # Baseline: one OP-aware episode per person with adequate lookback and time_zero inside same interval
    def _baseline_candidates(pid_set: set) -> pd.DataFrame:
        op_sub = observation_periods_df[observation_periods_df['person_id'].isin(pid_set)].copy()
        lb  = pd.Timedelta(days=int(getattr(CONFIG, 'RISKSET_MIN_LOOKBACK_DAYS', 1825)))
        lat = pd.Timedelta(days=int(latency_days))
        op_sub['candidate_index'] = op_sub['obs_start_date'] + lb
        ok = (op_sub['candidate_index'] + lat <= op_sub['obs_end_date'])
        base = (
            op_sub.loc[ok, ['person_id','candidate_index']]
            .sort_values(['person_id','candidate_index'])
            .drop_duplicates('person_id')
            .rename(columns={'candidate_index':'index_date'})
            .reset_index(drop=True)
        )
        return base

    baseline_cand = _baseline_candidates(person_ids)

    return {
        'A_to_B':       _finalize_cohort_op_aware(a_to_b_cand, 'A_to_B', latency_days),
        'B_no_prior_A': _finalize_cohort_op_aware(b_no_a_cand, 'B_no_prior_A', latency_days),
        'B_to_A':       _finalize_cohort_op_aware(b_to_a_cand, 'B_to_A', latency_days),
        'A_no_B':       _finalize_cohort_op_aware(a_no_b_cand, 'A_no_B', latency_days),
        'Baseline':     _finalize_cohort_op_aware(baseline_cand, 'Baseline', latency_days),
    }

# --- 5. Main loop over selected signals and parameter sets ---
all_sensitivity_results = []
total_iters = len(top_signals) * len(SENSITIVITY_GRID)

# Optional progress bar if tqdm is available from Cell 0
_progress = None
try:
    if 'tqdm' in globals() and (tqdm is not None):
        _progress = tqdm(total=total_iters, desc="Running Sensitivity Analyses")
except Exception:
    _progress = None

for _, signal_row in top_signals.iterrows():
    seq_idx  = int(signal_row['sequence_idx'])
    outcome  = str(signal_row['outcome'] if 'outcome' in signal_row else signal_row['Outcome'])
    event_A  = int(signal_row['event_A_vid'])
    event_B  = int(signal_row['event_B_vid'])

    for params in SENSITIVITY_GRID:
        try:
            ep_dict    = build_episodes_flexible(event_A, event_B, confirmation_pids,
                                                 params['latency_days'], params['max_gap_days'])
            seq_df      = ep_dict['A_to_B'].copy()
            comp_df     = ep_dict['B_no_prior_A'].copy()
            reverse_df  = ep_dict['B_to_A'].copy()
            a_no_b_df   = ep_dict.get('A_no_B', pd.DataFrame()).copy()
            baseline_df = ep_dict.get('Baseline', pd.DataFrame()).copy()

            # --- NEW: outcome‑aware covariates, anchored at time zero (Patch C) ---
            all_eps = pd.concat([
                seq_df[['person_id','index_date']],
                comp_df[['person_id','index_date']],
                reverse_df[['person_id','index_date']],
                a_no_b_df[['person_id','index_date']],
                baseline_df[['person_id','index_date']]
            ], ignore_index=True).drop_duplicates()

            idx_series = pd.Series(all_eps['index_date'].values, index=all_eps['person_id'].values)
            cov_df = get_covariates_at_index(
                idx_series,
                event_A_vid=event_A,
                event_B_vid=event_B,
                outcome_name=str(outcome)
            )

            if 'index_date' in cov_df.columns:
                cov_df['index_date'] = pd.to_datetime(cov_df['index_date'], errors='coerce')

            for name, _df in (('seq', seq_df), ('comp', comp_df), ('rev', reverse_df), ('ter', a_no_b_df), ('base', baseline_df)):
                if _df.empty:
                    continue
                _df = _df.merge(cov_df, on=['person_id','index_date'], how='left')
                if name == 'seq':       seq_df = _df
                elif name == 'comp':    comp_df = _df
                elif name == 'rev':     reverse_df = _df
                elif name == 'ter':     a_no_b_df = _df
                else:                   baseline_df = _df

            # Also add demographics needed by CONFIG.COVARIATES_FOR_ADJUSTMENT
            demogs = persons_df[['person_id','sex_at_birth','race_ethnicity']]
            seq_df      = seq_df.merge(demogs, on='person_id', how='left')
            comp_df     = comp_df.merge(demogs, on='person_id', how='left')
            if not reverse_df.empty:
                reverse_df = reverse_df.merge(demogs, on='person_id', how='left')
            if not a_no_b_df.empty:
                a_no_b_df = a_no_b_df.merge(demogs, on='person_id', how='left')
            if not baseline_df.empty:
                baseline_df = baseline_df.merge(demogs, on='person_id', how='left')


            # Require adequate comparator size for the primary comparison
            if len(seq_df) < CONFIG.MIN_SEQUENCE_SUPPORT or len(comp_df) < CONFIG.MIN_SEQUENCE_SUPPORT:
                if _progress: _progress.update(1)
                continue

            # --- PRIMARY weights: A→B vs B-no-prior-A ---
            seq_p  = seq_df.copy();  seq_p['group']  = 1
            comp_p = comp_df.copy(); comp_p['group'] = 0
            combined_primary = pd.concat([seq_p, comp_p], ignore_index=True)

            # Demographics were already merged into seq_df/comp_df above.
            # Coalesce any lingering *_x/_y columns and ensure covariates exist.
            combined_primary = _coalesce_demog_cols(combined_primary)
            combined_primary = _ensure_covariate_columns(combined_primary)

            Xp = pd.get_dummies(
                combined_primary[CONFIG.COVARIATES_FOR_ADJUSTMENT],
                dummy_na=True, drop_first=True
            )
            yp = combined_primary['group']

            model_p = LogisticRegression(solver='liblinear',
                                         random_state=CONFIG.RNG_SEED,
                                         max_iter=1000)
            model_p.fit(Xp, yp)
            ps_p = model_p.predict_proba(Xp)[:, 1]
            w_p  = np.where(yp == 1, 1/ps_p, 1/(1-ps_p))
            w_p  = np.clip(w_p, None, np.nanpercentile(w_p, 99))
            combined_primary['iptw_weight_primary'] = w_p

            # Map primary weights back to seq & comp by (person_id, index_date)
            key_cols = ['person_id','index_date']
            w_primary = combined_primary[key_cols + ['iptw_weight_primary']]
            seq_df  = seq_df.merge(w_primary,  on=key_cols, how='left')
            comp_df = comp_df.merge(w_primary, on=key_cols, how='left')

            # --- REVERSE weights: A→B vs B→A (if reverse cohort viable) ---
            if not reverse_df.empty and len(reverse_df) >= CONFIG.MIN_SEQUENCE_SUPPORT:
                seq_r  = seq_df.copy();    seq_r['group']  = 1
                rev_r  = reverse_df.copy(); rev_r['group'] = 0

                combined_reverse = pd.concat([seq_r, rev_r], ignore_index=True)

                # Demographics were already merged into seq_df/reverse_df above.
                # Coalesce any lingering *_x/_y columns and ensure covariates exist.
                combined_reverse = _coalesce_demog_cols(combined_reverse)
                combined_reverse = _ensure_covariate_columns(combined_reverse)

                Xr = pd.get_dummies(
                    combined_reverse[CONFIG.COVARIATES_FOR_ADJUSTMENT],
                    dummy_na=True, drop_first=True
                )
                yr = combined_reverse['group']

                model_r = LogisticRegression(solver='liblinear',
                                             random_state=CONFIG.RNG_SEED,
                                             max_iter=1000)
                model_r.fit(Xr, yr)
                ps_r = model_r.predict_proba(Xr)[:, 1]
                w_r  = np.where(yr == 1, 1/ps_r, 1/(1-ps_r))
                w_r  = np.clip(w_r, None, np.nanpercentile(w_r, 99))
                combined_reverse['iptw_weight_reverse'] = w_r

                w_reverse = combined_reverse[key_cols + ['iptw_weight_reverse']]
                seq_df      = seq_df.merge(     w_reverse, on=key_cols, how='left')
                reverse_df  = reverse_df.merge( w_reverse, on=key_cols, how='left')
            else:
                # Ensure column exists for downstream code
                reverse_df['iptw_weight_reverse'] = np.nan

            # --- TERTIARY weights: A->B vs A_no_B (if viable) ---
            if not a_no_b_df.empty and len(a_no_b_df) >= CONFIG.MIN_SEQUENCE_SUPPORT:
                seq_t  = seq_df.copy();    seq_t['group']  = 1
                ter_t  = a_no_b_df.copy(); ter_t['group'] = 0
                combined_ter = pd.concat([seq_t, ter_t], ignore_index=True)
                combined_ter = _coalesce_demog_cols(combined_ter)
                combined_ter = _ensure_covariate_columns(combined_ter)
                Xt = pd.get_dummies(combined_ter[CONFIG.COVARIATES_FOR_ADJUSTMENT], dummy_na=True, drop_first=True)
                yt = combined_ter['group']
                model_t = LogisticRegression(solver='liblinear', random_state=CONFIG.RNG_SEED, max_iter=1000)
                model_t.fit(Xt, yt)
                ps_t = model_t.predict_proba(Xt)[:, 1]
                w_t  = np.where(yt == 1, 1/ps_t, 1/(1-ps_t))
                w_t  = np.clip(w_t, None, np.nanpercentile(w_t, 99))
                combined_ter['iptw_weight_tertiary'] = w_t
                w_ter = combined_ter[key_cols + ['iptw_weight_tertiary']]
                seq_df    = seq_df.merge(w_ter, on=key_cols, how='left')
                a_no_b_df = a_no_b_df.merge(w_ter, on=key_cols, how='left')
            else:
                a_no_b_df['iptw_weight_tertiary'] = np.nan

            # --- BASELINE weights: A->B vs Baseline (if viable) ---
            if not baseline_df.empty and len(baseline_df) >= CONFIG.MIN_SEQUENCE_SUPPORT:
                seq_q  = seq_df.copy();      seq_q['group']  = 1
                base_q = baseline_df.copy(); base_q['group'] = 0
                combined_base = pd.concat([seq_q, base_q], ignore_index=True)
                combined_base = _coalesce_demog_cols(combined_base)
                combined_base = _ensure_covariate_columns(combined_base)
                Xq = pd.get_dummies(combined_base[CONFIG.COVARIATES_FOR_ADJUSTMENT], dummy_na=True, drop_first=True)
                yq = combined_base['group']
                model_q = LogisticRegression(solver='liblinear', random_state=CONFIG.RNG_SEED, max_iter=1000)
                model_q.fit(Xq, yq)
                ps_q = model_q.predict_proba(Xq)[:, 1]
                w_q  = np.where(yq == 1, 1/ps_q, 1/(1-ps_q))
                w_q  = np.clip(w_q, None, np.nanpercentile(w_q, 99))
                combined_base['iptw_weight_baseline'] = w_q
                w_base = combined_base[key_cols + ['iptw_weight_baseline']]
                seq_df      = seq_df.merge(w_base, on=key_cols, how='left')
                baseline_df = baseline_df.merge(w_base, on=key_cols, how='left')
            else:
                baseline_df['iptw_weight_baseline'] = np.nan

            # --- Unified analysis frame with BOTH weight columns ---
            for df in (seq_df, comp_df, reverse_df, a_no_b_df, baseline_df):
                if 'iptw_weight_primary' not in df.columns:
                    df['iptw_weight_primary'] = np.nan
                if 'iptw_weight_reverse' not in df.columns:
                    df['iptw_weight_reverse'] = np.nan
                if 'iptw_weight_tertiary' not in df.columns:
                    df['iptw_weight_tertiary'] = np.nan
                if 'iptw_weight_baseline' not in df.columns:
                    df['iptw_weight_baseline'] = np.nan

            analysis_df = pd.concat([seq_df, comp_df, reverse_df, a_no_b_df, baseline_df], ignore_index=True)

            # Run competing-risk analysis once per outcome using analyze_sequence_outcome
            results_list, _ = analyze_sequence_outcome(analysis_df, outcome)

            # Attach sequence id and sensitivity params
            for res in results_list:
                res['sequence_idx'] = int(seq_idx)
                res.update(params)
                all_sensitivity_results.append(res)

        except Exception as e:
            print(f"    - ERROR on seq {seq_idx}, params {params}: {e}")
        finally:
            if _progress: _progress.update(1)

if _progress:
    try:
        _progress.close()
    except Exception:
        pass

# --- 6. Save results ---
if not all_sensitivity_results:
    print("  • No sensitivity results were generated.")
    pd.DataFrame().to_parquet(output_path)
else:
    sensitivity_df = pd.DataFrame(all_sensitivity_results)
    # Convenience: compute RR vs. reverse for quick inspection
    with np.errstate(divide='ignore', invalid='ignore'):
        sensitivity_df['risk_ratio_reverse'] = sensitivity_df['cif_seq'] / sensitivity_df['cif_reverse']
    sensitivity_df['risk_ratio_reverse'].replace([np.inf, -np.inf], np.nan, inplace=True)

    sensitivity_df.to_parquet(output_path, index=False)
    print(f"  • Saved sensitivity analysis results to: {output_path.name} ({len(sensitivity_df):,} rows).")

# --- 7. Sample display (optional) ---
try:
    sdf = pd.read_parquet(output_path)
    if not sdf.empty:
        cols = [
            'sequence_idx','outcome','horizon_years',
            'latency_days','max_gap_days',
            'risk_ratio','risk_ratio_reverse','delta_cif'
        ]
        cols = [c for c in cols if c in sdf.columns]
        print("\n--- Sample of Sensitivity Analysis Results ---")
        display(sdf[cols].head(10))
        print("---------------------------------------------")
except Exception:
    pass

print("✅ Cell 12 complete: Sensitivity analysis finished with OP‑aware episode construction.")


# In[ ]:


# ──────────────────────────── Cell 13 ────────────────────────────
# Generate Final Suppressed Results Table
# ---------------------------------------------------------------------------
#
#  1. **Suppression:** Any count of individuals less than 20 is suppressed.
#  2. **Rounding:** All remaining counts are rounded down to the nearest
#     multiple of 5.
#  3. **Propagation:** Suppression is propagated to all derived metrics. If a
#     count is suppressed, the corresponding risk estimates, CIs, and q-value
#     are also suppressed.

import pandas as pd
import numpy as np
from pathlib import Path

print("▶ Cell 13: Generating Final Suppressed Results Table...")

# --- 1. Prerequisite Checks and Path Definitions ---
required_globals = ['CONFIG', 'log']
for var in required_globals:
    if var not in globals():
        raise NameError(f"❌ Prerequisite '{var}' not found. Please run preceding cells first.")

if 'vocab_df' not in globals():
    try:
        vocab_df = pd.read_parquet(CONFIG.SOURCE_CACHE_DIR / "event_vocab_v1.parquet")
    except (FileNotFoundError, KeyError):
        raise FileNotFoundError("Could not find `event_vocab_v1.parquet` in the source cache.")
if 'sequence_map_df' not in globals():
    sequence_map_df = pd.read_parquet(CONFIG.ARTIFACTS_DIR / "sequence_map_df.parquet")


raw_results_path = CONFIG.ARTIFACTS_DIR / "final_results_with_q.parquet"
output_path      = CONFIG.RESULTS_DIR / "final_results_publication.csv"

if not raw_results_path.exists():
    raise FileNotFoundError(f"The raw results file (with q-values) was not found at: {raw_results_path}. Run Cell 11b first.")

# --- 2. Load Data and Add Human-Readable Names ---
print(f"  • Loading raw confirmation results from: {raw_results_path.name}")
results_df = pd.read_parquet(raw_results_path)

if results_df.empty:
    print("  • Raw results file is empty. No publication table will be generated.")
    # Create an empty file to satisfy downstream dependencies
    pd.DataFrame().to_csv(output_path, index=False)
    # Exit cleanly
    raise SystemExit("Exiting Cell 13 as there are no results to process.")

print(f"    - Loaded {len(results_df):,} result rows.")

# Add human-readable event names
results_df = results_df.merge(sequence_map_df, on='sequence_idx', how='left')
vocab_lookup = vocab_df.set_index('concept_id')['concept_name'].to_dict()
results_df['event_A_name'] = results_df['event_A_vid'].map(vocab_lookup)
results_df['event_B_name'] = results_df['event_B_vid'].map(vocab_lookup)
results_df['sequence_readable'] = results_df['event_A_name'].fillna('Unknown') + " → " + results_df['event_B_name'].fillna('Unknown')

# Risk ratio vs reverse (point estimate)
if "risk_ratio_reverse" not in results_df.columns and {"cif_seq","cif_reverse"}.issubset(results_df.columns):
    with np.errstate(divide="ignore", invalid="ignore"):
        results_df["risk_ratio_reverse"] = results_df["cif_seq"] / results_df["cif_reverse"]
    results_df["risk_ratio_reverse"].replace([np.inf, -np.inf], np.nan, inplace=True)


# --- 3. Define and Apply Suppression and Rounding Rules ---
print("  • Applying AoU suppression and rounding rules...")

# ### MODIFIED ###: Expanded list of all count columns to process
count_cols_to_process = [
    'n_seq','n_cases_seq','n_competing_seq',
    'n_comp','n_cases_comp','n_competing_comp',
    'n_reverse','n_cases_reverse','n_competing_reverse',
    'n_tertiary','n_cases_tertiary','n_competing_tertiary',
    'n_baseline','n_cases_baseline','n_competing_baseline',
    'at_risk_seq','at_risk_comp','at_risk_reverse',
    'at_risk_tertiary','at_risk_baseline',
    'events_seq','events_comp','events_reverse',
    'events_tertiary','events_baseline'
]
existing_count_cols = [col for col in count_cols_to_process if col in results_df.columns]

def suppress_and_round(series: pd.Series) -> pd.Series:
    suppressed = series.where(series >= CONFIG.MIN_CELL_SIZE)
    rounded = (suppressed // CONFIG.ROUND_TO_NEAREST) * CONFIG.ROUND_TO_NEAREST
    return rounded.astype('Int64')

for col in existing_count_cols:
    results_df[f"{col}_suppressed"] = suppress_and_round(results_df[col])

# --- 4. Propagate Suppression to Derived Metrics ---
print("  • Propagating suppression to risk estimates, CIs, and q-values...")

seq_suppressed_mask    = results_df['n_cases_seq_suppressed'].isna()
comp_suppressed_mask   = results_df['n_cases_comp_suppressed'].isna()
rev_suppressed_mask    = results_df['n_cases_reverse_suppressed'].isna() if 'n_cases_reverse_suppressed' in results_df else pd.Series(False, index=results_df.index)
ter_suppressed_mask    = results_df['n_cases_tertiary_suppressed'].isna() if 'n_cases_tertiary_suppressed' in results_df else pd.Series(False, index=results_df.index)
base_suppressed_mask   = results_df['n_cases_baseline_suppressed'].isna() if 'n_cases_baseline_suppressed' in results_df else pd.Series(False, index=results_df.index)

metric_groups = {
    'primary': (seq_suppressed_mask | comp_suppressed_mask, ['cif_seq', 'cif_comp', 'delta_cif', 'risk_ratio', 'p_value', 'q_value_confirm']),
    'reverse': (seq_suppressed_mask | rev_suppressed_mask, ['cif_reverse', 'risk_ratio_reverse']),
    'tertiary':(seq_suppressed_mask | ter_suppressed_mask, ['cif_tertiary', 'delta_cif_tertiary', 'risk_ratio_tertiary', 'p_value_tertiary']),
    'baseline':(seq_suppressed_mask | base_suppressed_mask, ['cif_baseline', 'delta_cif_baseline', 'risk_ratio_baseline', 'p_value_baseline'])
}

for group, (mask, cols) in metric_groups.items():
    for col_stem in cols:
        for suffix in ['', '_ci_lower', '_ci_upper']:
            full_col = col_stem + suffix
            if full_col in results_df.columns:
                results_df.loc[mask, full_col] = np.nan

# --- 5. Create Final Publication-Ready DataFrame ---
print("  • Assembling the final publication table...")

# ### MODIFIED ###: Expanded list to include all comparators from the start
final_col_candidates = [
    'sequence_readable', 'outcome', 'horizon_years',
    'n_seq_suppressed', 'n_cases_seq_suppressed',
    'n_comp_suppressed', 'n_cases_comp_suppressed',
    'n_reverse_suppressed', 'n_cases_reverse_suppressed',
    'n_tertiary_suppressed', 'n_cases_tertiary_suppressed',
    'n_baseline_suppressed', 'n_cases_baseline_suppressed',
    'cif_seq', 'cif_comp', 'cif_reverse', 'cif_tertiary', 'cif_baseline',
    'delta_cif', 'risk_ratio', 'risk_ratio_reverse',
    'delta_cif_tertiary', 'risk_ratio_tertiary',
    'delta_cif_baseline', 'risk_ratio_baseline',
    'delta_cif_ci_lower', 'delta_cif_ci_upper',
    'risk_ratio_ci_lower', 'risk_ratio_ci_upper',
    'q_value_confirm'
]
existing_cols = [col for col in final_col_candidates if col in results_df.columns]
publication_df = results_df[existing_cols].copy()

# Create formatted CI strings
if all(c in publication_df.columns for c in ['delta_cif_ci_lower', 'delta_cif_ci_upper']):
    publication_df['delta_cif_ci_95'] = publication_df.apply(
        lambda row: f"({row['delta_cif_ci_lower']:.3f}, {row['delta_cif_ci_upper']:.3f})" if pd.notna(row['delta_cif_ci_lower']) else np.nan, axis=1
    )
if all(c in publication_df.columns for c in ['risk_ratio_ci_lower', 'risk_ratio_ci_upper']):
    publication_df['risk_ratio_ci_95'] = publication_df.apply(
        lambda row: f"({row['risk_ratio_ci_lower']:.2f}, {row['risk_ratio_ci_upper']:.2f})" if pd.notna(row['risk_ratio_ci_lower']) else np.nan, axis=1
    )

# ### MODIFIED ###: Expanded rename dictionary
publication_df = publication_df.rename(columns={
    'sequence_readable': 'Sequence',
    'outcome': 'Outcome',
    'horizon_years': 'Horizon (Years)',
    'n_seq_suppressed': 'N (A→B)',
    'n_cases_seq_suppressed': 'Cases (A→B)',
    'cif_seq': 'CIF (A→B)',
    'n_comp_suppressed': 'N (B, no prior A)',
    'n_cases_comp_suppressed': 'Cases (B, no prior A)',
    'cif_comp': 'CIF (B, no prior A)',
    'n_reverse_suppressed': 'N (B→A)',
    'n_cases_reverse_suppressed': 'Cases (B→A)',
    'cif_reverse': 'CIF (B→A)',
    'n_tertiary_suppressed': 'N (A, no subseq. B)',
    'n_cases_tertiary_suppressed': 'Cases (A, no subseq. B)',
    'cif_tertiary': 'CIF (A, no subseq. B)',
    'n_baseline_suppressed': 'N (Baseline)',
    'n_cases_baseline_suppressed': 'Cases (Baseline)',
    'cif_baseline': 'CIF (Baseline)',
    'delta_cif': 'Risk Difference (vs Primary)',
    'risk_ratio': 'Risk Ratio (vs Primary)',
    'risk_ratio_reverse': 'Risk Ratio (vs Reverse)',
    'risk_ratio_tertiary': 'Risk Ratio (vs A, no subseq. B)',
    'risk_ratio_baseline': 'Risk Ratio (vs Baseline)',
    'q_value_confirm': 'q-value (Confirmation)',
    'delta_cif_ci_95': '95% CI (Risk Difference)',
    'risk_ratio_ci_95': '95% CI (Risk Ratio)'
})

# ### MODIFIED ###: Expanded list for final column ordering
final_pub_cols_ordered = [
    'Sequence', 'Outcome', 'Horizon (Years)',
    'N (A→B)', 'Cases (A→B)', 'CIF (A→B)',
    'N (B, no prior A)', 'Cases (B, no prior A)', 'CIF (B, no prior A)',
    'N (B→A)', 'Cases (B→A)', 'CIF (B→A)',
    'N (A, no subseq. B)', 'Cases (A, no subseq. B)', 'CIF (A, no subseq. B)',
    'N (Baseline)', 'Cases (Baseline)', 'CIF (Baseline)',
    'Risk Difference (vs Primary)', '95% CI (Risk Difference)',
    'Risk Ratio (vs Primary)', '95% CI (Risk Ratio)',
    'Risk Ratio (vs Reverse)',
    'Risk Ratio (vs A, no subseq. B)',
    'Risk Ratio (vs Baseline)',
    'q-value (Confirmation)'
]
final_pub_cols_exist = [col for col in final_pub_cols_ordered if col in publication_df.columns]
publication_df = publication_df[final_pub_cols_exist]

# --- 6. Save the Final Table to CSV ---
publication_df.to_csv(output_path, index=False, float_format='%.4f')
print(f"  • Successfully saved the final suppressed results to: {output_path}")

# --- 7. Final Summary ---
print("\n--- Final Table Summary ---")
print(f"Total rows in publication table: {len(publication_df):,}")
print("\nTop 10 rows from the final publication table:")
display(publication_df.head(10))
print("---------------------------")

print("✅ Cell 13 complete: Final publication-ready table has been generated.")
```
